# Six-month sequelae of post-vaccination SARS-CoV-2 infection: a retrospective cohort study of 10,024 breakthrough infections

**DOI:** 10.1101/2021.10.26.21265508

**Authors:** Maxime Taquet, Quentin Dercon, Paul J Harrison

## Abstract

Vaccination has proven effective against infection with SARS-CoV-2, as well as death and hospitalisation following COVID-19 illness. However, little is known about the effect of vaccination on other acute and post-acute outcomes of COVID-19. Data were obtained from the TriNetX electronic health records network (over 81 million patients mostly in the USA). Using a retrospective cohort study and time-to-event analysis, we compared the incidences of COVID-19 outcomes between individuals who received a COVID-19 vaccine (approved for use in the USA) at least 2 weeks before SARS-CoV-2 infection and propensity score-matched individuals unvaccinated for COVID-19 but who had received an influenza vaccine. Outcomes were ICD-10 codes representing documented COVID-19 sequelae in the 6 months after a confirmed SARS-CoV-2 infection (recorded between January 1 and August 31, 2021). Associations with the number of vaccine doses (1 vs. 2) and age (< 60 vs. ≥ 60 years-old) were assessed. Among 10,024 vaccinated individuals with SARS-CoV-2 infection, 9479 were matched to unvaccinated controls. Receiving at least one COVID-19 vaccine dose was associated with a significantly lower risk of respiratory failure, ICU admission, intubation/ventilation, hypoxaemia, oxygen requirement, hypercoagulopathy/venous thromboembolism, seizures, psychotic disorder, and hair loss (each as composite endpoints with death to account for competing risks; HR 0.70-0.83, Bonferroni-corrected p<.05), but not other outcomes, including long-COVID features, renal disease, mood, anxiety, and sleep disorders. Receiving 2 vaccine doses was associated with lower risks for most outcomes. Associations between prior vaccination and outcomes of SARS-CoV-2 infection were marked in those < 60 years-old, whereas no robust associations were observed in those ≥ 60 years-old. In summary, COVID-19 vaccination is associated with lower risk of several, but not all, COVID-19 sequelae in those with breakthrough SARS-CoV-2 infection. These benefits of vaccination were clear in younger people but not in the over-60s. The findings may inform service planning, contribute to forecasting public health impacts of vaccination programmes, and highlight the need to identify additional interventions for COVID-19 sequelae.

## Introduction

The observation that individuals can be infected with SARS-CoV-2 after being vaccinated against COVID-19 (so-called breakthrough infections) has caused concerns (Nixon and Ndhlovu, 2021). These concerns are mitigated by abundant evidence that the risk of severe COVID-19 illness (as proxied by hospitalisation, admission to intensive care unit, and mortality) is lessened by vaccination (Agrawal et al., 2021; Antonelli et al., 2021; Bahl et al., 2021; Butt et al., 2021; Cabezas et al., 2021; Glatman-Freedman et al., 2021; Haas et al., 2021; Hyams et al., 2021; Mateo-Urdiales et al., 2021; Roest et al., 2021). One case-control study investigated the association between self-reported SARS-CoV-2 infection in 908 pairs of vaccinated and unvaccinated individuals and self-reported symptoms beyond 28 days (Antonelli et al., 2021). It found that compared to unvaccinated individuals, those with breakthrough infection were at a lower risk of symptoms beyond 28 days.

However, how COVID-19 vaccination affects the broad spectrum of sequelae of SARS-CoV-2 infection remains elusive. In particular, it is unknown if vaccinated individuals are at the same risk as unvaccinated individuals of venous thromboembolisms, ischaemic strokes, neuropsychiatric complications, long-COVID presentations, and other post-acute sequelae following infection with SARS-CoV-2. In addition, unvaccinated people might have health behaviours related to vaccination hesitancy (Latkin et al., 2021), and this potential source of bias has not been addressed by previous studies.

This cohort study based on electronic health records compares the 6-months outcomes of SARS-CoV-2 infection among individuals who were (vs. those who were not) vaccinated against COVID-19. We investigated a range of outcomes (both acute and post-acute) with documented associations with COVID-19 across multiple body systems.

## Methods

### Data and study design

The study used TriNetX Analytics, a federated network of linked EHRs recording anonymized data from 59 healthcare organizations (HCOs), primarily in the USA, totalling 81 million patients. Available data include demographics, diagnoses (ICD-10 codes), procedures (Current Procedural Terminology [CPT] codes), and measurements (e.g. blood pressure). The HCOs consist in a mixture of primary care centres, hospitals, and specialist units. They provide data from uninsured as well as insured individuals. Data de-identification is attested to and receives a formal determination by a qualified expert as defined in Section §164.514(b)(1) of the HIPAA Privacy Rule. This formal determination supersedes TriNetX’s waiver from the Western Institutional Review Board (IRB). Using the TriNetX user interface, cohorts are created based on inclusion and exclusion criteria, matched for confounding variables, and compared for outcomes of interest over specified time periods. For further details about TriNetX, see Appendix pp. 1-2.

### Cohorts

Both the primary and control cohorts were defined as all patients who had, between January 1, 2021 and August 31, 2021, a confirmed SARS-CoV-2 infection, which we defined as either a confirmed diagnosis of COVID-19 (ICD-10 code U07.1) or a first positive PCR test for SARS-CoV-2. In the primary cohort, patients were included only if their confirmed SARS-CoV-2 infection occurred at least 14 days after a recorded administration of a COVID-19 vaccine approved for use in the USA (i.e. BNT162b2 ‘Pfizer/BioNTech’, mRNA-1273 ‘Moderna’, or Ad26.COV2.S ‘Janssen’). In the control cohort, patients were included only if no recorded vaccine against COVID-19 was recorded before their SARS-CoV-2 infection and if they had received a vaccine against influenza at any time. In the USA, the Centers for Disease Control and Prevention (CDC) recommend yearly influenza vaccination to everyone over the age of 6 months. This inclusion criterion thus excludes patients with obvious vaccine hesitancy (so-called ‘anti-vaxxers’) as this is correlated with other health-related behaviours that might confound associations with COVID-19 outcomes (Latkin et al., 2021). More details are provided in the Appendix pp. 2-4.

### Covariates

A set of established and suspected risk factors for COVID-19 and for more severe COVID-19 illness was used (de Lusignan et al., 2020; Taquet et al., 2021c; Williamson et al., 2020): age, sex, race, ethnicity, obesity, hypertension, diabetes, chronic kidney disease, asthma, chronic lower respiratory diseases, nicotine dependence, substance misuse, ischaemic heart disease and other forms of heart disease, socioeconomic deprivation, cancer (and haematological cancer in particular), chronic liver disease, stroke, dementia, organ transplant, rheumatoid arthritis, lupus, psoriasis, and disorders involving an immune mechanism. To capture these risk factors in patients’ health records, 55 variables were used. More details including ICD-10 codes are provided in the Appendix pp. 4-5. Cohorts were matched for all these variables, as described below. In addition, cohorts were stratified by the date of the SARS-CoV-2 infection in 2-monthly periods (January 1 to February 28, 2021, March 1 to April 30, 2021, May 1 to June 30, 2021, and July 1 to August 31, 2021) and matching was achieved independently within each period (guaranteeing that as many patients in the matched cohorts had their SARS-CoV-2 infection in every 2-month period).

### Outcomes

We investigated the 6-month incidence of all acute and post-acute outcomes which have been shown to be significantly associated with COVID-19 in four large-scale studies based on electronic health records (Al-Aly et al., 2021; Daugherty et al., 2021; Taquet et al., 2021b, 2021a), namely:

▪ Hospitalisation
▪ Intensive Care Unit (ICU) admission
▪ Death
▪ Intubation/Ventilation
▪ Respiratory failure
▪ Hypoxaemia
▪ Oxygen requirement
▪ Long COVID feature (any and each of the following as defined in Ref. 18)
  - Abdominal symptoms
  - Abnormal breathing
  - Anxiety/Depression
  - Chest/Throat pain
  - Cognitive symptoms
  - Fatigue
  - Headache
  - Myalgia
  - Other pain
▪ Hypertension
▪ Arrhythmia
▪ Cardiac failure
▪ Cardiomyopathy
▪ Myocarditis
▪ Coronary disease
▪ Hypercoagulopathy/Deep vein thrombosis (DVT)/Pulmonary embolism (PE)
▪ Ischaemic stroke
▪ Cerebral haemorrhage
▪ Peripheral neuropathy
▪ Seizures
▪ Type 2 diabetes mellitus
▪ Liver disease
▪ Kidney disease
▪ Interstitial lung disease
▪ Urticaria
▪ Sleep disorders
▪ Gastro-oesophageal reflux disease (GORD)
▪ Hair loss
▪ Hyperlipidaemia
▪ Joint pain
▪ Obesity
▪ Anosmia
▪ Nerve/nerve root/plexus disorder
▪ Myoneural junction/muscle disease
▪ Psychotic disorder
▪ Mood disorder
▪ Anxiety disorder

Each outcome was defined as the set of corresponding ICD-10, CPT, and VA Formulary codes as specified in the original study which showed their association with COVID-19. To account for death as a competing risk and thus address survivorship bias, each outcome was analysed as part of a composite outcome with death as the other component (Manja et al., 2017). While necessary to address survivorship bias, this approach is unlikely to provide insight into the risk of outcomes which are much rarer than death itself. The analysis was performed on October 12, 2021. More details, and ICD-10/CPT/VA Formulary codes, are provided in the Appendix pp. 5-6.

### Secondary analyses

We assessed whether the associations between prior vaccinations and outcomes of SARS-CoV-2 infection were moderated by the number of vaccine doses received and by age at the time of infection. This was achieved by restricting the primary cohort to (i) those who had received only one vaccine dose at least 14 days before infection, (ii) those who had received two vaccine doses 14 days before infection, (iii) those <60 years-old, and (iv) those ≥60 years-old. For the latter two subgroup analyses, the control cohorts were also restricted to those <60 and ≥60 years-old respectively.

### Statistical analyses

Propensity score matching (carried out within the TriNetX network) was used to create cohorts with matched baseline characteristics (Austin, 2011). Propensity score 1:1 matching used a greedy nearest neighbour approach with a caliper distance of 0.1 pooled standard deviations of the logit of the propensity score. Any characteristic with a standardized mean difference (SMD) between cohorts lower than 0.1 is considered well matched (Haukoos and Lewis, 2015). The Kaplan-Meier estimator was used to estimate the incidence of each outcome. Hazard ratios (HRs) with 95% confidence intervals were calculated using the Cox model and the null hypothesis of no difference between cohorts was tested using log-rank tests. The proportional hazard assumption was tested using the generalized Schoenfeld approach. When the assumption was violated, a time-varying HR was estimated using natural cubic splines fitted to the log-cumulative hazard (Royston and Parmar, 2002). The contribution of the individual outcomes of interest within the composite endpoint (with death as the other component) was reported as the number of events of interest over the total number of events (e.g. the number of respiratory failures over the number of respiratory failures or deaths).

We statistically tested whether the associations between prior COVID-19 vaccine and outcomes of SARS-CoV-2 infection was moderated by age. This was achieved for each outcome by testing whether the ratio between the HR in those ≥60 and the HR in those <60 years-old was statistically significantly different from 1. Similarly, moderation by the number of vaccine doses was achieved by testing whether the ratio between the HR in those who had received 1 dose and the HR in those who had received 2 doses at the time of infection was statistically significantly different from 1.

Further details are provided in the appendix p. 7. Statistical analyses were conducted in R version 3.6.3 except for the log-rank tests which were performed within TriNetX. Statistical significance was set at two-sided p-values <0.05. Bonferroni correction for multiple comparisons was applied to correct for the simultaneous assessments of 45 outcomes in the primary analysis. A REporting of studies Conducted using Observational Routinely-collected health Data (RECORD) statement was completed (see Appendix).

## Results

A total of 10,024 individuals with SARS-CoV-2 infections recorded at least 2 weeks after a first dose of vaccine against COVID-19 were identified (mean [SD] age at infection: 57.0 [17.9] years-old, 59.4% female). Among them, 65.1% were vaccinated with BNT162b2 ‘Pfizer/BioNTech’, 9.0% with mRNA-1273 ‘Moderna’, 1.6% with Ad26.COV2.S ‘Janssen’, and 24.4% with unspecified subtype. 9479 of these individuals were matched to 9479 individuals with SARS-CoV-2 infections who did not have a COVID-19 vaccine before SARS-CoV-2 infection. The main demographic features and comorbidities of both cohorts are summarised in table 1 (additional baseline characteristics presented in the appendix pp. 18-19). Adequate propensity-score matching (standardised mean difference < 0.1) was achieved for all comparisons and baseline characteristics and all subgroups (appendix pp. 18-27).

**Table 1.**
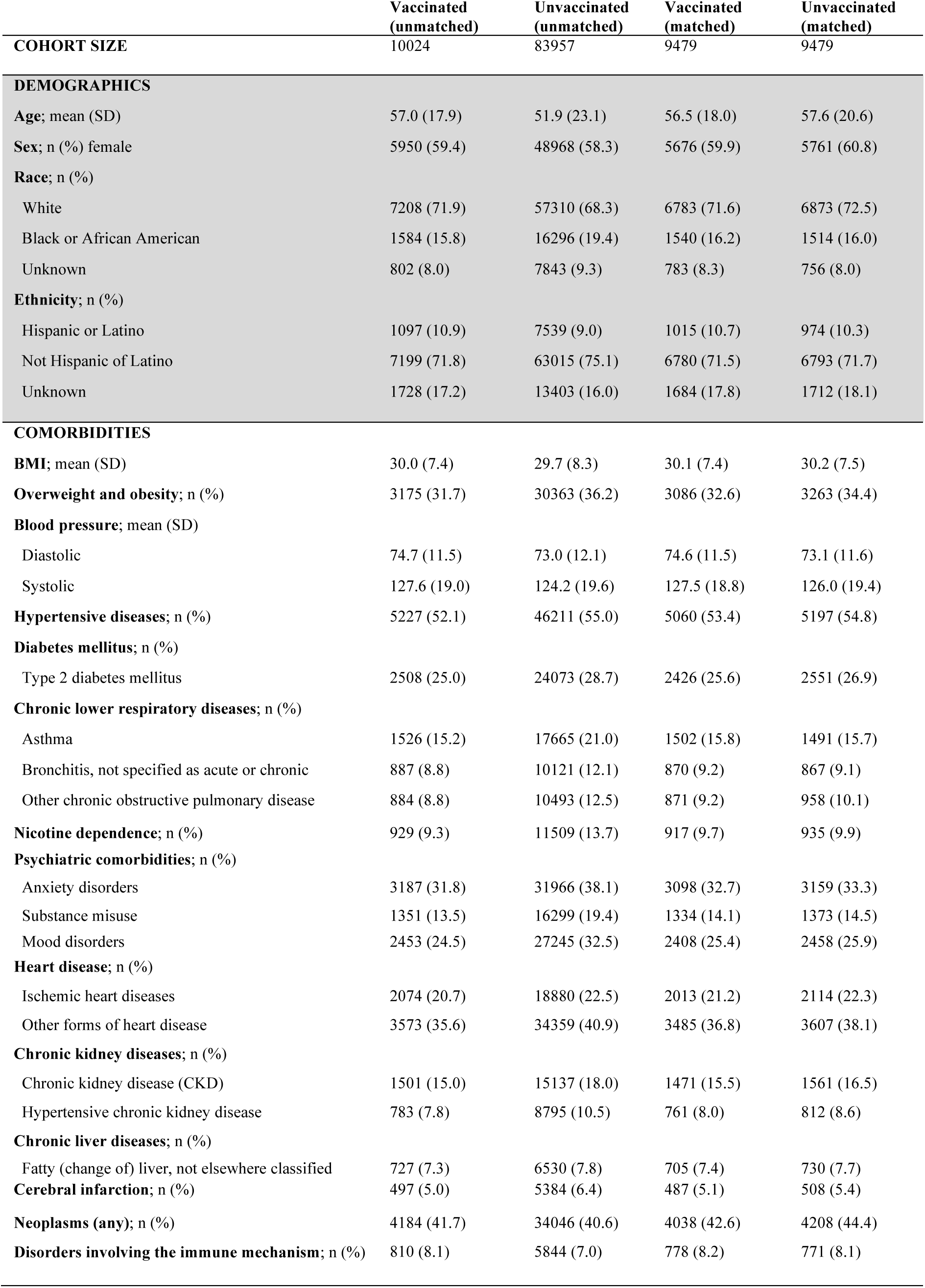
Baseline characteristics and major outcomes for the vaccinated and unvaccinated cohorts, before and after propensity score matching. Only characteristics with an overall prevalence above 5% after matching are presented here; for additional baseline characteristics see appendix pp. 18-19

|  | **Vaccinated**<br>**(unmatched)** | **Unvaccinated**<br>**(unmatched)** | **Vaccinated**<br>**(matched)** | **Unvaccinated**<br>**(matched)** |
| --- | --- | --- | --- | --- |
| **COHORT SIZE** | 10024 | 83957 | 9479 | 9479 |
| **DEMOGRAPHICS** |  |  |  |  |
| **Age; mean (SD)** | 57.0 (17.9) | 51.9 (23.1) | 56.5 (18.0) | 57.6 (20.6) |
| **Sex; n (%) female** | 5950 (59.4) | 48968 (58.3) | 5676 (59.9) | 5761 (60.8) |
| **Race; n (%)** |  |  |  |  |
| White | 7208 (71.9) | 57310 (68.3) | 6783 (71.6) | 6873 (72.5) |
| Black or African American | 1584 (15.8) | 16296 (19.4) | 1540 (16.2) | 1514 (16.0) |
| Unknown | 802 (8.0) | 7843 (9.3) | 783 (8.3) | 756 (8.0) |
| **Ethnicity; n (%)** |  |  |  |  |
| Hispanic or Latino | 1097 (10.9) | 7539 (9.0) | 1015 (10.7) | 974 (10.3) |
| Not Hispanic of Latino | 7199 (71.8) | 63015 (75.1) | 6780 (71.5) | 6793 (71.7) |
| Unknown | 1728 (17.2) | 13403 (16.0) | 1684 (17.8) | 1712 (18.1) |
| **COMORBIDITIES** |  |  |  |  |
| **BMI; mean (SD)** | 30.0 (7.4) | 29.7 (8.3) | 30.1 (7.4) | 30.2 (7.5) |
| **Overweight and obesity; n (%)** | 3175 (31.7) | 30363 (36.2) | 3086 (32.6) | 3263 (34.4) |
| **Blood pressure; mean (SD)** |  |  |  |  |
| Diastolic | 74.7 (11.5) | 73.0 (12.1) | 74.6 (11.5) | 73.1 (11.6) |
| Systolic | 127.6 (19.0) | 124.2 (19.6) | 127.5 (18.8) | 126.0 (19.4) |
| **Hypertensive diseases; n (%)** | 5227 (52.1) | 46211 (55.0) | 5060 (53.4) | 5197 (54.8) |
| **Diabetes mellitus; n (%)** |  |  |  |  |
| Type 2 diabetes mellitus | 2508 (25.0) | 24073 (28.7) | 2426 (25.6) | 2551 (26.9) |
| **Chronic lower respiratory diseases; n (%)** |  |  |  |  |
| Asthma | 1526 (15.2) | 17665 (21.0) | 1502 (15.8) | 1491 (15.7) |
| Bronchitis, not specified as acute or chronic | 887 (8.8) | 10121 (12.1) | 870 (9.2) | 867 (9.1) |
| Other chronic obstructive pulmonary disease | 884 (8.8) | 10493 (12.5) | 871 (9.2) | 958 (10.1) |
| **Nicotine dependence; n (%)** | 929 (9.3) | 11509 (13.7) | 917 (9.7) | 935 (9.9) |
| **Psychiatric comorbidities; n (%)** |  |  |  |  |
| Anxiety disorders | 3187 (31.8) | 31966 (38.1) | 3098 (32.7) | 3159 (33.3) |
| Substance misuse | 1351 (13.5) | 16299 (19.4) | 1334 (14.1) | 1373 (14.5) |
| Mood disorders | 2453 (24.5) | 27245 (32.5) | 2408 (25.4) | 2458 (25.9) |
| **Heart disease; n (%)** |  |  |  |  |
| Ischemic heart diseases | 2074 (20.7) | 18880 (22.5) | 2013 (21.2) | 2114 (22.3) |
| Other forms of heart disease | 3573 (35.6) | 34359 (40.9) | 3485 (36.8) | 3607 (38.1) |
| **Chronic kidney diseases; n (%)** |  |  |  |  |
| Chronic kidney disease (CKD) | 1501 (15.0) | 15137 (18.0) | 1471 (15.5) | 1561 (16.5) |
| Hypertensive chronic kidney disease | 783 (7.8) | 8795 (10.5) | 761 (8.0) | 812 (8.6) |
| **Chronic liver diseases; n (%)** |  |  |  |  |
| Fatty (change of) liver, not elsewhere classified | 727 (7.3) | 6530 (7.8) | 705 (7.4) | 730 (7.7) |
| **Cerebral infarction; n (%)** | 497 (5.0) | 5384 (6.4) | 487 (5.1) | 508 (5.4) |
| **Neoplasms (any); n (%)** | 4184 (41.7) | 34046 (40.6) | 4038 (42.6) | 4208 (44.4) |
| **Disorders involving the immune mechanism; n (%)** | 810 (8.1) | 5844 (7.0) | 778 (8.2) | 771 (8.1) |

We estimated the HRs for the occurrence within 6 months of infection of a range of health events (combined with death in composite endpoints) previously documented to occur at an increased rate after SARS-CoV-2 infections. As seen in Fig. 1 and Fig. 2 (see Appendix pp. 9-12 for all Kaplan-Meier curves and pp. 28-29 for a summary table), compared to unvaccinated individuals, those who were vaccinated at the time of SARS-CoV-2 infection were at a significantly lower risk of composite outcome of death and respiratory failure (HR 0.70, 95% CI 0.63-0.78, Bonferroni-corrected p<0.0001), intubation/ventilation (HR 0.72, 95% CI 0.61-0.84, Bonferroni-corrected p=0.0024), hypoxaemia (HR 0.72, 95% CI 0.65-0.80, Bonferroni-corrected p<0.0001), seizures (HR 0.73, 95% CI 0.62-0.86, Bonferroni-corrected p=0.0057), ICU admission (HR 0.75, 95% CI 0.65-0.85, Bonferroni-corrected p<0.0001), psychotic disorder (HR 0.75, 95% CI 0.63-0.89, Bonferroni-corrected p=0.036), hair loss (HR 0.75, 95% CI 0.64-0.88, Bonferroni-corrected p=0.024), hypercoagulopathy or venous thromboembolism (HR 0.81, 95% CI 0.72-0.91, Bonferroni-corrected p=0.014), and oxygen requirement (HR 0.83, 95% CI 0.75-0.92, Bonferroni-corrected p=0.011). In contrast, there was no significant difference in the risk of many other outcomes including composite of death and any long-COVID feature (HR 1.01, 95% CI 0.96-1.05, p=0.83, Bonferroni-corrected p=1.0), Type 2 diabetes mellitus (HR 0.97, 95% CI 0.90-1.05, p=0.45, Bonferroni-corrected p=1.0), mood disorder (HR 1.05, 95% CI 0.96-1.15, p=0.27, Bonferroni-corrected p=1.0), and anxiety disorder (HR 1.06, 95% CI 0.97-1.15, p=0.20, Bonferroni-corrected p=1.0), among others. There was no violation of the proportionality assumption for 35 out of 45 outcomes (Appendix pp. 30-31 and p. 13 for the time-varying HR for the other 10 outcomes). For most of the other 10 outcomes (including death), the time-varying HRs were significantly lower than 1 (indicating a stronger association with vaccination status) earlier in the follow-up window.

**Fig. 1.**
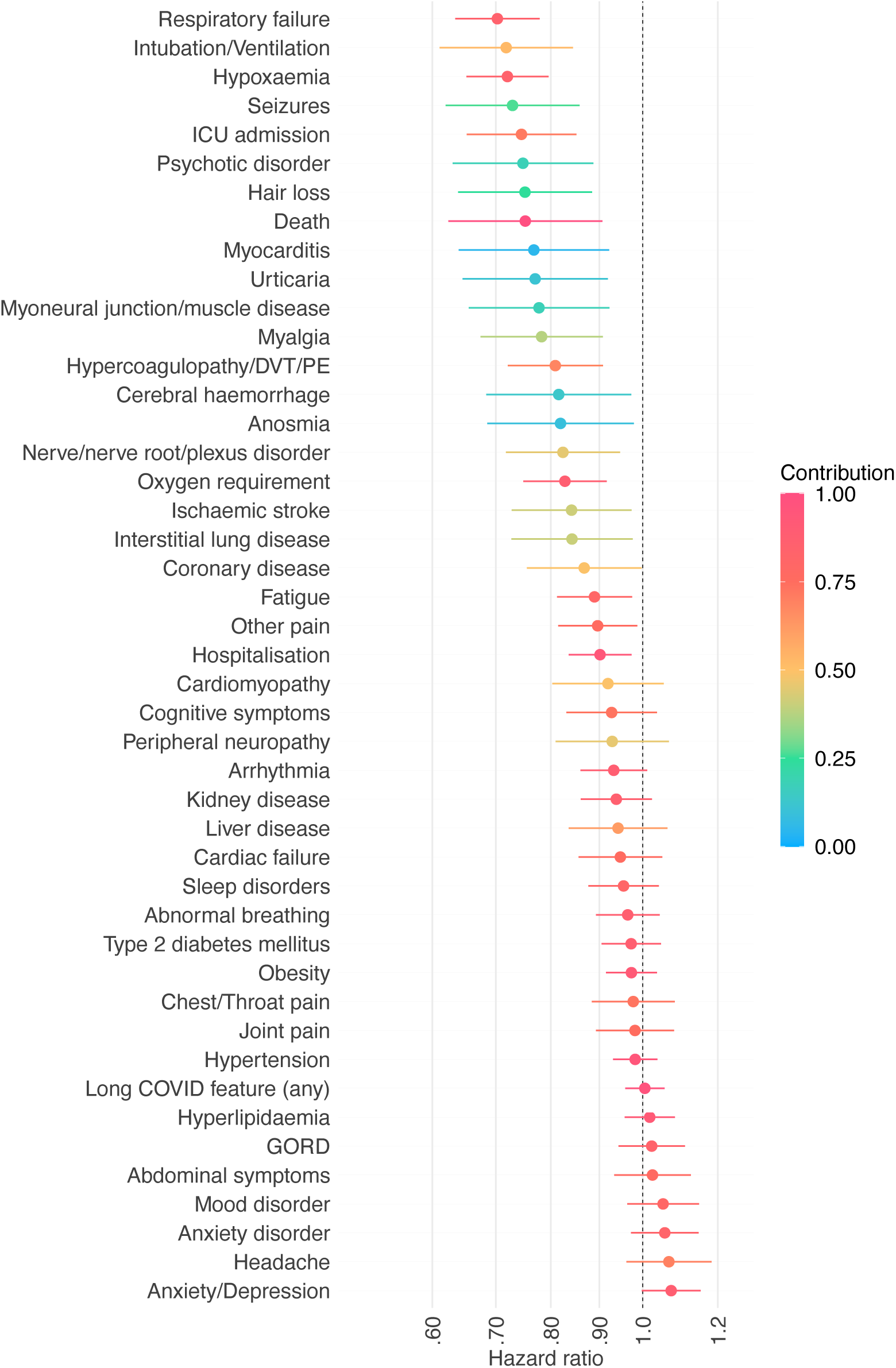
Hazard ratios for the outcome within 6 months of infection with SARS-CoV-2 between individuals vaccinated vs. unvaccinated against COVID-19. HR lower than 1 indicate outcomes less common among vaccinated individuals. Horizontal bars represent 95% confidence intervals. Each outcome is a composite endpoint with death as a component to address competing risks. The contribution of the outcome of interest to the overall incidence of the composite endpoint is encoded by the colour.

**Fig. 2.**
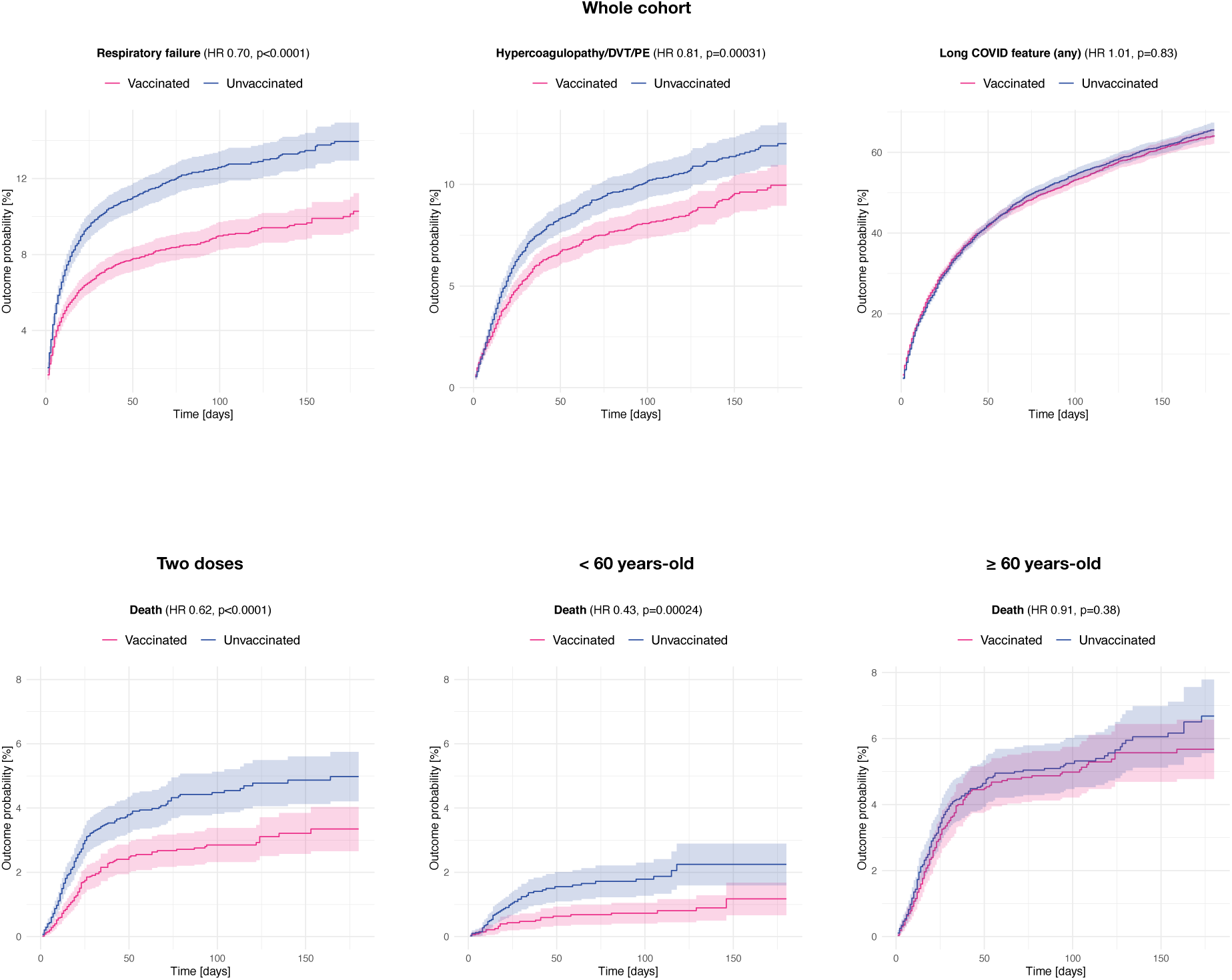
Kaplan-Meier estimates for the incidence of outcomes of SARS-CoV-2 infection between vaccinated and unvaccinated individuals in the whole cohort (top) and for the incidence of death in different subgroups (bottom). 95% confidence intervals are shaded. For Kaplan-Meier curves of all other outcomes in the whole cohort, see appendix pp. 9-12. DVT=Deep vein thrombosis. PE=Pulmonary embolism.

Besides the outcomes shown to be significantly associated with vaccination in the primary analysis, those who had received two vaccine doses at the time of SARS-CoV-2 infection were also at a significantly lower risk of myalgia, myocarditis, cerebral haemorrhage, interstitial lung disease, urticaria, anosmia, and myoneural junction/muscle disease (each as composite endpoints with death), and at a significantly lower risk of death (Fig. 3 and appendix pp. 14, 32-33). In those who had received only one vaccine dose, HRs followed a similar pattern, though they were generally closer to 1 (Fig. 3 and appendix pp. 15, 34-35). For each of the outcome, the HR in those who had received 2 doses was not statistically significantly different from the HR in those who had received 1 vaccine dose (appendix pp. 36-37).

**Fig. 3.**
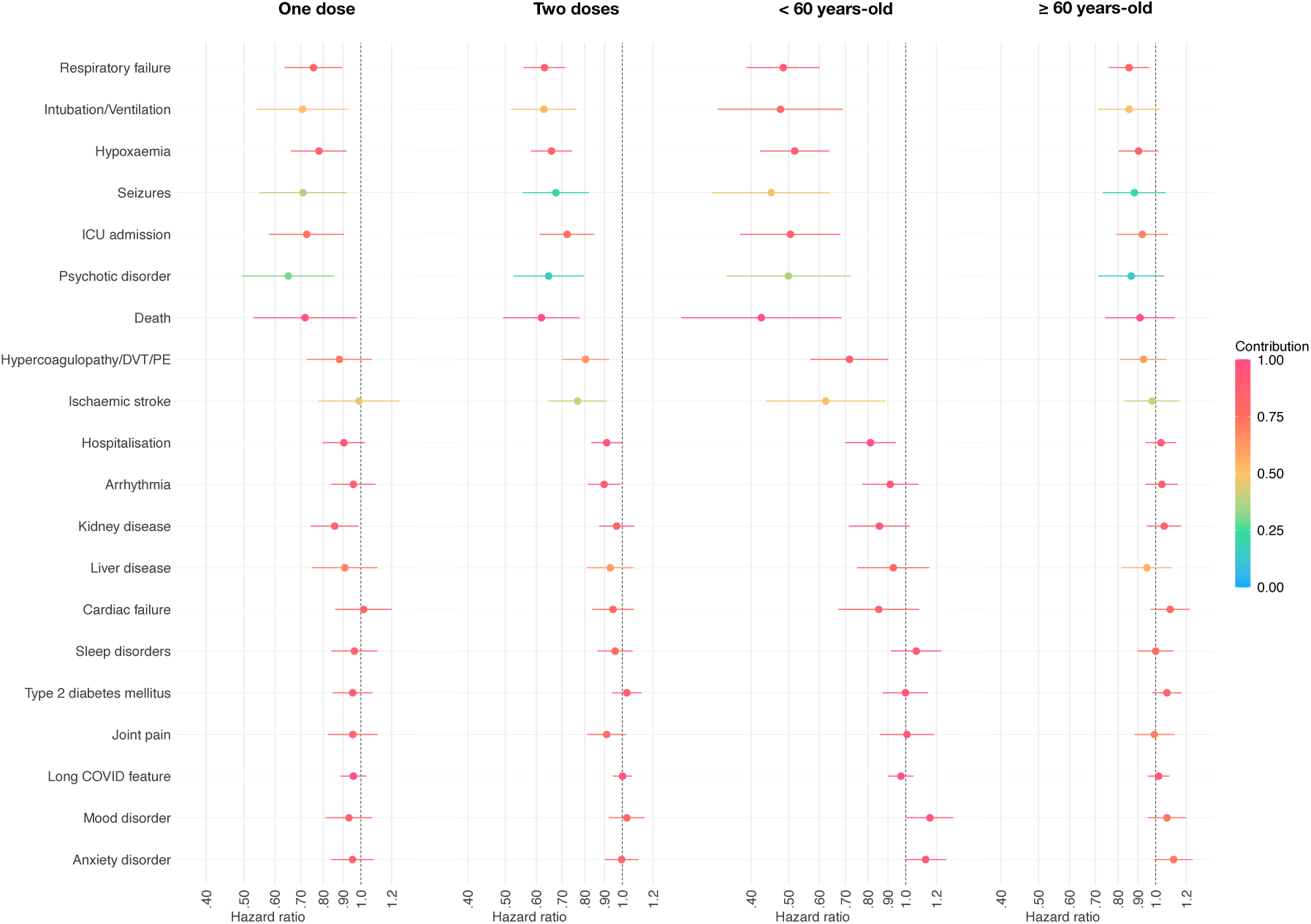
Hazard ratios for the outcomes within 6 months of infection with SARS-CoV-2 between individuals who received one dose of the vaccine (vs. unvaccinated individuals), those who received two doses of the vaccine (vs. unvaccinated individuals), vaccinated vs. unvaccinated individuals under the age of 60, and vaccinated vs. unvaccinated individuals over the age of 60. HR lower than 1 indicate outcomes less common among vaccinated individuals. Horizontal bars represent 95% confidence intervals. Each outcome is a composite endpoint with death as a component to address competing risks. The contribution of the outcome of interest to the overall incidence of the composite endpoint is encoded by the colour. Only a subset of representative outcomes is displayed. The same figures with all outcomes are presented in the appendix pp. 14-17.

We found a substantial effect of age on the results. Many HRs in younger individuals (< 60 years-old) were in general lower (i.e. favouring vaccination even more) for outcomes significantly associated with vaccination (Fig. 3 and appendix pp. 16, 38-39). These lower HRs were observed against the backdrop of lower *absolute* risks for most outcomes among younger individuals (appendix pp. 38-39). In contrast, in individuals ≥60 years-old, although most trends were similar to those in the younger group, none of the HRs were statistically significantly different from 1 after Bonferroni correction (Fig. 3 and appendix pp. 17, 40-41). For all but one outcomes associated with vaccination status in the primary analysis, the HR in the older group was significantly higher (i.e. favouring vaccination less) than the HR in the younger group (Appendix pp. 42-43). For instance, the HR for the composite outcome of death or respiratory failure was 0.48 (95% CI 0.39-0.60) in individuals <60 years-old and 0.85 (95% CI 0.76-0.96) in those ≥60 years-old (ratio between the two: 1.77, 95% CI 1.38-2.26, p<0.0001). The only exception was the composite of death and hypercoagulopathy/DVT/PE (HR in the older group 0.93, HR in the younger group 0.72, ratio between the two 1.30, 95% CI 1.00-1.70, p=0.054).

## Discussion

It is established that vaccination protects against hospitalisation, ICU admission, and death from COVID-19 (Agrawal et al., 2021; Antonelli et al., 2021; Bahl et al., 2021; Butt et al., 2021; Cabezas et al., 2021; Glatman-Freedman et al., 2021; Haas et al., 2021; Hyams et al., 2021; Mateo-Urdiales et al., 2021; Roest et al., 2021) but little was known about other outcomes of breakthrough SARS-CoV-2 infections. The data presented in this study, from a large-scale electronic health records network, confirm that vaccination protects against death and ICU admission following breakthrough SARS-CoV-2 infection and provide estimates of HR for these outcomes in a general population. Our study also shows that vaccination against COVID-19 is associated with lower risk of additional outcomes that had not been assessed in previous studies, namely respiratory failure, hypoxaemia, oxygen requirement, hypercoagulopathy or venous thromboembolism, seizures, psychotic disorder, and hair loss.

On the other hand, previous vaccination does not appear to be protective against several previously documented outcomes of COVID-19 such as long-COVID features, arrhythmia, joint pain, type 2 diabetes, liver disease, sleep disorders, and mood and anxiety disorders. The narrow confidence intervals (related to the high incidence of these outcomes post-COVID) rules out the possibility that these negative findings are merely a result of lack of statistical power. The inclusion of death in a composite endpoint with these outcomes rules out survivorship bias as an explanation. Instead, these negative findings might indicate that these outcomes arise through different pathophysiological mechanisms than outcomes which are affected by prior vaccination. For example, for anxiety and depression, it might be that antagonistic forces are at play with the protective effect of the vaccine being counteracted by the additional stressor of being infected despite being vaccinated.

The absence of a protective effect against long-COVID features is concerning given the high incidence and burden of these sequelae of COVID-19 (Taquet et al., 2021a). However, the risk of several individual long-COVID features were significantly associated with prior vaccination (but did not survive correction for multiple comparisons): myalgia (HR 0.78, 95% CI 0.67-0.91), fatigue (HR 0.89, 95% CI 0.81-0.97), and pain (HR 0.90, 95% CI 0.81-0.99), with potentially additional protection after a second dose of the vaccine against abnormal breathing (HR 0.89, 95% CI 0.81-0.98) and cognitive symptoms (HR 0.87, 95% CI 0.76-0.99). Relative differences in the incidence of individual long-COVID features might explain why our findings differ from those of an app-based survey suggesting that vaccination is overall protective against long-COVID symptoms (Antonelli et al., 2021). Other reasons might also explain this difference. First, the present study used data routinely collected from a general population rather than self-selected individuals. Second, this study uses a large sample size and matches cohorts on a broad range of recorded comorbidities. To further decrease selection bias, the control cohort of this study was selected among those having received an influenza vaccine thus helping control for the confounding effect of vaccine hesitancy and related health behaviours (Latkin et al., 2021). Third, by using symptoms and diagnoses extracted from health records rather than self-reported, our study might capture the most severe presentation of symptoms and these might be less prone to demand characteristics and other forms of detection bias. Finally, there might be differences in the type of vaccines used between the two study. In particular, no ChAdOx1 nCov-19 (‘Oxford/AstraZeneca’) vaccine was used in our study (as this vaccine is not used in the USA).

The findings that vaccination against SARS-CoV-2 does not protect against some of the post-acute outcomes of COVID-19 should not obscure the fact that vaccination remains an important protective factor against these outcomes at the population level, since the best way to prevent those outcomes is to prevent SARS-CoV-2 infection in the first place. This is also the case in those ≥ 60 years-old (Polack et al., 2020). However, our results highlight that some post-acute outcomes of SARS-CoV-2 (and notably long-COVID presentations) are likely to persist even after successful vaccination of the population, so long as breakthrough infections occur. These findings thus help in determining the necessary service provision. They also underline the urgency to identify other preventive or curative interventions to mitigate the impact of such COVID-19 sequelae.

Death was included in a composite endpoint with each outcome of interest to address competing risks (Manja et al., 2017). This implies that for outcomes much rarer than death (which is the minority of outcomes investigated), HRs might be driven by death rather than the outcome of interest. For instance, the 6-month incidence of psychotic disorders in the unvaccinated cohort was 1.11% whereas the death rate was 4.5% in this cohort. Hence the HR of 0.75 for the composite endpoint of death and psychotic disorder is in part driven by differences in death rate. Observing the HR for the outcome of interest in isolation (i.e. not composed with death) is informative so long as it is lower than 1. A HR lower than 1 suggests a lower rate of the outcome of interest despite survivorship bias (which in the present study increases HRs). If the HR is higher than 1, it is impossible to know whether the outcome is more common among vaccinated individuals or if it is a result of survivorship bias. HRs for all outcomes in isolation are provided in Appendix pp. 44-53 for the primary and secondary analyses. For instance, the HR for psychotic disorder measured in isolation was 0.79, indicating that the incidence of this outcome might indeed be lower in vaccinated than unvaccinated individuals and its significant HR might not be merely driven by differences in death rates.

Importantly, the protective effects of the vaccine against acute severity of infection and some post-acute sequelae appears to affect primarily those <60 years-old. In this group, the effects were large and robust, whereas in the <u>≥</u>60 year-old group, effects were smaller and not statistically robust. This finding regarding age cannot be explained by differences in statistical power since the younger and older subgroups have similar sample sizes (4633 and 4657 respectively) and outcomes are in general more frequent in the older subgroup. Instead, it might be due to differences in contributions of pathophysiological pathways of breakthrough infections between younger and older individuals. In younger patients, effective B-cell response to vaccination might be followed by infection with variants against which antibodies have less neutralising activity (Wu et al., 2021). In older patients, the B-cell response to vaccination might itself be ineffective (Siegrist and Aspinall, 2009) so that the clinical presentation of breakthrough infection is similar to that of infection in unvaccinated individuals. Regardless of explanation, the seemingly age-limited benefits of vaccination on the severity and sequelae of SARS-CoV-2 infection has clear public health implications.

This study has several limitations beyond those inherent to research using EHRs (Casey et al., 2016; Taquet et al., 2021b) (summarised in the appendix p. 2) such as the unknown completeness of records, no validation of diagnoses, and sparse information on socioeconomic and lifestyle factors. First, we do not know which SARS-CoV-2 variant individual patients were infected with and this might affect the protective effect of vaccines. There is evidence that variants of concerns are overrepresented in breakthrough infections (McEwen et al., 2021). Since such variants tend to be associated with worse outcomes (Nyberg et al., 2021), their enrichment in breakthrough infections means that some HRs presented in this study might be conservative estimates. Second, it might be that vaccination status affects the probability to seek or receive medical attention, particularly for less severe outcomes. Third, this study says nothing about the outcomes in patients infected with SARS-CoV-2 but who did not get tested nor diagnosed with COVID-19. Fourth, this study did not investigate whether the association between vaccination status and outcomes of SARS-CoV-2 infection was moderated by timing of the vaccine with respect to the infection. Fifth, we could not compare the different vaccines against each other since the majority had received the Pfizer/BioNTech vaccine. Finally, as an observational study, causation cannot be inferred (though the specificity of the association with some outcomes and not others and the dose-response relationship support causality as an explanation).

In summary, the present data show that prior vaccination against COVID-19, especially after two doses, is associated with significantly less risk of many but not all outcomes of COVID-19, in younger but not older individuals. These findings may inform service planning, contribute to forecasting public health impacts of vaccination programmes, and highlight the urgent need to identify or develop additional preventive and curative interventions for sequelae of COVID-19.

## Data Availability

The TriNetX system returned the results of these analyses as csv files which were downloaded and archived. Data presented in this paper and the Appendix can be freely accessed at [URL to be added on publication]. Additionally, TriNetX will grant access to researchers if they have a specific concern (via the third-party agreement option).

## Contributors

MT and PJH conceptualised the study, acquired funding, defined the methodology, and were involved in project administration and supervision. MT and QD curated the data, did the formal analyses, and validated the findings. MT created visualizations and wrote the original draft. MT, QD, and PJH reviewed and edited the manuscript for style and content. PJH and MT were granted unrestricted access to the TriNetX Analytics network for the purposes of research, and with no constraints on the analyses performed nor the decision to publish. The corresponding author had full access to all the data in the study, conducted the analysis, and takes responsibility for the integrity of the data and the accuracy of the data analysis.

## Acknowledgments

Work supported by the National Institute for Health Research (NIHR) Oxford Health Biomedical Research Centre (grant BRC-1215-20005). MT is an NIHR Academic Clinical Fellow and NIHR Oxford Health BRC Senior Research Fellow. QD is supported by the Medical Research Council (SUAG/077 G101400) and an AXA Research Fund Fellowship (G102329). The views expressed are those of the authors and not necessarily those of the UK National Health Service, NIHR, or the UK Department of Health.

