## Appendix for "Six-month sequelae of post-vaccination SARS-CoV-2 infection: a retrospective cohort study of 10,024 breakthrough infections"

### Supplementary Methods

#### TriNetX network

This section provides an expanded version of our previous description of the network.[1,2]

*Legal and ethical status*
TriNetX's Analytics network is compliant with the Health Insurance Portability and Accountability Act (HIPAA), the US federal law which protects the privacy and security of healthcare data. TriNetX is certified to the ISO 27001:2013 standard and maintains an Information Security Management System (ISMS) to ensure the protection of the healthcare data it has access to and to meet the requirements of the HIPAA Security Rule. Any data displayed on the TriNetX Platform in aggregate form, or any patient level data provided in a data set generated by the TriNetX Platform, only contains de-identified data as per the de-identification standard defined in Section §164.514(a) of the HIPAA Privacy Rule. The process by which the data is de-identified is attested to through a formal determination by a qualified expert as defined in Section §164.514(b)(1) of the HIPAA Privacy Rule. This formal determination by a qualified expert, refreshed in December 2020, supersedes the need for TriNetX's previous waiver from the Western Institutional Review Board (IRB). The network contains data that are provided by participating Health Care Organizations (HCOs), each of which represents and warrants that it has all necessary rights, consents, approvals and authority to provide the data to TriNetX under a Business Associate Agreement (BAA), so long as their name remains anonymous as a data source and their data are utilized for research purposes. The data shared through the TriNetX Platform are attenuated to ensure that they do not include sufficient information to facilitate the determination of which HCO contributed which specific information about a patient.

*Acquisition of data, quality control, and other procedures*
The data are stored onboard a TriNetX appliance – a physical server residing at the institution's data centre or a virtual hosted appliance. The TriNetX platform is a fleet of these appliances connected into a federated network able to broadcast queries to each appliance. Results are subsequently collected and aggregated.

Once the data are sent to the network, they are mapped to a standard and controlled set of clinical terminologies and undergo a data quality assessment including 'data cleaning' that rejects records which do not meet the TriNetX quality standards. HIPAA compliance of the clinical patient data is achieved using de-identification. Different data modalities are available in the network. They include demographics (coded to HL7 version 3 administrative standards), diagnoses (represented by ICD-10-CM codes), procedures (coded in ICD-10-PCS or CPT), and measurements (coded to LOINC). While extensive information is provided about patients' diagnoses and procedures, other variables (such as socioeconomic and lifetime factors are not comprehensively represented).

The data from a typical HCO generally go back around 7 years, with some going back 13 years. The data are continuously updated. HCOs update their data at various times, with most refreshing every 1, 2, or 4 weeks.

The data come primarily (>93%) from HCOs in the USA, with the remainder coming from India, Australia, Malaysia, Taiwan, Spain, UK, and Bulgaria. As noted above, to comply with legal frameworks and ethical guidelines guarding against data re-identification, the identity of participating HCOs and their individual contribution to each dataset are not disclosed to researchers.

Data quality assessment followed a standardised strategy wherein the data are reviewed for conformance (adherence to specified standards and formats), completeness (quantifying data presence or absence) and plausibility (believability of the data from a clinical perspective). There are pre-defined metrics for each of the above assessment categories. Results for these metrics are visualised and reviewed for each new site that joins the network as well as on an ongoing basis. Any identified issue is communicated to the data provider and resolved before continuing data collection.

The basic formatting of contributed data is also checked (e.g. to ensure that dates are properly represented). Records are checked against a list of required fields (e.g., patient identifier) and rejects those records for which the required information is missing. Referential integrity checking is done to ensure that data spanning multiple database tables can be successfully joined together. As the data are refreshed, changes in volume of data over time is monitored to ensure data validity. At least one non-demographic fact for each patient is required for them to be counted in the dataset. Patient records with only demographics information are discarded.

The software also undergoes quality control. The engineers testing the software are independent from the engineers developing it. Each test code is checked by two independent testing engineers. Each piece of software is tested extensively against a range of synthetic data (i.e. generated for the purpose of testing) for which the expected output is established independently. If the software fails to return this output, then the software is deemed to have failed the test and is examined and modified accordingly. For statistical software (including that used for propensity score matching, for Kaplan-Meier analysis, etc), an additional quality control step is implemented. Two independent codes are written in two different programming languages (typically R and python) and the statistical results are compared. If discrepancies are identified, then the codes are deemed to have failed the test and are examined and modified accordingly. All the code is reviewed independently by another engineer.

The test strategy follows three levels of granularity:

1. Unit tests: These test specific blocks, or units, of code that perform specific actions (e.g. querying the database).
2. Integration tests: These ensure that different components are working together correctly.
3. End-to-end tests: These tests run the entire system and check the final output.

*Some comments on advantages and disadvantages of EHR data*
One advantage of EHR data, like those in TriNetX, over insurance claim data is that both insured and uninsured patients are included. An advantage of EHR data over survey data is that they represent the diagnostic rates in the population presenting to healthcare facilities. This provides an accurate account of the burden of specific diagnoses on healthcare systems. However, there are also limitations inherent to research using electronic health records,[3–5] including TriNetX:

1. Patients with acute or post-acute sequelae of COVID-19 but were not diagnosed are not included leading to underestimation of actual incidences.
2. Despite the matching and use of various comparison cohorts, there may well be residual confounding, particularly related to social and economic factors which are not well captured in EHR networks and which might influence outcomes post COVID-19.
3. We do not know which diagnoses were made in primary or secondary care or specialist facilities, nor by whom.
4. A patient may be seen in different HCOs for different parts of their care, and if one HCO is not part of the federated network then part of their medical records may not be available. Using a network of HCOs (rather than a single HCO) limits this possibility but does not fully remove it.
5. How long a symptom/diagnosis persists is difficult to assess using EHR data as this is not typically coded. As a result, we can comment on incidence of new cases but cannot assess the duration of clinical features.
6. Since the data are presented as they are recorded, we cannot be sure that there has not been mis-recording of information, adding a degree of noise to the data.
7. Historical data before the start of EHRs (or the addition of an HCO to the network) may well be incomplete.

**Definition of cohorts**

*Definitions*

1) A confirmed diagnosis of COVID-19 was defined as the presence of ICD-10 code U07.1 in the patient' EHR.

2) A positive PCR test for SARS-CoV-2 was defined as any of the following:
    a. Positive SARS-CoV-2 RNA in Respiratory specimen
    b. Positive SARS-CoV-2 RNA in Unspecified specimen
    c. Positive SARS-CoV-2 N gene in Respiratory specimen
    d. Positive SARS-CoV-2 N gene in Unspecified specimen
    e. Positive SARS-CoV-2 RdRp gene in Respiratory specimen
    f. Positive SARS-CoV-2 E gene in Respiratory specimen

g. Positive SARS-CoV-2 E gene in Unspecified specimen
h. Positive SARS-CoV-2 RNA panel in Respiratory specimen
i. Positive SARS-CoV-2 RNA panel in Unspecified specimen
j. Positive SARS-CoV-2 RNA in Nasopharynx
k. Positive SARS coronavirus 2 and related RNA
l. Positive SARS-related coronavirus RNA in Respiratory specimen
m. Positive SARS coronavirus 2 ORF1ab in Respiratory specimen

3) An mRNA vaccine against COVID-19 was defined as any of the following:
   a. Severe acute respiratory syndrome coronavirus 2 (SARS-CoV-2) (Coronavirus disease [COVID-19]) vaccine, mRNA-LNP, spike protein, preservative free, 30 mcg/0.3mL dosage, diluent reconstituted, for intramuscular use
   b. Immunization administration by intramuscular injection of severe acute respiratory syndrome coronavirus 2 (SARS-CoV-2) (Coronavirus disease [COVID-19]) vaccine, mRNA-LNP, spike protein, preservative free, 30 mcg/0.3mL dosage, diluent reconstituted; first dose
   c. Immunization administration by intramuscular injection of severe acute respiratory syndrome coronavirus 2 (SARS-CoV-2) (Coronavirus disease [COVID-19]) vaccine, mRNA-LNP, spike protein, preservative free, 30 mcg/0.3mL dosage, diluent reconstituted; second dose
   d. Severe acute respiratory syndrome coronavirus 2 (SARS-CoV-2) (Coronavirus disease [COVID-19]) vaccine, mRNA-LNP, spike protein, preservative free, 100 mcg/0.5mL dosage, for intramuscular use
   e. Immunization administration by intramuscular injection of severe acute respiratory syndrome coronavirus 2 (SARS-CoV-2) (Coronavirus disease [COVID-19]) vaccine, mRNA-LNP, spike protein, preservative free, 100 mcg/0.5mL dosage; first dose
   f. Immunization administration by intramuscular injection of severe acute respiratory syndrome coronavirus 2 (SARS-CoV-2) (Coronavirus disease [COVID-19]) vaccine, mRNA-LNP, spike protein, preservative free, 100 mcg/0.5mL dosage; second dose
   g. Severe acute respiratory syndrome coronavirus 2 (SARS-CoV-2) (coronavirus disease [COVID-19]) vaccine, DNA, spike protein, adenovirus type 26 (Ad26) vector, preservative free, 5x10^10 viral particles/0.5mL dosage, for intramuscular use
   h. Immunization administration by intramuscular injection of severe acute respiratory syndrome coronavirus 2 (SARS-CoV-2) (coronavirus disease [COVID-19]) vaccine, DNA, spike protein, adenovirus type 26 (Ad26) vector, preservative free, 5x10^10 viral particles/0.5mL dosage, single dose
   i. SARS-CoV-2 (COVID-19) vaccine, mRNA spike protein
   j. SARS-CoV-2 (COVID-19) vaccine

*Inclusion and exclusion criteria for cohorts*

The primary cohort was defined as all individuals who met the following criteria:

A. The individual had a confirmed diagnosis of COVID-19 or a positive PCR test for SARS-CoV-2 between January 1, and August 31, 2021.

B. The individual had received an mRNA vaccine against COVID-19 at least two weeks before the first occurrence of Event A

The control cohort was defined as all individuals who met the following criteria:

A. The individual had a confirmed diagnosis of COVID-19 or a positive PCR test for SARS-CoV-2 between January 1, and August 31, 2021.

B. The individual had not received an mRNA vaccine against COVID-19 before any Event A

C. The individual had a recorded vaccine against influenza at some point in their EHR

Note that individuals who received a vaccine after infection with SARS-CoV-2 were included in the control cohort. This serves two purposes. First, it further decreases the risk of bias due to differences in health behaviour related to vaccine hesitancy. Second, a cohort defined by excluding those who have ever received a vaccine against COVID-19 would be enriched for people who have died shortly after infection with SARS-CoV-2 infection, thus introducing bias.

**Definition of covariates**

To reduce the effect of confounding on associations, cohorts were matched for established or suspected risk factors for COVID-19[6–9] and for established risk factors for COVID-19 death[10] (taken to be risk factors of a more severe COVID-19 illness). The following confounding factors were therefore included (with ICD-10/CDC codes in brackets):

1) **Age** at the time of diagnosis.
2) **Sex** coded as female, male, or other.
3) **Race** encoded as 6 separate dichotomous variables: White (2106-3), Black or African American (2054-5), American Indian or Alaska Native (1002-5), Asian (2028-9), Native Hawaiian or Other Pacific Islander (2076-8), or Unknown Race (2131-1).
4) **Ethnicity** encoded as Hispanic or Latino (2135-2), Not Hispanic or Latino (2186-5), or Unknown Ethnicity.
5) **Socioeconomic deprivation** encoded as the ICD-10 code for Problems related to housing and economic circumstances (Z59).
6) **Obesity** encoded as one dichotomous variable and one categorical variable: Overweight and obesity (E66) and body mass index (categorised into < 25 kg/m$^2$, 25-30 kg/m$^2$, ≥ 30 kg/m$^2$ which are the WHO thresholds for not obese, pre-obese, and obesity).
7) **Hypertension** encoded as 2 dichotomous and 2 categorical variables: Hypertensive diseases (I10-I16), the now deprecated version that was used until 2018 Hypertension diseases (I10-I15), measurements of systolic blood pressure (categorised into < 140mmHg, 140-160mmHg, and ≥ 160mmHg), and diastolic blood pressure (categorised into < 90mmHg, 90-100mmHg, and ≥ 100mmHg). The blood pressure categories correspond to the absence of hypertension, stage 1 hypertension, and stage 2 (and over) hypertension as per the NICE guidelines.
8) **Diabetes mellitus** encoded as 2 dichotomous variables: Type 1 diabetes mellitus (E10) and Type 2 diabetes mellitus (E11).
9) **Chronic lower respiratory diseases** encoded by each sub-category of the corresponding ICD-10 group: Bronchitis, not specified as acute or chronic (J40), Simple and mucopurulent chronic bronchitis (J41), Unspecified chronic bronchitis (J42), Emphysema (J43), Other chronic obstructive pulmonary disease (J44), Asthma (J45), Bronchiectasis (J47).
10) **Nicotine dependence** encoded as the corresponding ICD-10 diagnosis (F17.2).
11) **Substance use disorders** encoded as the ICD-10 code for mental and behavioural disorders due to psychoactive substance use (F10-F19).
12) **Psychotic disorders** encoded as the ICD-10 code for schizophrenia, schizotypal, delusional, and other non-mood psychotic disorders (F20-F29).
13) **Mood disorders** encoded as the ICD-10 code for mood disorders (F30-F39).
14) **Anxiety disorders** encoded as the ICD-10 code for anxiety, dissociative, stress-related, somatoform and other nonpsychotic mental disorders (F40-F48).
15) **Heart diseases** encoded as 2 categorical variables: Ischaemic heart disease (I20-I25) and Other forms of heart disease (I30-I52).
16) **Chronic kidney disease** encoded as 2 dichotomous variables: Chronic kidney disease (N18) and Hypertensive chronic kidney disease (I12).
17) **Chronic liver disease** encoded as 8 categorical variables: Alcoholic liver disease (K70), Hepatic failure, not elsewhere classified (K72), Chronic hepatitis, not elsewhere classified (K73), Fibrosis and cirrhosis of liver (K74), Fatty (change of) liver, not elsewhere classified (K76.0), Chronic passive congestion of liver (K76.1), Portal hypertension (K76.6), Other specified diseases of liver (K76.8).
18) **Stroke** encoded as the dichotomous variable Cerebral infarction (I63) .
19) **Dementia** encoded as 6 dichotomous variables: Vascular dementia (F01), Dementia in other diseases classified elsewhere (F02), Unspecified dementia (F03), Alzheimer's disease (G30), Frontotemporal dementia (G31.0), and Dementia with Lewy bodies (G31.83).
20) **Cancer and haematological cancer in particular** encoded as 2 dichotomous variables: Neoplasms (C00-D49) and Malignant neoplasms of lymphoid, hematopoietic and related tissue (C81-C96).
21) **Organ transplant** encoded as 2 dichotomous variables: Renal Transplantation Procedures and Liver Transplantation Procedures.

22) **Rheumatoid arthritis** encoded as 2 dichotomous variables: Rheumatoid arthritis with rheumatoid factor (M05) and Other rheumatoid arthritis (M06).
23) **Lupus** encoded as a dichotomous variable corresponding ICD-10 code (M32).
24) **Psoriasis** encoded as a dichotomous variable corresponding ICD-10 code (L40).
25) **Disorders involving an immune mechanism** encoded as a dichotomous variable "Certain disorders involving the immune mechanism" (D80-D89).

Each individual code was considered a confounding factor in and of itself so that matching was achieved for each of them individually. For instance, matching was achieved for each subcategory (and not just for the whole category) of chronic lower respiratory diseases. For variables representing diagnoses and socioeconomic deprivation, an individual was considered positive if the diagnostic was recorded at least once in their health record before the index event. For categorical variables representing measurements (i.e. BMI and blood pressures), all available measurements for all individuals were used and propensity score matching sought to define cohorts with similar numbers of measurements falling into each category.

**Definition of outcomes**

We included all outcomes whose rates were found to be significantly higher after COVID-19 in at least one of 4 large-scale EHR-based studies.[2,11–13] If an outcome was defined differently in two different studies, then it was defined by combining the codes used in both definitions to encompass all events that would have qualify as an outcome with either definitions.

All outcomes were defined as an event recorded in the patient's electronic health. Specifically, the following ICD-10 codes (with the ICD-10 labels in brackets), Current Procedural Terminology (CPT) for procedures, and VA Formulary for medications codes were used to define outcomes.

1. **Anosmia**: R43.0
2. **Anxiety disorder**: F40-F48
3. **Arrhythmia**: I47 ('Paroxysmal tachycardia'), I48 ('Atrial fibrillation and flutter'), I49 ('Other cardiac arrhythmias'), or R00.0 ('Tachycardia, unspecified')
4. **Cardiac failure**: I50 ('Heart failure'), I11.0 ('Hypertensive heart disease with heart failure'), or I13.0 ('Hypertensive heart and chronic kidney disease with heart failure and stage 1 through stage 4 chronic kidney disease, or unspecified chronic kidney disease')
5. **Cardiomyopathy**: I42 ('Cardiomyopathy'), I43 ('Cardiomyopathy in diseases classified elsewhere'), I25.5 ('Ischaemic cardiomyopathy'), B33.24 ('Viral cardiomyopathy'), or A36.81 ('Diphtheritic cardiomyopathy')
6. **Cerebral haemorrhage**: I60-I62
7. **Coronary disease**: I21 ('Acute myocardial infarction'), I22 ('Subsequent ST elevation and non-ST elevation myocardial infarction'), I20.0 ('Unstable angina'), I24 ('Other acute ischaemic heart disease'), or R57.0 ('Cardiogenic shock')
8. **Death**
9. **Gastro-oesophageal reflux disease (GORD)**: K21
10. **Hair loss**: L63 ('Alopecia areata'), L64 ('Androgenic alopecia'), L65 ('Other non-scarring hair loss'), or L66 ('Cicatricial alopecia')
11. **Hospitalisation**: 1013659 ('Hospital inpatient services') or 1013729 ('Critical care services')
12. **Hypercoagulopathy/Deep vein thrombosis (DVT) /Pulmonary embolism (PE)**: D68 ('Other coagulation defects'), I82 ('Other venous embolism and thrombosis'), or I26 ('Pulmonary embolism')
13. **Hyperlipidaemia**: E78.0-E78.5
14. **Hypertension**: I10-I16 ('Hypertensive diseases') or I27 ('Other pulmonary heart diseases')
15. **Hypoxaemia**: R09.02 ('Hypoxaemia') or J96.01 ('Acute respiratory failure with hypoxia')
16. **ICU admission**: 1013729 ('Critical care services')
17. **Interstitial lung disease**: J84 ('Other interstitial lung disease')
18. **Intubation/Ventilation**: 31500 ('Intubation, endotracheal, emergency procedure'), 5A1945Z ('Respiratory Ventilation, 24-96 Consecutive Hours'), 5A1955Z ('Respiratory Ventilation, Greater than 96 Consecutive Hours'), 5A1935Z ('Respiratory Ventilation, Less than 24 Consecutive Hours'), 0BH13EZ ('Insertion of Endotracheal Airway into Trachea, Percutaneous Approach'), 0BH17EZ ('Insertion of Endotracheal Airway into Trachea, Via Natural or Artificial Opening'), 0BH18EZ ('Insertion of Endotracheal Airway into Trachea, Via Natural or Artificial Opening Endoscopic'), 1022227 ('Extracorporeal membrane oxygenation (ECMO)/extracorporeal life support (ECLS)

provided by physician'), 39.65 ('Extracorporeal membrane oxygenation [ECMO]'), or 1015098 ('Ventilator Management')

19. **Ischaemic stroke**: I63 ('Cerebral infarction') or H34.1 ('Central retinal artery occlusion')
20. **Joint pain**: M25.5 ('Pain in joint')
21. **Kidney disease**: N18 ('Chronic kidney disease'), I12 ('Hypertensive chronic kidney disease'), I13 ('Hypertensive heart and chronic kidney disease'), N17 ('Acute kidney failure'), I12 ('Hypertensive chronic kidney disease'), I13 ('Hypertensive heart and chronic kidney disease'), or R88.0 ('Cloudy dialysis effluent')
22. **Liver disease**: K70-K76 ('Diseases of liver'), I85 ('Esophageal varices'), B18 ('Chronic viral hepatitis'), or I86.4 ('Gastric varices')
23. **Long COVID feature (any)**: Any of the following as defined in an EHR-based study of long-COVID features[11]
    a. **Abdominal symptoms**: R10 ('Abdominal and pelvic pain'), R19.4 ('Change in bowel habit'), or R19.7 ('Diarrhoea, unspecified').
    b. **Abnormal breathing**: R06 ('Abnormalities of breathing').
    c. **Anxiety/Depression**: F30-F39 ('Mood disorders') or F40-F48 ('Anxiety, dissociative, stress-related, somatoform and other nonpsychotic mental disorders'). F30-39 also encompasses diagnoses other than depression (e.g. mania), but these comprise only a small fraction of cases and so we use Anxiety/Depression for convenience.
    d. **Chest/Throat pain**: R07 ('Pain in throat and chest').
    e. **Cognitive symptoms**: R40 ('Somnolence, stupor and coma'), R41 ('Other symptoms and signs involving cognitive functions and awareness'), R48 ('Dyslexia and other symbolic dysfunction'), G93.40 ('Encephalopathy, unspecified'), G31.84 ('Mild cognitive impairment'), G30 ('Alzheimer's disease'), G31.0 ('Frontotemporal dementia'), G31.83 ('Dementia with Lewy bodies'), F01 ('Vascular dementia'), F02 ('Dementia in other disease classified elsewhere'), F03 ('Unspecified dementia'), F05 ('Delirium due to known physiological condition'), or F06.8 ('Other specified mental disorders due to known physiological condition'). These terms were used to capture the range of diagnostic codes that patients presenting with 'brain fog' might receive.
    f. **Fatigue**: G93.3 ('Postviral fatigue syndrome') or R53 ('Malaise and fatigue').
    g. **Headache**: R51 ('Headache'), G43 ('Migraine'), or G44 ('Other headache syndrome').
    h. **Myalgia**: M79.1 ('Myalgia') or M60 ('Myositis'). Inclusion of the latter category was intended to capture those patients who received a specific diagnosis when presenting with myalgia.
    i. **Other pain**: G89 ('Pain not elsewhere specified') or R52 ('Pain, unspecified').
24. **Mood disorder**: F30-F39
25. **Myocarditis**: I40 ('Acute myocarditis'), B33.22 ('Viral myocarditis'), or I51.4 ('Myocarditis, unspecified')
26. **Myoneural junction/muscle disease**: G70-G73
27. **Nerve/nerve root/plexus disorder**: G50-G59
28. **Obesity**: E66 ('Overweight and obesity') or BMI recorded $\geq 30$ kg/m$^2$
29. **Oxygen requirement**: RE600-7806 ('Oxygen'), Oxygen saturation measures $\leq 92\%$, 5A09 ('Physiological Systems / Assistance / Respiratory'), Z99.81 ('Dependence on supplemental oxygen'), 5A05121 ('Extracorporeal Hyperbaric Oxygenation, Intermittent'), 5A0512C ('Extracorporeal Supersaturated Oxygenation, Intermittent'), 5A05221 (Extracorporeal Hyperbaric Oxygenation, Continuous'), 5A0522C ('Extracorporeal Supersaturated Oxygenation, Continuous')
30. **Peripheral neuropathy**: G62.9 ('Polyneuropathy, unspecified'), or G63 ('Polyneuropathy in diseases classified elsewhere')
31. **Psychotic disorder**: F20-F29
32. **Respiratory failure**: J96 ('Respiratory failure, not elsewhere classified')
33. **Seizures**: G40 ('Epilepsy and recurrent seizures')
34. **Sleep disorders**: G47
35. **Type 2 diabetes mellitus**: E11
36. **Urticaria**: L50

**Details on statistical analyses**

*Implementation details of propensity score matching*
In propensity score matching, the propensity score was calculated using a logistic regression (implemented by the function LogisticRegression of the scikit-learn package in Python 3.7) including each of the covariates mentioned above. To eliminate the influence of ordering of records, the order of the records in the covariate matrix were randomised before matching.

*Testing proportional hazards*
The assumption that the hazards were proportional when accounting for the two phases was tested using the generalized Schoenfeld approach[14] implemented in the cox.zph function of the survival package (version 3.2.3) in R. If the proportional hazard assumption was found to be violated (i.e. statistical evidence from a score test indicating a non-zero slope in the scaled Schoenfeld residuals over time), then the time-varying HR was assessed using natural cubic splines (in log-time) to the log-cumulative hazard[15]. This was achieved using the generalized survival models of the rstpm2 package (version 1.5.1) in R[16]. As recommended by Royston and Parmar[15], splines with 1, 2, and 3 degrees of freedom were estimated for both the baseline log-cumulative hazard and its cohort dependency and the number of degrees of freedom leading to the lowest Akaike Information Criterion (AIC) was selected. This was achieved on a per-comparison basis so that more complex time dependency (i.e. higher number of degrees of freedom) could be selected for a specific comparison if there was enough evidence in the data to support such complexity.

*Accounting for competing risks*
Because none of the outcomes can occur once a patient has died, death is a competing risk for all of the outcomes that we considered. Various approaches exist to account for competing risks in time-to-event analysis. A simple and often used approach is to include the competing risk in a composite endpoint.[17] This was the approach used here. Other, more elaborate, approaches seek to estimate the cause-specific hazard.[18] However, these were not achievable within TriNetX nor on the data returned by queries to TriNetX.

*Assessing moderation of associations*
The Cox model used in the primary analysis reads:

$$\lambda(t|\text{Vaccinated}) = \lambda_0(t)\exp(\beta_0 + \beta_V \text{Vaccinated}),$$

where Vaccinated=1 if the individual has been vaccinated, and 0 otherwise. The HR is then obtained as:

$$HR = \frac{\lambda(t|\text{Vaccinated=1})}{\lambda(t|\text{Vaccinated=0})} = \exp(\beta_V).$$

This is the standard approach to calculating HRs. To assess whether associations between vaccination status and COVID-19 outcomes are moderated by a variable *X*, we add an interaction term as follows:

$$\lambda(t|\text{Vaccinated}, X) = \lambda_0(t)\exp(\beta_0 + \beta_V \text{Vaccinated} + \beta_X X + \beta_{XV}\, X \times \text{Vaccinated}).$$

If X is a dummy variable (i.e. it only takes value 1 or 0), then the HR related to vaccination status for *X*=0 is:

$$HR_{X=0} = \frac{\lambda(t|\text{Vaccinated=1}, X\text{=0})}{\lambda(t|\text{Vaccinated=0}, X\text{=0})} = \exp(\beta_V),$$

and the HR related to vaccination status for *X*=1 is:

$$HR_{X=1} = \frac{\lambda(t|\text{Vaccinated=1}, X\text{=1})}{\lambda(t|\text{Vaccinated=0}, X\text{=1})} = \exp(\beta_V + \beta_{XV}) = \exp(\beta_V)\exp(\beta_{XV}).$$

Therefore, the ratio between the two (which are presented for each outcome in Table S10 and S13) is:

$$\frac{HR_{X=1}}{HR_{X=0}} = \exp(\beta_{XV}).$$

Testing whether the ratio of HR is equal to 1 thus amounts to testing whether $\beta_{XV}$ is equal to zero, and this is tested as part of the estimation of the Cox model with coxph function from the Survival package (version 3.2.3).

### Supplementary figures

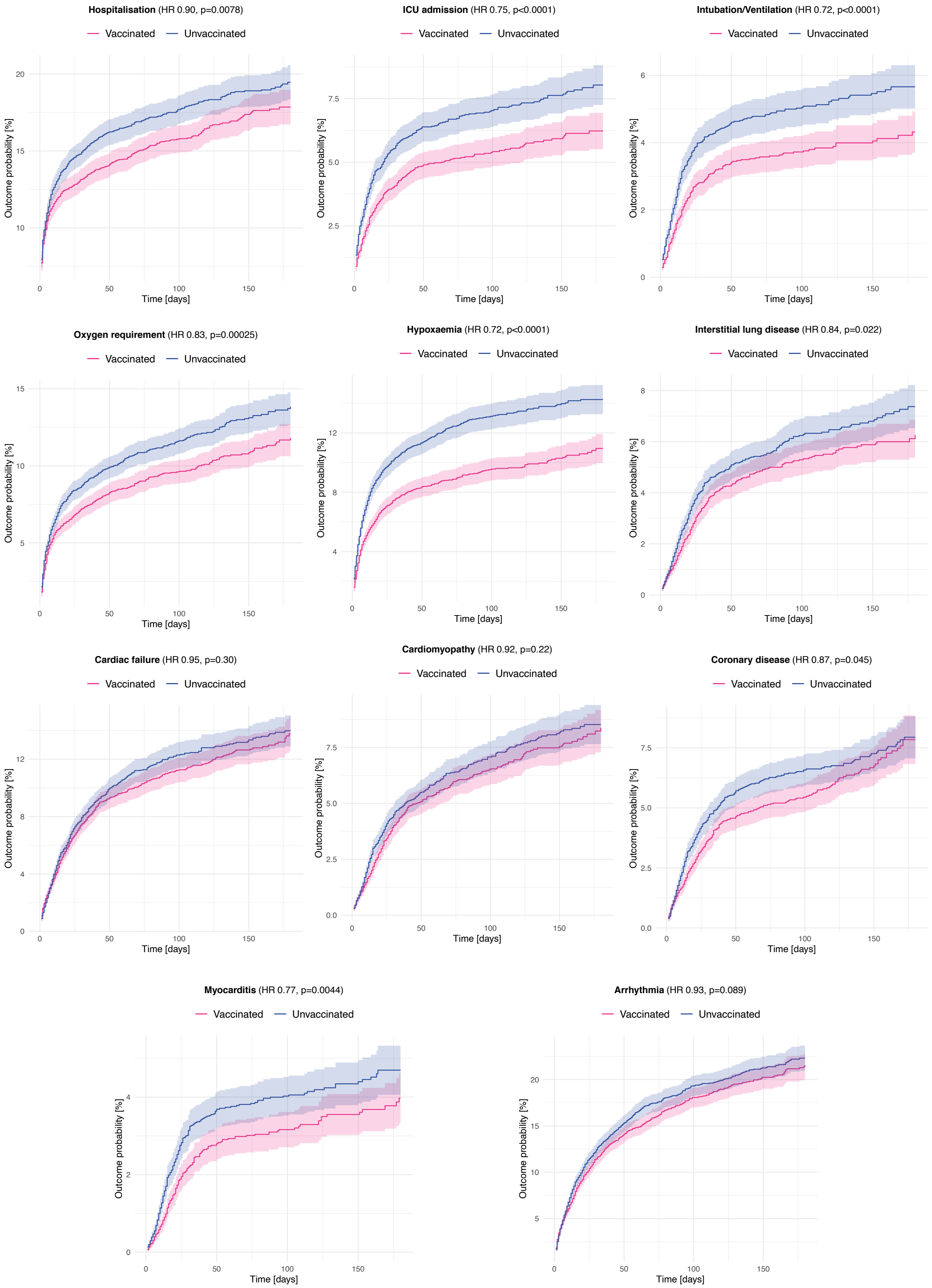

**Fig. S1** – Same figure as Fig. 2 in the main manuscript but representing Kaplan-Meier curves for additional outcomes (broadly service use, respiratory and cardiac outcomes).

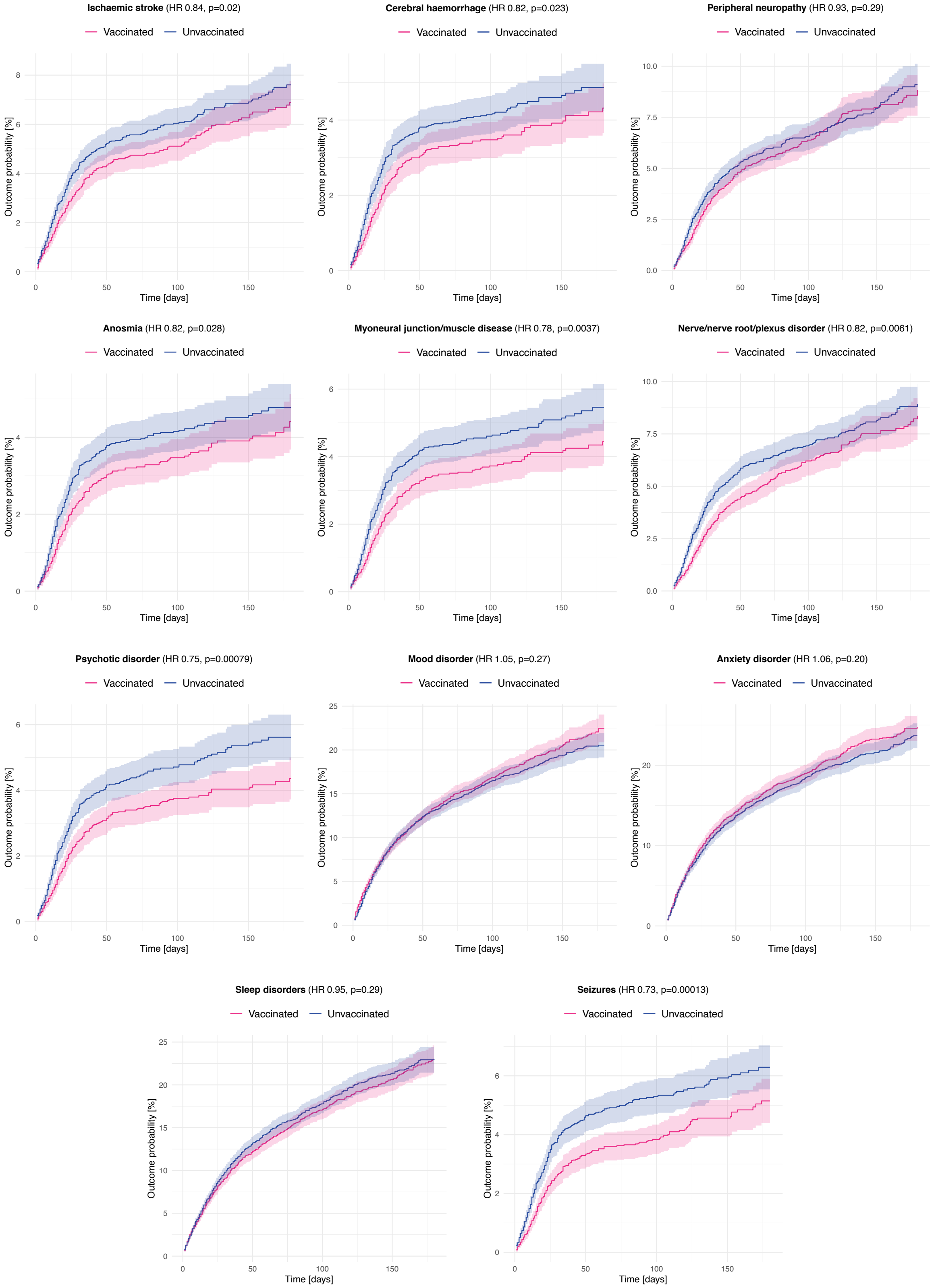

**Fig. S2** – Same figure as Fig. 2 in the main manuscript but representing Kaplan-Meier curves for additional outcomes (broadly the neuropsychiatric outcomes).

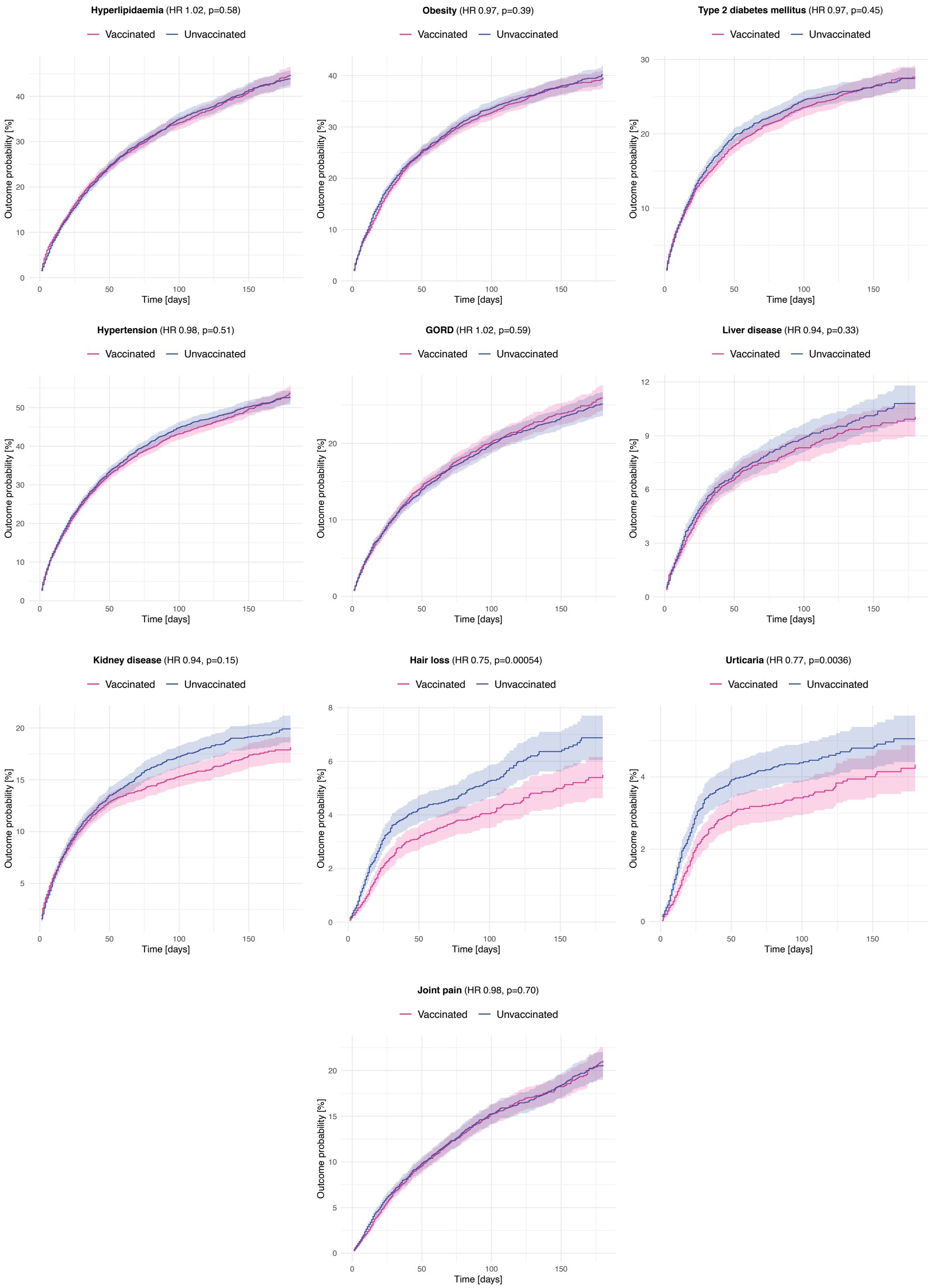

**Fig. S3** – Same figure as Fig. 2 in the main manuscript but representing Kaplan-Meier curves for additional outcomes (broadly the outcomes affecting other body systems not represented in Fig. S2-S3).

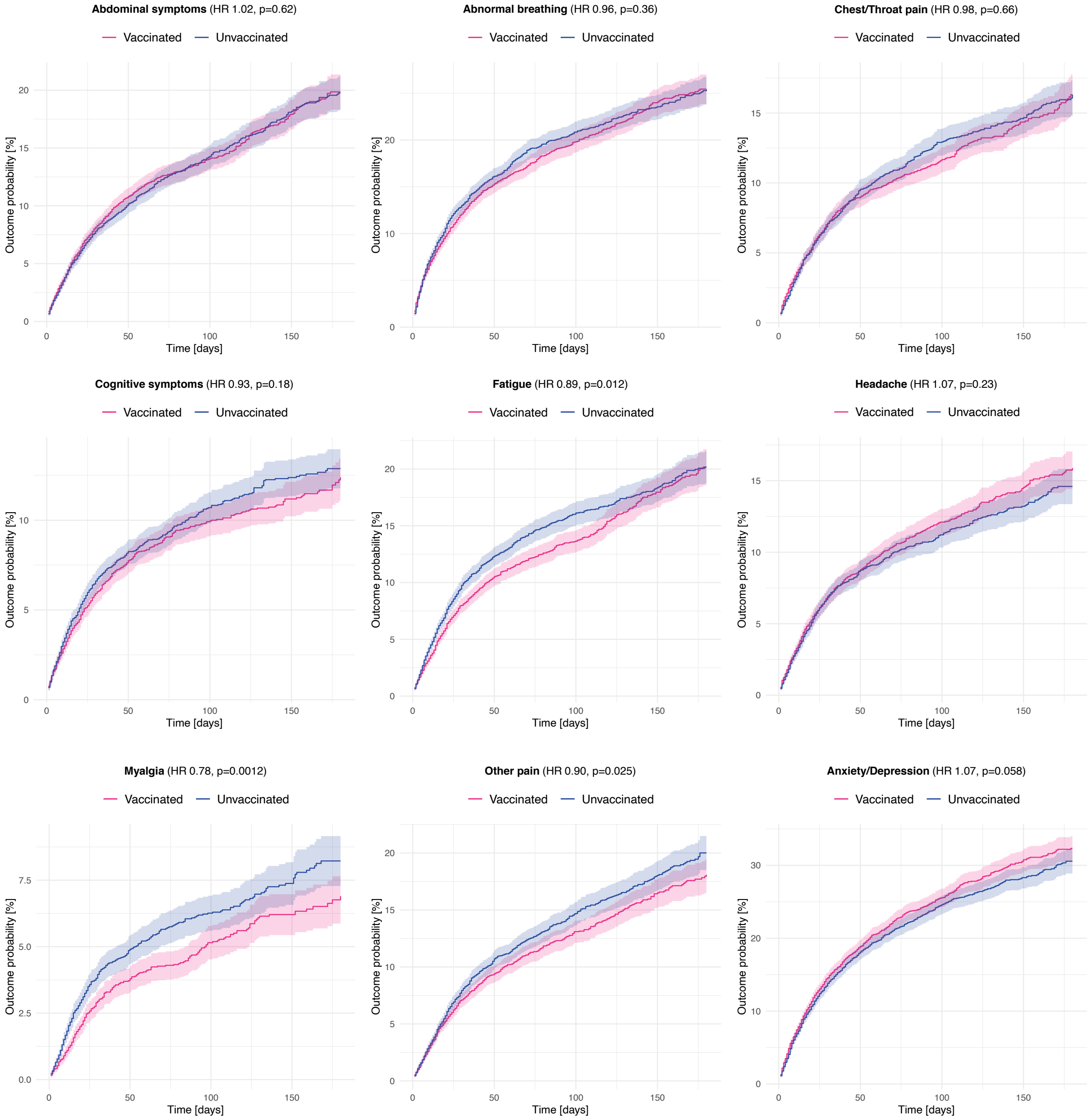

**Fig. S4** – Same figure as Fig. 2 in the main manuscript but representing Kaplan-Meier curves for additional the outcomes termed "long-COVID features".

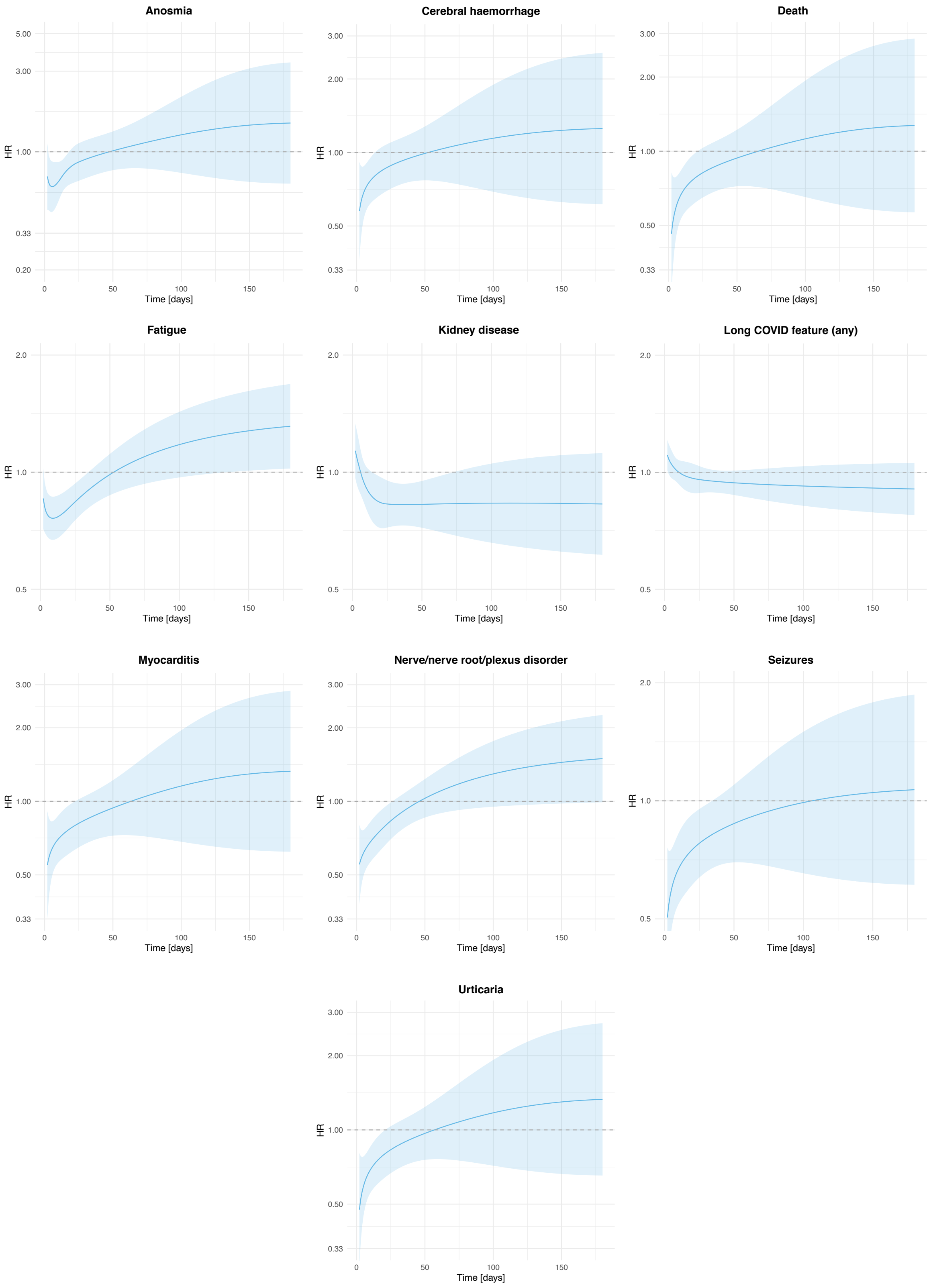

**Fig. S5** – Time-varying hazard ratios for the outcomes for which there was evidence of non-proportionality of hazards (as per Table S7).

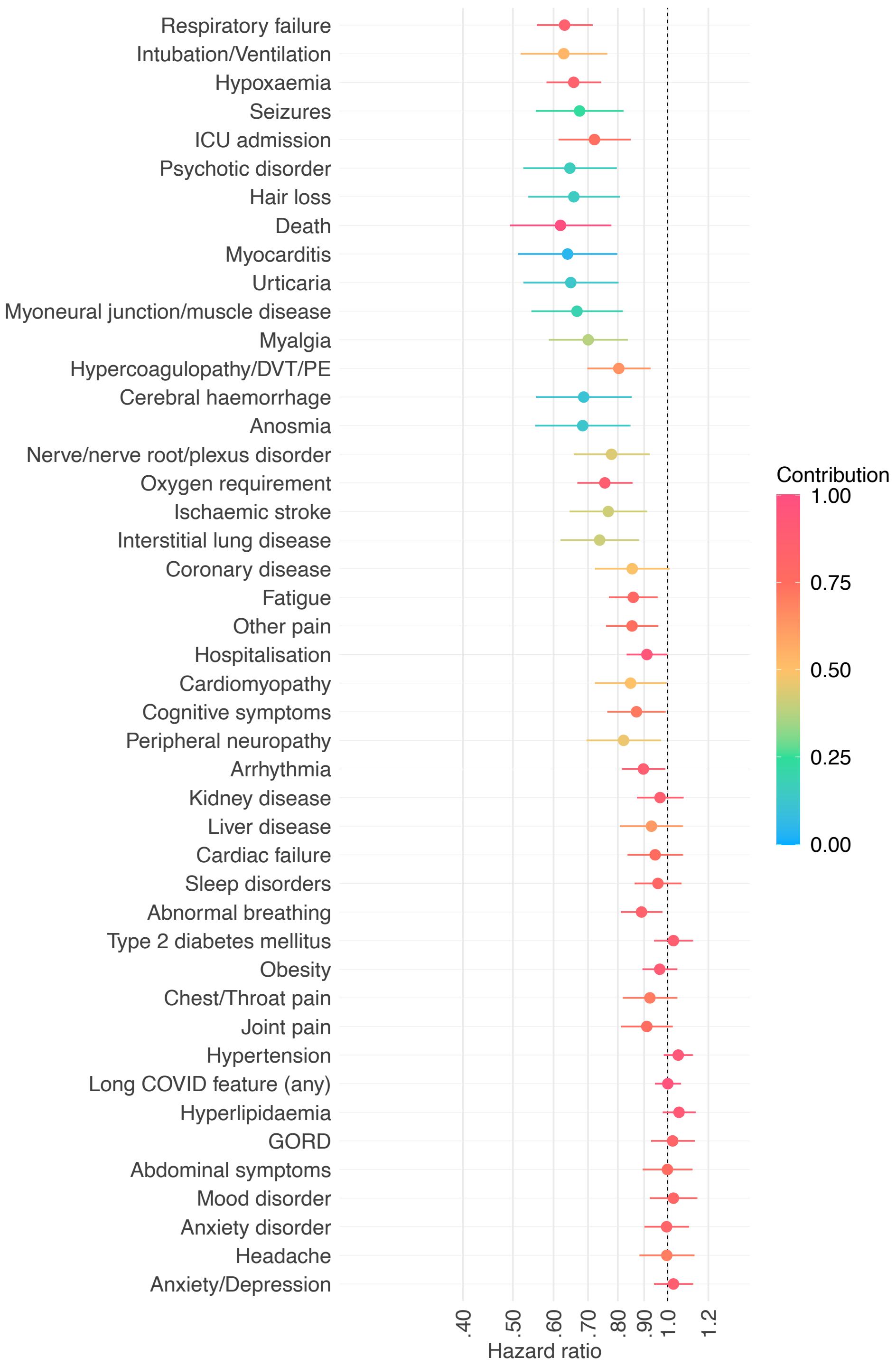

**Fig. S6** – Same figure as Fig. 3 in the main manuscript for the comparison of individuals having received two doses of the vaccine vs. unvaccinated individuals but representing hazard ratios for *all* outcomes. HR lower than 1 indicate outcomes less common among vaccinated individuals. Horizontal bars represent 95% confidence intervals. Each outcome is a composite outcome with death as a component to address competing risks. The contribution of the outcome of interest to the overall incidence of the composite endpoint is encoded by the colour.

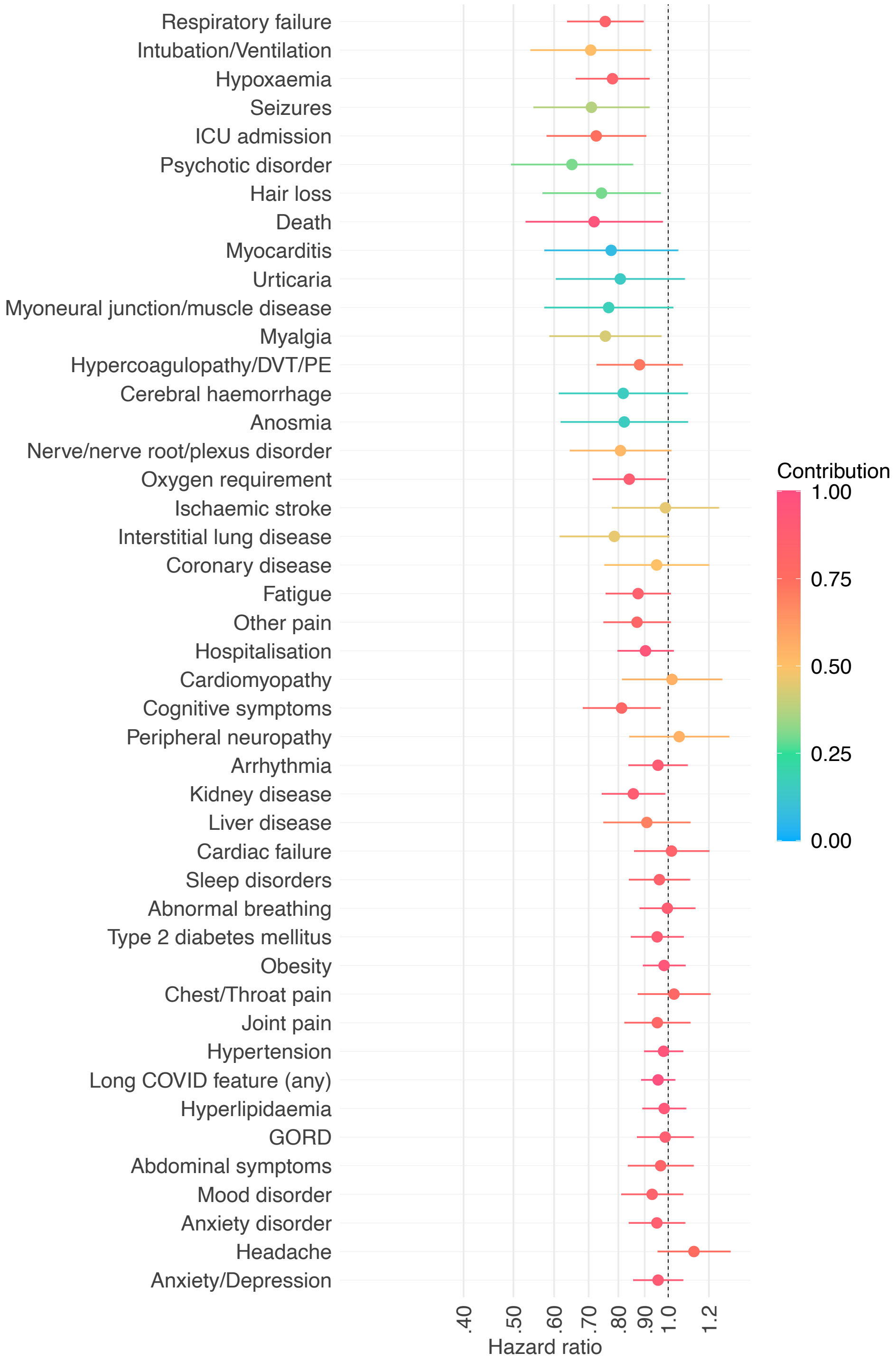

**Fig. S7** – Same figure as Fig. 3 in the main manuscript for the comparison of individuals having received only one dose of the vaccine at the time of SARS-CoV-2 infection vs. unvaccinated individuals but representing hazard ratios for *all* outcomes. HR lower than 1 indicate outcomes less common among vaccinated individuals. Horizontal bars represent 95% confidence intervals. Each outcome is a composite outcome with death as a component to address competing risks. The contribution of the outcome of interest to the overall incidence of the composite endpoint is encoded by the colour.

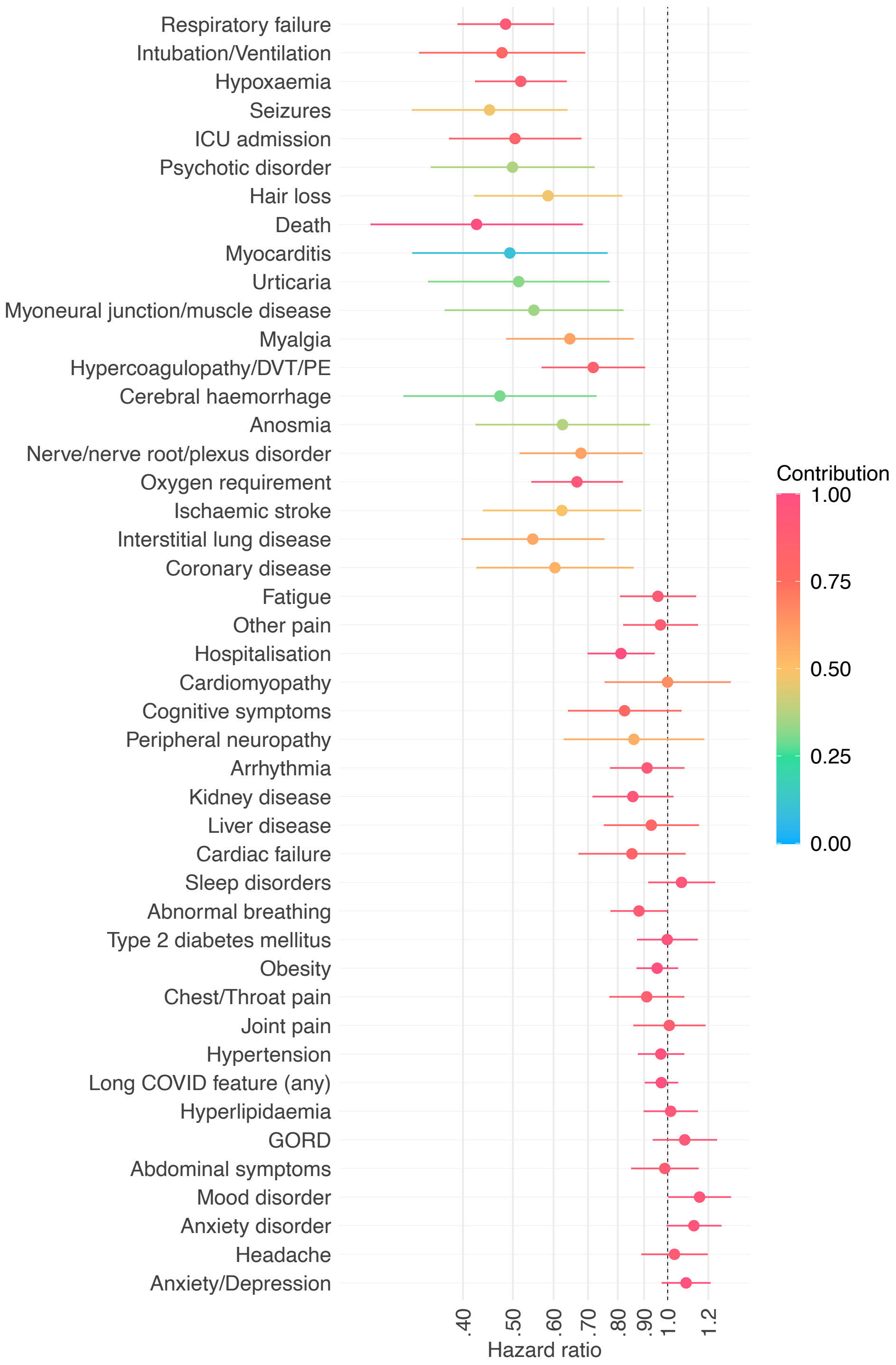

**Fig. S8** – Same figure as Fig. 3 in the main manuscript for the comparison of vaccinated vs. unvaccinated individuals among people < 60 years-old, and representing hazard ratios for *all* outcomes. HR lower than 1 indicate outcomes less common among vaccinated individuals. Horizontal bars represent 95% confidence intervals. Each outcome is a composite outcome with death as a component to address competing risks. The contribution of the outcome of interest to the overall incidence of the composite endpoint is encoded by the colour.

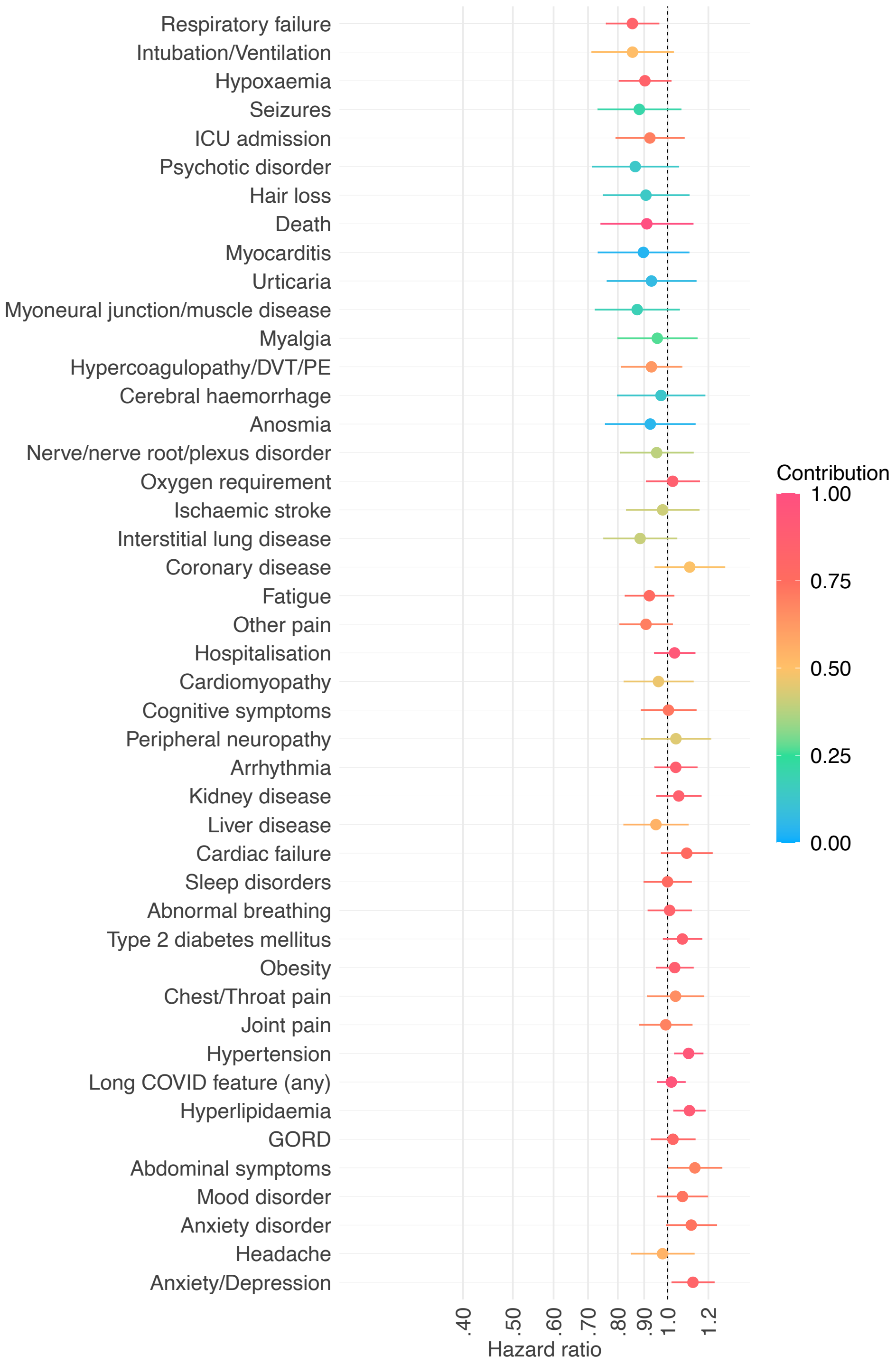

**Fig. S9** – Same figure as Fig. 3 in the main manuscript for the comparison of vaccinated vs. unvaccinated individuals among people ≥ 60 years-old, and representing hazard ratios for *all* outcomes. HR lower than 1 indicate outcomes less common among vaccinated individuals. Horizontal bars represent 95% confidence intervals. Each outcome is a composite outcome with death as a component to address competing risks. The contribution of the outcome of interest to the overall incidence of the composite endpoint is encoded by the colour.

#### Supplementary tables

**Table S1** – Characteristics of the vaccinated and unvaccinated cohorts, before and after propensity score matching. SMD = Standardised Mean Difference.

| | Vaccinated (unmatched) | Unvaccinated (unmatched) | Vaccinated (matched) | Unvaccinated (matched) | SMD |
|---|---|---|---|---|---|
| Number | 10024 | 83957 | 9479 | 9479 | - |
| DEMOGRAPHICS | | | | | |
| Age; mean (SD) | 57.0 (17.9) | 51.9 (23.1) | 56.5 (18.0) | 57.6 (20.6) | 0.06 |
| Sex; n (%) | | | | | |
| Female | 5950 (59.4) | 48968 (58.3) | 5676 (59.9) | 5761 (60.8) | 0.02 |
| Male | 4072 (40.6) | 34981 (41.7) | 3801 (40.1) | 3716 (39.2) | 0.02 |
| Other | 20 (0.2) | 40 (0.0) | 20 (0.2) | 10 (0.1) | 0.03 |
| Race; n (%) | | | | | |
| White | 7208 (71.9) | 57310 (68.3) | 6783 (71.6) | 6873 (72.5) | 0.02 |
| Black or African American | 1584 (15.8) | 16296 (19.4) | 1540 (16.2) | 1514 (16.0) | 0.007 |
| Asian | 368 (3.7) | 1992 (2.4) | 317 (3.3) | 285 (3.0) | 0.02 |
| American Indian or Alaska Native | 61 (0.6) | 345 (0.4) | 57 (0.6) | 57 (0.6) | 0 |
| Native Hawaiian or Other Pacific Islander | 42 (0.4) | 171 (0.2) | 40 (0.4) | 30 (0.3) | 0.02 |
| Unknown | 802 (8.0) | 7843 (9.3) | 783 (8.3) | 756 (8.0) | 0.01 |
| Ethnicity; n (%) | | | | | |
| Hispanic or Latino | 1097 (10.9) | 7539 (9.0) | 1015 (10.7) | 974 (10.3) | 0.01 |
| Not Hispanic or Latino | 7199 (71.8) | 63015 (75.1) | 6780 (71.5) | 6793 (71.7) | 0.003 |
| Unknown | 1728 (17.2) | 13403 (16.0) | 1684 (17.8) | 1712 (18.1) | 0.008 |
| Problems related to housing and economic circumstances; n (%) | 135 (1.3) | 1735 (2.1) | 133 (1.4) | 146 (1.5) | 0.01 |
| COMORBIDITIES | | | | | |
| BMI; mean (SD) | 30.0 (7.4) | 29.7 (8.3) | 30.1 (7.4) | 30.2 (7.5) | 0.03 |
| Overweight and obesity; n (%) | 3175 (31.7) | 30363 (36.2) | 3086 (32.6) | 3263 (34.4) | 0.04 |
| Blood pressure; mean (SD) | | | | | |
| Diastolic | 74.7 (11.5) | 73.0 (12.1) | 74.6 (11.5) | 73.1 (11.6) | 0.1 |
| Systolic | 127.6 (19.0) | 124.2 (19.6) | 127.5 (18.8) | 126.0 (19.4) | 0.08 |
| Hypertensive diseases; n (%) | 5227 (52.1) | 46211 (55.0) | 5060 (53.4) | 5197 (54.8) | 0.03 |
| Diabetes mellitus; n (%) | | | | | |
| Type 1 diabetes mellitus | 436 (4.3) | 4825 (5.7) | 417 (4.4) | 470 (5.0) | 0.03 |
| Type 2 diabetes mellitus | 2508 (25.0) | 24073 (28.7) | 2426 (25.6) | 2551 (26.9) | 0.03 |
| Chronic lower respiratory diseases; n (%) | | | | | |
| Asthma | 1526 (15.2) | 17665 (21.0) | 1502 (15.8) | 1491 (15.7) | 0.003 |
| Bronchiectasis | 217 (2.2) | 1711 (2.0) | 211 (2.2) | 233 (2.5) | 0.02 |
| Bronchitis, not specified as acute or chronic | 887 (8.8) | 10121 (12.1) | 870 (9.2) | 867 (9.1) | 0.001 |
| Emphysema | 398 (4.0) | 3981 (4.7) | 387 (4.1) | 413 (4.4) | 0.01 |
| Other chronic obstructive pulmonary disease | 884 (8.8) | 10493 (12.5) | 871 (9.2) | 958 (10.1) | 0.03 |
| Simple and mucopurulent chronic bronchitis | 133 (1.3) | 1210 (1.4) | 129 (1.4) | 147 (1.6) | 0.02 |
| Unspecified chronic bronchitis | 126 (1.3) | 1433 (1.7) | 125 (1.3) | 126 (1.3) | 0.0009 |
| Nicotine dependence; n (%) | 929 (9.3) | 11509 (13.7) | 917 (9.7) | 935 (9.9) | 0.006 |
| Psychiatric comorbidities; n (%) | | | | | |
| Anxiety disorders | 3187 (31.8) | 31966 (38.1) | 3098 (32.7) | 3159 (33.3) | 0.01 |

| | | | | | |
|---|---|---|---|---|---|
| Substance misuse | 1351 (13.5) | 16299 (19.4) | 1334 (14.1) | 1373 (14.5) | 0.01 |
| Mood disorders | 2453 (24.5) | 27245 (32.5) | 2408 (25.4) | 2458 (25.9) | 0.01 |
| Psychotic disorders | 176 (1.8) | 2827 (3.4) | 175 (1.8) | 176 (1.9) | 0.0008 |
| Heart disease; n (%) | | | | | |
| Ischemic heart diseases | 2074 (20.7) | 18880 (22.5) | 2013 (21.2) | 2114 (22.3) | 0.03 |
| Other forms of heart disease | 3573 (35.6) | 34359 (40.9) | 3485 (36.8) | 3607 (38.1) | 0.03 |
| Chronic kidney diseases; n (%) | | | | | |
| Chronic kidney disease (CKD) | 1501 (15.0) | 15137 (18.0) | 1471 (15.5) | 1561 (16.5) | 0.03 |
| Hypertensive chronic kidney disease | 783 (7.8) | 8795 (10.5) | 761 (8.0) | 812 (8.6) | 0.02 |
| Chronic liver diseases; n (%) | | | | | |
| Alcoholic liver disease | 88 (0.9) | 848 (1.0) | 84 (0.9) | 86 (0.9) | 0.002 |
| Chronic hepatitis, not elsewhere classified | 57 (0.6) | 340 (0.4) | 53 (0.6) | 53 (0.6) | 0 |
| Chronic passive congestion of liver | 140 (1.4) | 1419 (1.7) | 136 (1.4) | 157 (1.7) | 0.02 |
| Fatty (change of) liver, not elsewhere classified | 727 (7.3) | 6530 (7.8) | 705 (7.4) | 730 (7.7) | 0.01 |
| Fibrosis and cirrhosis of liver | 251 (2.5) | 2269 (2.7) | 240 (2.5) | 266 (2.8) | 0.02 |
| Hepatic failure, not elsewhere classified | 141 (1.4) | 1248 (1.5) | 132 (1.4) | 128 (1.4) | 0.004 |
| Portal hypertension | 118 (1.2) | 1047 (1.2) | 111 (1.2) | 133 (1.4) | 0.02 |
| Cerebral infarction; n (%) | 497 (5.0) | 5384 (6.4) | 487 (5.1) | 508 (5.4) | 0.01 |
| Dementia; n (%) | | | | | |
| Alzheimer's disease | 70 (0.7) | 1181 (1.4) | 68 (0.7) | 71 (0.7) | 0.004 |
| Dementia in other diseases classified elsewhere | 97 (1.0) | 1555 (1.9) | 95 (1.0) | 95 (1.0) | 0 |
| Dementia with Lewy bodies | 40 (0.4) | 149 (0.2) | 40 (0.4) | 40 (0.4) | 0 |
| Frontotemporal dementia | 20 (0.2) | 89 (0.1) | 20 (0.2) | 30 (0.3) | 0.02 |
| Unspecified dementia | 179 (1.8) | 3013 (3.6) | 178 (1.9) | 183 (1.9) | 0.004 |
| Vascular dementia | 60 (0.6) | 963 (1.1) | 59 (0.6) | 66 (0.7) | 0.009 |
| Neoplasms; n (%) | | | | | |
| Malignant neoplasms of lymphoid, hematopoietic and related tissue | 392 (3.9) | 2718 (3.2) | 387 (4.1) | 397 (4.2) | 0.005 |
| Neoplasms (any) | 4184 (41.7) | 34046 (40.6) | 4038 (42.6) | 4208 (44.4) | 0.04 |
| Organ transplant; n (%) | | | | | |
| Liver Transplantation Procedures | 50 (0.5) | 140 (0.2) | 47 (0.5) | 43 (0.5) | 0.006 |
| Renal Transplantation Procedures | 124 (1.2) | 704 (0.8) | 114 (1.2) | 113 (1.2) | 0.001 |
| Psoriasis; n (%) | 246 (2.5) | 2142 (2.6) | 242 (2.6) | 254 (2.7) | 0.008 |
| Rheumatoid arthritis; n (%) | | | | | |
| Other rheumatoid arthritis | 348 (3.5) | 3069 (3.7) | 341 (3.6) | 349 (3.7) | 0.005 |
| Rheumatoid arthritis with rheumatoid factor | 123 (1.2) | 1022 (1.2) | 118 (1.2) | 130 (1.4) | 0.01 |
| Systemic lupus erythematosus (SLE); n (%) | 125 (1.2) | 1129 (1.3) | 122 (1.3) | 120 (1.3) | 0.002 |
| Disorders involving the immune mechanism; n (%) | 810 (8.1) | 5844 (7.0) | 778 (8.2) | 771 (8.1) | 0.003 |

**Table S2** – Characteristics of the vaccinated (1 dose only) and unvaccinated cohorts after propensity score matching. SMD = Standardised Mean Difference.

| | Vaccinated | Unvaccinated | SMD |
|---|---|---|---|
| Number | 2996 | 2996 | - |
| DEMOGRAPHICS | | | |
| Age; mean (SD) | 58.2 (17.2) | 58.6 (19.7) | 0.02 |
| Sex; n (%) | | | |
| Female | 1808 (60.3) | 1837 (61.3) | 0.02 |
| Male | 1181 (39.4) | 1150 (38.4) | 0.02 |
| Other | 30 (1.0) | 20 (0.7) | 0.04 |
| Race; n (%) | | | |
| White | 2043 (68.2) | 2047 (68.3) | 0.003 |
| Black or African American | 534 (17.8) | 522 (17.4) | 0.01 |
| Asian | 123 (4.1) | 129 (4.3) | 0.01 |
| American Indian or Alaska Native | 70 (2.3) | 60 (2.0) | 0.02 |
| Native Hawaiian or Other Pacific Islander | 30 (1.0) | 20 (0.7) | 0.04 |
| Unknown | 281 (9.4) | 288 (9.6) | 0.008 |
| Ethnicity; n (%) | | | |
| Hispanic or Latino | 297 (9.9) | 296 (9.9) | 0.001 |
| Not Hispanic or Latino | 2246 (75.0) | 2267 (75.7) | 0.02 |
| Unknown | 453 (15.1) | 435 (14.5) | 0.02 |
| Problems related to housing and economic circumstances; n (%) | 83 (2.8) | 88 (2.9) | 0.01 |
| COMORBIDITIES | | | |
| BMI; mean (SD) | 30.9 (7.5) | 30.3 (7.5) | 0.08 |
| Overweight and obesity; n (%) | 1000 (33.4) | 956 (31.9) | 0.03 |
| Blood pressure; mean (SD) | | | |
| Diastolic | 74.3 (11.6) | 73.0 (12.2) | 0.1 |
| Systolic | 127.0 (18.8) | 126.1 (19.6) | 0.05 |
| Hypertensive diseases; n (%) | 1604 (53.5) | 1639 (54.7) | 0.02 |
| Diabetes mellitus; n (%) | | | |
| Type 1 diabetes mellitus | 134 (4.5) | 124 (4.1) | 0.02 |
| Type 2 diabetes mellitus | 803 (26.8) | 768 (25.6) | 0.03 |
| Chronic lower respiratory diseases; n (%) | | | |
| Asthma | 506 (16.9) | 539 (18.0) | 0.03 |
| Bronchiectasis | 84 (2.8) | 82 (2.7) | 0.004 |
| Bronchitis, not specified as acute or chronic | 243 (8.1) | 241 (8.0) | 0.002 |
| Emphysema | 128 (4.3) | 140 (4.7) | 0.02 |
| Other chronic obstructive pulmonary disease | 278 (9.3) | 289 (9.6) | 0.01 |
| Simple and mucopurulent chronic bronchitis | 80 (2.7) | 80 (2.7) | 0 |
| Unspecified chronic bronchitis | 80 (2.7) | 82 (2.7) | 0.004 |
| Nicotine dependence; n (%) | 319 (10.6) | 331 (11.0) | 0.01 |
| Psychiatric comorbidities; n (%) | | | |
| Anxiety disorders | 1047 (34.9) | 1039 (34.7) | 0.006 |
| Substance misuse | 455 (15.2) | 457 (15.3) | 0.002 |
| Mood disorders | 811 (27.1) | 815 (27.2) | 0.003 |

| | | | |
|---|---|---|---|
| Psychotic disorders | 86 (2.9) | 101 (3.4) | 0.03 |
| Heart disease; n (%) | | | |
| Ischemic heart diseases | 606 (20.2) | 627 (20.9) | 0.02 |
| Other forms of heart disease | 1119 (37.3) | 1132 (37.8) | 0.009 |
| Chronic kidney diseases; n (%) | | | |
| Chronic kidney disease (CKD) | 477 (15.9) | 486 (16.2) | 0.008 |
| Hypertensive chronic kidney disease | 245 (8.2) | 249 (8.3) | 0.005 |
| Chronic liver diseases; n (%) | | | |
| Alcoholic liver disease | 80 (2.7) | 70 (2.3) | 0.02 |
| Chronic hepatitis, not elsewhere classified | 60 (2.0) | 70 (2.3) | 0.02 |
| Chronic passive congestion of liver | 81 (2.7) | 80 (2.7) | 0.002 |
| Fatty (change of) liver, not elsewhere classified | 230 (7.7) | 208 (6.9) | 0.03 |
| Fibrosis and cirrhosis of liver | 94 (3.1) | 94 (3.1) | 0 |
| Hepatic failure, not elsewhere classified | 85 (2.8) | 84 (2.8) | 0.002 |
| Portal hypertension | 81 (2.7) | 81 (2.7) | 0 |
| Cerebral infarction; n (%) | 147 (4.9) | 145 (4.8) | 0.003 |
| Dementia; n (%) | | | |
| Alzheimer's disease | 80 (2.7) | 80 (2.7) | 0 |
| Dementia in other diseases classified elsewhere | 70 (2.3) | 63 (2.1) | 0.02 |
| Dementia with Lewy bodies | 30 (1.0) | 30 (1.0) | 0 |
| Frontotemporal dementia | 10 (0.3) | 20 (0.7) | 0.05 |
| Unspecified dementia | 80 (2.7) | 87 (2.9) | 0.01 |
| Vascular dementia | 70 (2.3) | 70 (2.3) | 0 |
| Neoplasms; n (%) | | | |
| Malignant neoplasms of lymphoid, hematopoietic and related tissue | 110 (3.7) | 126 (4.2) | 0.03 |
| Neoplasms (any) | 1225 (40.9) | 1229 (41.0) | 0.003 |
| Organ transplant; n (%) | | | |
| Liver Transplantation Procedures | 70 (2.3) | 70 (2.3) | 0 |
| Renal Transplantation Procedures | 80 (2.7) | 73 (2.4) | 0.01 |
| Psoriasis; n (%) | 103 (3.4) | 86 (2.9) | 0.03 |
| Rheumatoid arthritis; n (%) | | | |
| Other rheumatoid arthritis | 123 (4.1) | 115 (3.8) | 0.01 |
| Rheumatoid arthritis with rheumatoid factor | 70 (2.3) | 70 (2.3) | 0 |
| Systemic lupus erythematosus (SLE); n (%) | 87 (2.9) | 83 (2.8) | 0.008 |
| Disorders involving the immune mechanism; n (%) | 277 (9.2) | 292 (9.7) | 0.02 |

**Table S3** – Characteristics of the vaccinated (2 doses only) and unvaccinated cohorts after propensity score matching. SMD = Standardised Mean Difference.

| | **Vaccinated** | **Unvaccinated** | **SMD** |
|---|---|---|---|
| Number | 6957 | 6957 | - |
| DEMOGRAPHICS | | | |
| Age; mean (SD) | 57.2 (17.8) | 58.3 (20.6) | 0.06 |
| Sex; n (%) | | | |
| Female | 4071 (58.5) | 4133 (59.4) | 0.02 |
| Male | 2884 (41.5) | 2822 (40.6) | 0.02 |
| Other | 20 (0.3) | 20 (0.3) | 0 |
| Race; n (%) | | | |
| White | 5032 (72.3) | 5088 (73.1) | 0.02 |
| Black or African American | 1082 (15.6) | 1055 (15.2) | 0.01 |
| Asian | 248 (3.6) | 250 (3.6) | 0.002 |
| American Indian or Alaska Native | 53 (0.8) | 43 (0.6) | 0.02 |
| Native Hawaiian or Other Pacific Islander | 30 (0.4) | 30 (0.4) | 0 |
| Unknown | 552 (7.9) | 522 (7.5) | 0.02 |
| Ethnicity; n (%) | | | |
| Hispanic or Latino | 775 (11.1) | 775 (11.1) | 0 |
| Not Hispanic or Latino | 5030 (72.3) | 4994 (71.8) | 0.01 |
| Unknown | 1152 (16.6) | 1188 (17.1) | 0.01 |
| Problems related to housing and economic circumstances; n (%) | 97 (1.4) | 102 (1.5) | 0.006 |
| COMORBIDITIES | | | |
| BMI; mean (SD) | 29.9 (7.2) | 29.9 (7.5) | 0.002 |
| Overweight and obesity; n (%) | 2255 (32.4) | 2321 (33.4) | 0.02 |
| Blood pressure; mean (SD) | | | |
| Diastolic | 74.7 (11.5) | 73.3 (11.2) | 0.1 |
| Systolic | 127.9 (18.8) | 126.5 (19.0) | 0.08 |
| Hypertensive diseases; n (%) | 3754 (54.0) | 3852 (55.4) | 0.03 |
| Diabetes mellitus; n (%) | | | |
| Type 1 diabetes mellitus | 306 (4.4) | 303 (4.4) | 0.002 |
| Type 2 diabetes mellitus | 1771 (25.5) | 1787 (25.7) | 0.005 |
| Chronic lower respiratory diseases; n (%) | | | |
| Asthma | 1061 (15.3) | 1075 (15.5) | 0.006 |
| Bronchiectasis | 161 (2.3) | 181 (2.6) | 0.02 |
| Bronchitis, not specified as acute or chronic | 629 (9.0) | 651 (9.4) | 0.01 |
| Emphysema | 285 (4.1) | 291 (4.2) | 0.004 |
| Other chronic obstructive pulmonary disease | 612 (8.8) | 641 (9.2) | 0.01 |
| Simple and mucopurulent chronic bronchitis | 106 (1.5) | 105 (1.5) | 0.001 |
| Unspecified chronic bronchitis | 98 (1.4) | 113 (1.6) | 0.02 |
| Nicotine dependence; n (%) | 642 (9.2) | 682 (9.8) | 0.02 |
| Psychiatric comorbidities; n (%) | | | |
| Anxiety disorders | 2243 (32.2) | 2352 (33.8) | 0.03 |
| Substance misuse | 950 (13.7) | 1005 (14.4) | 0.02 |
| Mood disorders | 1723 (24.8) | 1820 (26.2) | 0.03 |
| Psychotic disorders | 122 (1.8) | 132 (1.9) | 0.01 |

| | | | |
|---|---|---|---|
| Heart disease; n (%) | | | |
| Ischemic heart diseases | 1510 (21.7) | 1560 (22.4) | 0.02 |
| Other forms of heart disease | 2563 (36.8) | 2630 (37.8) | 0.02 |
| Chronic kidney diseases; n (%) | | | |
| Chronic kidney disease (CKD) | 1091 (15.7) | 1104 (15.9) | 0.005 |
| Hypertensive chronic kidney disease | 578 (8.3) | 612 (8.8) | 0.02 |
| Chronic liver diseases; n (%) | | | |
| Alcoholic liver disease | 66 (0.9) | 67 (1.0) | 0.001 |
| Chronic hepatitis, not elsewhere classified | 50 (0.7) | 50 (0.7) | 0 |
| Chronic passive congestion of liver | 107 (1.5) | 115 (1.7) | 0.009 |
| Fatty (change of) liver, not elsewhere classified | 520 (7.5) | 527 (7.6) | 0.004 |
| Fibrosis and cirrhosis of liver | 179 (2.6) | 177 (2.5) | 0.002 |
| Hepatic failure, not elsewhere classified | 92 (1.3) | 98 (1.4) | 0.007 |
| Portal hypertension | 84 (1.2) | 91 (1.3) | 0.009 |
| Cerebral infarction; n (%) | 346 (5.0) | 369 (5.3) | 0.01 |
| Dementia; n (%) | | | |
| Alzheimer's disease | 61 (0.9) | 82 (1.2) | 0.03 |
| Dementia in other diseases classified elsewhere | 79 (1.1) | 98 (1.4) | 0.02 |
| Dementia with Lewy bodies | 40 (0.6) | 40 (0.6) | 0 |
| Frontotemporal dementia | 20 (0.3) | 20 (0.3) | 0 |
| Unspecified dementia | 121 (1.7) | 138 (2.0) | 0.02 |
| Vascular dementia | 48 (0.7) | 49 (0.7) | 0.002 |
| Neoplasms; n (%) | | | |
| Malignant neoplasms of lymphoid, hematopoietic and related tissue | 314 (4.5) | 297 (4.3) | 0.01 |
| Neoplasms (any) | 3009 (43.3) | 3099 (44.5) | 0.03 |
| Organ transplant; n (%) | | | |
| Liver Transplantation Procedures | 45 (0.6) | 49 (0.7) | 0.007 |
| Renal Transplantation Procedures | 102 (1.5) | 106 (1.5) | 0.005 |
| Psoriasis; n (%) | 189 (2.7) | 195 (2.8) | 0.005 |
| Rheumatoid arthritis; n (%) | | | |
| Other rheumatoid arthritis | 227 (3.3) | 255 (3.7) | 0.02 |
| Rheumatoid arthritis with rheumatoid factor | 89 (1.3) | 104 (1.5) | 0.02 |
| Systemic lupus erythematosus (SLE); n (%) | 75 (1.1) | 85 (1.2) | 0.01 |
| Disorders involving the immune mechanism; n (%) | 604 (8.7) | 590 (8.5) | 0.007 |

**Table S4** – Characteristics of the under-60 vaccinated and unvaccinated cohorts after propensity score matching. SMD = Standardised Mean Difference.

| | **Vaccinated** | **Unvaccinated** | **SMD** |
|---|---|---|---|
| Number | 4633 | 4633 | - |
| DEMOGRAPHICS | | | |
| Age; mean (SD) | 41.5 (11.7) | 42.1 (13.7) | 0.06 |
| Sex; n (%) | | | |
| Female | 2976 (64.2) | 3030 (65.4) | 0.02 |
| Male | 1650 (35.6) | 1598 (34.5) | 0.02 |
| Other | 30 (0.6) | 30 (0.6) | 0 |
| Race; n (%) | | | |
| White | 3069 (66.2) | 3118 (67.3) | 0.02 |
| Black or African American | 831 (17.9) | 805 (17.4) | 0.01 |
| Asian | 225 (4.9) | 192 (4.1) | 0.03 |
| American Indian or Alaska Native | 46 (1.0) | 45 (1.0) | 0.002 |
| Native Hawaiian or Other Pacific Islander | 40 (0.9) | 40 (0.9) | 0 |
| Unknown | 475 (10.3) | 488 (10.5) | 0.009 |
| Ethnicity; n (%) | | | |
| Hispanic or Latino | 628 (13.6) | 637 (13.7) | 0.006 |
| Not Hispanic or Latino | 3131 (67.6) | 3115 (67.2) | 0.007 |
| Unknown | 874 (18.9) | 881 (19.0) | 0.004 |
| Problems related to housing and economic circumstances; n (%) | 50 (1.1) | 55 (1.2) | 0.01 |
| COMORBIDITIES | | | |
| BMI; mean (SD) | 30.6 (8.0) | 30.8 (8.0) | 0.02 |
| Overweight and obesity; n (%) | 1510 (32.6) | 1552 (33.5) | 0.02 |
| Blood pressure; mean (SD) | | | |
| Diastolic | 77.1 (10.9) | 75.6 (11.6) | 0.1 |
| Systolic | 124.2 (16.2) | 122.7 (17.4) | 0.09 |
| Hypertensive diseases; n (%) | 1589 (34.3) | 1638 (35.4) | 0.02 |
| Diabetes mellitus; n (%) | | | |
| Type 1 diabetes mellitus | 146 (3.2) | 153 (3.3) | 0.009 |
| Type 2 diabetes mellitus | 772 (16.7) | 812 (17.5) | 0.02 |
| Chronic lower respiratory diseases; n (%) | | | |
| Asthma | 760 (16.4) | 743 (16.0) | 0.01 |
| Bronchiectasis | 70 (1.5) | 59 (1.3) | 0.02 |
| Bronchitis, not specified as acute or chronic | 334 (7.2) | 362 (7.8) | 0.02 |
| Emphysema | 59 (1.3) | 57 (1.2) | 0.004 |
| Other chronic obstructive pulmonary disease | 162 (3.5) | 154 (3.3) | 0.01 |
| Simple and mucopurulent chronic bronchitis | 46 (1.0) | 43 (0.9) | 0.007 |
| Unspecified chronic bronchitis | 40 (0.9) | 40 (0.9) | 0 |
| Nicotine dependence; n (%) | 420 (9.1) | 437 (9.4) | 0.01 |
| Psychiatric comorbidities; n (%) | | | |
| Anxiety disorders | 1596 (34.4) | 1630 (35.2) | 0.02 |
| Substance misuse | 588 (12.7) | 599 (12.9) | 0.007 |
| Mood disorders | 1141 (24.6) | 1176 (25.4) | 0.02 |
| Psychotic disorders | 82 (1.8) | 91 (2.0) | 0.01 |

| | | | |
|---|---|---|---|
| Heart disease; n (%) | | | |
| Ischemic heart diseases | 373 (8.1) | 383 (8.3) | 0.008 |
| Other forms of heart disease | 1007 (21.7) | 1010 (21.8) | 0.002 |
| Chronic kidney diseases; n (%) | | | |
| Chronic kidney disease (CKD) | 357 (7.7) | 354 (7.6) | 0.002 |
| Hypertensive chronic kidney disease | 197 (4.3) | 196 (4.2) | 0.001 |
| Chronic liver diseases; n (%) | | | |
| Alcoholic liver disease | 47 (1.0) | 52 (1.1) | 0.01 |
| Chronic hepatitis, not elsewhere classified | 40 (0.9) | 40 (0.9) | 0 |
| Chronic passive congestion of liver | 60 (1.3) | 64 (1.4) | 0.008 |
| Fatty (change of) liver, not elsewhere classified | 299 (6.5) | 310 (6.7) | 0.01 |
| Fibrosis and cirrhosis of liver | 72 (1.6) | 74 (1.6) | 0.003 |
| Hepatic failure, not elsewhere classified | 61 (1.3) | 58 (1.3) | 0.006 |
| Portal hypertension | 49 (1.1) | 53 (1.1) | 0.008 |
| Cerebral infarction; n (%) | 97 (2.1) | 101 (2.2) | 0.006 |
| Dementia; n (%) | | | |
| Alzheimer's disease | 0 (0.0) | 10 (0.2) | 0.07 |
| Dementia in other diseases classified elsewhere | 30 (0.6) | 30 (0.6) | 0 |
| Dementia with Lewy bodies | 0 (0.0) | 0 (0.0) | - |
| Frontotemporal dementia | 0 (0.0) | 0 (0.0) | - |
| Unspecified dementia | 20 (0.4) | 20 (0.4) | 0 |
| Vascular dementia | 10 (0.2) | 10 (0.2) | 0 |
| Neoplasms; n (%) | | | |
| Malignant neoplasms of lymphoid, hematopoietic and related tissue | 129 (2.8) | 116 (2.5) | 0.02 |
| Neoplasms (any) | 1516 (32.7) | 1540 (33.2) | 0.01 |
| Organ transplant; n (%) | | | |
| Liver Transplantation Procedures | 40 (0.9) | 40 (0.9) | 0 |
| Renal Transplantation Procedures | 75 (1.6) | 75 (1.6) | 0 |
| Psoriasis; n (%) | 96 (2.1) | 122 (2.6) | 0.04 |
| Rheumatoid arthritis; n (%) | | | |
| Other rheumatoid arthritis | 97 (2.1) | 98 (2.1) | 0.002 |
| Rheumatoid arthritis with rheumatoid factor | 55 (1.2) | 54 (1.2) | 0.002 |
| Systemic lupus erythematosus (SLE); n (%) | 78 (1.7) | 79 (1.7) | 0.002 |
| Disorders involving the immune mechanism; n (%) | 352 (7.6) | 365 (7.9) | 0.01 |

**Table S5** – Characteristics of the over-60 vaccinated and unvaccinated cohorts after propensity score matching**.** SMD = Standardised Mean Difference.

| | **Vaccinated** | **Unvaccinated** | **SMD** |
|---|---|---|---|
| Number | 4657 | 4657 | - |
| DEMOGRAPHICS | | | |
| Age; mean (SD) | 71.8 (8.0) | 71.6 (8.1) | 0.03 |
| Sex; n (%) | | | |
| Female | 2585 (55.5) | 2590 (55.6) | 0.002 |
| Male | 2070 (44.4) | 2065 (44.3) | 0.002 |
| Other | 10 (0.2) | 10 (0.2) | 0 |
| Race; n (%) | | | |
| White | 3550 (76.2) | 3576 (76.8) | 0.01 |
| Black or African American | 691 (14.8) | 684 (14.7) | 0.004 |
| Asian | 100 (2.1) | 91 (2.0) | 0.01 |
| American Indian or Alaska Native | 40 (0.9) | 40 (0.9) | 0 |
| Native Hawaiian or Other Pacific Islander | 20 (0.4) | 20 (0.4) | 0 |
| Unknown | 296 (6.4) | 280 (6.0) | 0.01 |
| Ethnicity; n (%) | | | |
| Hispanic or Latino | 318 (6.8) | 323 (6.9) | 0.004 |
| Not Hispanic or Latino | 3565 (76.6) | 3582 (76.9) | 0.009 |
| Unknown | 774 (16.6) | 752 (16.1) | 0.01 |
| Problems related to housing and economic circumstances; n (%) | 82 (1.8) | 84 (1.8) | 0.003 |
| COMORBIDITIES | | | |
| BMI; mean (SD) | 29.6 (6.8) | 29.5 (6.9) | 0.02 |
| Overweight and obesity; n (%) | 1548 (33.2) | 1560 (33.5) | 0.005 |
| Blood pressure; mean (SD) | | | |
| Diastolic | 72.4 (11.6) | 71.9 (11.5) | 0.05 |
| Systolic | 130.2 (20.4) | 128.8 (20.4) | 0.07 |
| Hypertensive diseases; n (%) | 3404 (73.1) | 3342 (71.8) | 0.03 |
| Diabetes mellitus; n (%) | | | |
| Type 1 diabetes mellitus | 275 (5.9) | 260 (5.6) | 0.01 |
| Type 2 diabetes mellitus | 1645 (35.3) | 1629 (35.0) | 0.007 |
| Chronic lower respiratory diseases; n (%) | | | |
| Asthma | 726 (15.6) | 754 (16.2) | 0.02 |
| Bronchiectasis | 148 (3.2) | 131 (2.8) | 0.02 |
| Bronchitis, not specified as acute or chronic | 528 (11.3) | 521 (11.2) | 0.005 |
| Emphysema | 337 (7.2) | 349 (7.5) | 0.01 |
| Other chronic obstructive pulmonary disease | 709 (15.2) | 694 (14.9) | 0.009 |
| Simple and mucopurulent chronic bronchitis | 104 (2.2) | 104 (2.2) | 0 |
| Unspecified chronic bronchitis | 104 (2.2) | 109 (2.3) | 0.007 |
| Nicotine dependence; n (%) | 495 (10.6) | 525 (11.3) | 0.02 |
| Psychiatric comorbidities; n (%) | | | |
| Anxiety disorders | 1487 (31.9) | 1504 (32.3) | 0.008 |
| Substance misuse | 736 (15.8) | 771 (16.6) | 0.02 |
| Mood disorders | 1251 (26.9) | 1242 (26.7) | 0.004 |
| Psychotic disorders | 95 (2.0) | 106 (2.3) | 0.02 |

| | | | |
|---|---|---|---|
| Heart disease; n (%) | | | |
| Ischemic heart diseases | 1620 (34.8) | 1627 (34.9) | 0.003 |
| Other forms of heart disease | 2446 (52.5) | 2408 (51.7) | 0.02 |
| Chronic kidney diseases; n (%) | | | |
| Chronic kidney disease (CKD) | 1106 (23.7) | 1125 (24.2) | 0.01 |
| Hypertensive chronic kidney disease | 566 (12.2) | 591 (12.7) | 0.02 |
| Chronic liver diseases; n (%) | | | |
| Alcoholic liver disease | 51 (1.1) | 60 (1.3) | 0.02 |
| Chronic hepatitis, not elsewhere classified | 48 (1.0) | 47 (1.0) | 0.002 |
| Chronic passive congestion of liver | 85 (1.8) | 87 (1.9) | 0.003 |
| Fatty (change of) liver, not elsewhere classified | 397 (8.5) | 401 (8.6) | 0.003 |
| Fibrosis and cirrhosis of liver | 174 (3.7) | 193 (4.1) | 0.02 |
| Hepatic failure, not elsewhere classified | 77 (1.7) | 77 (1.7) | 0 |
| Portal hypertension | 74 (1.6) | 92 (2.0) | 0.03 |
| Cerebral infarction; n (%) | 392 (8.4) | 400 (8.6) | 0.006 |
| Dementia; n (%) | | | |
| Alzheimer's disease | 69 (1.5) | 79 (1.7) | 0.02 |
| Dementia in other diseases classified elsewhere | 89 (1.9) | 99 (2.1) | 0.02 |
| Dementia with Lewy bodies | 40 (0.9) | 40 (0.9) | 0 |
| Frontotemporal dementia | 20 (0.4) | 30 (0.6) | 0.03 |
| Unspecified dementia | 172 (3.7) | 194 (4.2) | 0.02 |
| Vascular dementia | 58 (1.2) | 76 (1.6) | 0.03 |
| Neoplasms; n (%) | | | |
| Malignant neoplasms of lymphoid, hematopoietic and related tissue | 251 (5.4) | 243 (5.2) | 0.008 |
| Neoplasms (any) | 2487 (53.4) | 2486 (53.4) | 0.0004 |
| Organ transplant; n (%) | | | |
| Liver Transplantation Procedures | 40 (0.9) | 30 (0.6) | 0.02 |
| Renal Transplantation Procedures | 56 (1.2) | 47 (1.0) | 0.02 |
| Psoriasis; n (%) | 140 (3.0) | 139 (3.0) | 0.001 |
| Rheumatoid arthritis; n (%) | | | |
| Other rheumatoid arthritis | 244 (5.2) | 230 (4.9) | 0.01 |
| Rheumatoid arthritis with rheumatoid factor | 84 (1.8) | 86 (1.8) | 0.003 |
| Systemic lupus erythematosus (SLE); n (%) | 53 (1.1) | 53 (1.1) | 0 |
| Disorders involving the immune mechanism; n (%) | 413 (8.9) | 424 (9.1) | 0.008 |

**Table S6** – Percent incidences and HRs (95% CI) for all outcomes (each as part of a composite endpoint with death as the other component) in the propensity score matched vaccinated and unvaccinated cohorts (*n*=9479 individuals in each cohort).

| Outcome | Incidence % (95% CI) in vaccinated cohort | Incidence % (95% CI) in unvaccinated cohort | HR (95% CI) | p-value |
|---|---|---|---|---|
| Abdominal symptoms | 19.86 (18.32-21.36) | 19.76 (18.28-21.22) | 1.02 (0.93-1.12) | 0.62 |
| Abnormal breathing | 25.44 (23.85-27.00) | 25.39 (23.87-26.88) | 0.96 (0.89-1.04) | 0.36 |
| Anosmia | 4.41 (3.68-5.13) | 4.77 (4.15-5.39) | 0.82 (0.69-0.98) | 0.03 |
| Anxiety disorder | 24.74 (23.14-26.30) | 23.68 (22.12-25.21) | 1.06 (0.97-1.15) | 0.2 |
| Anxiety/Depression | 32.33 (30.60-34.02) | 30.57 (28.89-32.22) | 1.07 (1.00-1.15) | 0.06 |
| Arrhythmia | 21.53 (20.10-22.94) | 22.31 (20.95-23.66) | 0.93 (0.86-1.01) | 0.09 |
| Cardiac failure | 13.77 (12.55-14.97) | 13.98 (12.89-15.05) | 0.95 (0.86-1.05) | 0.3 |
| Cardiomyopathy | 8.37 (7.38-9.36) | 8.52 (7.64-9.39) | 0.92 (0.80-1.05) | 0.22 |
| Cerebral haemorrhage | 4.32 (3.66-4.97) | 4.87 (4.23-5.49) | 0.82 (0.68-0.97) | 0.02 |
| Chest/Throat pain | 16.31 (14.84-17.76) | 16.31 (14.99-17.60) | 0.98 (0.88-1.08) | 0.66 |
| Cognitive symptoms | 12.39 (11.18-13.58) | 12.87 (11.78-13.96) | 0.93 (0.83-1.04) | 0.18 |
| Coronary disease | 7.85 (6.83-8.85) | 7.94 (7.06-8.81) | 0.87 (0.75-1.00) | 0.05 |
| Death | 3.74 (3.12-4.35) | 4.50 (3.89-5.10) | 0.75 (0.62-0.91) | 0.003 |
| Fatigue | 20.17 (18.56-21.75) | 20.18 (18.75-21.58) | 0.89 (0.81-0.97) | 0.01 |
| GORD | 25.92 (24.26-27.55) | 25.13 (23.56-26.67) | 1.02 (0.94-1.11) | 0.59 |
| Hair loss | 5.50 (4.70-6.29) | 6.88 (6.05-7.70) | 0.75 (0.64-0.88) | 0.0005 |
| Headache | 15.89 (14.54-17.21) | 14.60 (13.36-15.83) | 1.07 (0.96-1.18) | 0.23 |
| Hospitalisation | 17.85 (16.72-18.97) | 19.47 (18.35-20.57) | 0.90 (0.84-0.97) | 0.008 |
| Hypercoagulopathy/DVT/PE | 9.96 (8.95-10.95) | 12.00 (10.96-13.03) | 0.81 (0.72-0.91) | 0.0003 |
| Hyperlipidaemia | 44.74 (42.71-46.70) | 43.84 (41.96-45.65) | 1.02 (0.96-1.08) | 0.58 |
| Hypertension | 53.69 (51.61-55.69) | 52.67 (50.79-54.47) | 0.98 (0.93-1.04) | 0.51 |
| Hypoxaemia | 10.95 ( 9.97-11.92) | 14.25 (13.26-15.22) | 0.72 (0.65-0.80) | <0.0001 |
| ICU admission | 6.23 (5.51-6.94) | 8.03 (7.25-8.81) | 0.75 (0.65-0.85) | <0.0001 |
| Interstitial lung disease | 6.26 (5.47-7.06) | 7.37 (6.52-8.21) | 0.84 (0.73-0.98) | 0.02 |
| Intubation/Ventilation | 4.32 (3.70-4.93) | 5.66 (5.01-6.30) | 0.72 (0.61-0.84) | <0.0001 |
| Ischaemic stroke | 6.88 (6.01-7.75) | 7.61 (6.73-8.47) | 0.84 (0.73-0.97) | 0.02 |
| Joint pain | 21.07 (19.39-22.72) | 20.63 (19.08-22.15) | 0.98 (0.89-1.08) | 0.7 |
| Kidney disease | 18.14 (16.85-19.42) | 19.90 (18.61-21.18) | 0.94 (0.86-1.02) | 0.15 |
| Liver disease | 10.06 (9.06-11.04) | 10.80 ( 9.78-11.80) | 0.94 (0.84-1.06) | 0.33 |
| Long COVID feature (any) | 64.18 (62.26-66.00) | 65.57 (63.68-67.36) | 1.01 (0.96-1.05) | 0.83 |
| Mood disorder | 22.47 (20.87-24.04) | 20.55 (19.15-21.93) | 1.05 (0.96-1.15) | 0.27 |
| Myalgia | 6.90 (5.96-7.82) | 8.22 (7.28-9.16) | 0.78 (0.67-0.91) | 0.001 |
| Myocarditis | 3.97 (3.33-4.62) | 4.70 (4.06-5.33) | 0.77 (0.64-0.92) | 0.004 |
| Myoneural junction/muscle disease | 4.44 (3.79-5.09) | 5.46 (4.76-6.15) | 0.78 (0.66-0.92) | 0.004 |
| Nerve/nerve root/plexus disorder | 8.37 (7.32-9.40) | 8.92 (7.95- 9.89) | 0.82 (0.72-0.95) | 0.006 |
| Obesity | 39.63 (37.81-41.39) | 40.15 (38.38-41.87) | 0.97 (0.91-1.04) | 0.39 |
| Other pain | 18.09 (16.60-19.56) | 20.01 (18.50-21.49) | 0.90 (0.81-0.99) | 0.03 |
| Oxygen requirement | 11.81 (10.73-12.88) | 13.83 (12.75-14.90) | 0.83 (0.75-0.92) | 0.0002 |
| Peripheral neuropathy | 8.83 (7.77- 9.87) | 9.10 (8.06-10.13) | 0.93 (0.81-1.07) | 0.29 |
| Psychotic disorder | 4.36 (3.72-5.00) | 5.62 (4.92-6.30) | 0.75 (0.63-0.89) | 0.0008 |
| Respiratory failure | 10.28 (9.32-11.23) | 13.95 (12.94-14.94) | 0.70 (0.63-0.78) | <0.0001 |
| Seizures | 5.15 (4.39-5.90) | 6.29 (5.54-7.04) | 0.73 (0.62-0.86) | 0.0001 |

| | | | | |
|---|---|---|---|---|
| Sleep disorders | 23.00 (21.34-24.62) | 23.04 (21.52-24.53) | 0.95 (0.88-1.04) | 0.29 |
| Type 2 diabetes mellitus | 27.63 (26.10-29.13) | 27.43 (25.99-28.85) | 0.97 (0.90-1.05) | 0.45 |
| Urticaria | 4.34 (3.67-5.00) | 5.05 (4.41-5.69) | 0.77 (0.65-0.92) | 0.004 |

**Table S7** – P-values for the test of proportional hazards (obtained using the generalized Schoenfeld test) for the primary analysis. A value lower than 0.05 indicates evidence for non-proportional hazards.

| Outcome | p-value for the proportionality assumption |
|---|---|
| Abdominal symptoms | 0.35 |
| Abnormal breathing | 0.49 |
| Anosmia | 0.025 |
| Anxiety disorder | 0.96 |
| Anxiety/Depression | 0.85 |
| Arrhythmia | 0.88 |
| Cardiac failure | 0.63 |
| Cardiomyopathy | 0.13 |
| Cerebral haemorrhage | 0.029 |
| Chest/Throat pain | 0.29 |
| Cognitive symptoms | 0.79 |
| Coronary disease | 0.064 |
| Death | 0.011 |
| Fatigue | 0.009 |
| GORD | 0.62 |
| Hair loss | 0.082 |
| Headache | 0.54 |
| Hospitalisation | 0.052 |
| Hypercoagulopathy/DVT/PE | 0.62 |
| Hyperlipidaemia | 0.12 |
| Hypertension | 0.85 |
| Hypoxaemia | 0.76 |
| ICU admission | 0.13 |
| Interstitial lung disease | 0.49 |
| Intubation/Ventilation | 0.075 |
| Ischaemic stroke | 0.054 |
| Joint pain | 0.16 |
| Kidney disease | 0.0053 |
| Liver disease | 0.83 |
| Long COVID feature (any) | 0.0047 |
| Mood disorder | 0.81 |
| Myalgia | 0.082 |
| Myocarditis | 0.016 |
| Myoneural junction/muscle disease | 0.08 |
| Nerve/nerve root/plexus disorder | 0.00018 |
| Obesity | 0.4 |
| Other pain | 0.72 |
| Oxygen requirement | 0.72 |
| Peripheral neuropathy | 0.052 |

| | |
|---|---|
| Psychotic disorder | 0.14 |
| Respiratory failure | 0.48 |
| Seizures | 0.019 |
| Sleep disorders | 0.5 |
| Type 2 diabetes mellitus | 0.99 |
| Urticaria | 0.0083 |

**Table S8** – Percent incidences and HRs (95% CI) for all outcomes (each as part of a composite endpoint with death as the other component) in the matched vaccinated (2 doses only) and unvaccinated cohorts (*n*=6957 individuals in each cohort).

| Outcome | Incidence % (95% CI) in vaccinated cohort | Incidence % (95% CI) in unvaccinated cohort | HR (95% CI) | p-value |
|---|---|---|---|---|
| Abdominal symptoms | 20.08 (18.06-22.04) | 21.07 (18.99-23.09) | 1.00 (0.89-1.12) | 0.99 |
| Abnormal breathing | 24.41 (22.42-26.35) | 25.29 (23.53-27.01) | 0.89 (0.81-0.98) | 0.01 |
| Anosmia | 4.13 (3.24-5.02) | 5.36 (4.56-6.15) | 0.68 (0.55-0.84) | 0.0004 |
| Anxiety disorder | 22.96 (21.05-24.82) | 25.15 (23.06-27.19) | 1.00 (0.90-1.10) | 0.93 |
| Anxiety/Depression | 31.04 (28.75-33.26) | 32.27 (29.94-34.52) | 1.03 (0.94-1.12) | 0.55 |
| Arrhythmia | 20.47 (18.83-22.07) | 23.37 (21.53-25.16) | 0.90 (0.81-0.99) | 0.03 |
| Cardiac failure | 13.83 (12.26-15.36) | 14.84 (13.31-16.34) | 0.95 (0.84-1.07) | 0.37 |
| Cardiomyopathy | 8.79 (7.47-10.10) | 9.22 (8.07-10.36) | 0.85 (0.72-0.99) | 0.04 |
| Cerebral haemorrhage | 3.83 (3.10-4.55) | 5.35 (4.54-6.16) | 0.69 (0.56-0.85) | 0.0005 |
| Chest/Throat pain | 16.72 (14.56-18.82) | 16.67 (14.94-18.37) | 0.92 (0.82-1.04) | 0.2 |
| Cognitive symptoms | 11.94 (10.49-13.37) | 12.95 (11.68-14.21) | 0.87 (0.76-0.99) | 0.04 |
| Coronary disease | 8.90 (7.32-10.46) | 7.90 (6.84-8.95) | 0.85 (0.72-1.01) | 0.06 |
| Death | 3.35 (2.66-4.03) | 4.98 (4.21-5.74) | 0.62 (0.49-0.78) | <0.0001 |
| Fatigue | 20.75 (18.41-23.03) | 20.33 (18.62-22.01) | 0.86 (0.77-0.96) | 0.005 |
| GORD | 26.94 (24.68-29.14) | 25.64 (23.55-27.68) | 1.02 (0.93-1.13) | 0.65 |
| Hair loss | 4.63 (3.75-5.49) | 6.64 (5.66-7.62) | 0.66 (0.54-0.81) | <0.0001 |
| Headache | 16.13 (14.43-17.80) | 15.62 (14.01-17.20) | 1.00 (0.88-1.13) | 0.95 |
| Hospitalisation | 18.59 (16.93-20.21) | 19.57 (18.17-20.94) | 0.91 (0.83-1.00) | 0.04 |
| Hypercoagulopathy/DVT/PE | 9.91 (8.63-11.18) | 11.81 (10.45-13.14) | 0.80 (0.70-0.92) | 0.002 |
| Hyperlipidaemia | 49.72 (46.64-52.63) | 44.74 (42.33-47.06) | 1.05 (0.98-1.13) | 0.17 |
| Hypertension | 57.88 (54.73-60.82) | 52.55 (50.08-54.89) | 1.05 (0.98-1.12) | 0.15 |
| Hypoxaemia | 10.65 (9.27-12.02) | 14.82 (13.49-16.14) | 0.66 (0.58-0.74) | <0.0001 |
| ICU admission | 6.24 (5.30-7.18) | 8.21 (7.25-9.16) | 0.72 (0.61-0.85) | <0.0001 |
| Interstitial lung disease | 5.97 (5.00-6.93) | 8.24 (7.16-9.32) | 0.74 (0.62-0.88) | 0.0006 |
| Intubation/Ventilation | 4.20 (3.45-4.95) | 6.22 (5.38-7.05) | 0.63 (0.52-0.76) | <0.0001 |
| Ischaemic stroke | 6.77 (5.68-7.85) | 8.14 (7.05-9.23) | 0.77 (0.64-0.91) | 0.002 |
| Joint pain | 21.07 (18.84-23.23) | 22.55 (20.40-24.63) | 0.91 (0.81-1.02) | 0.11 |
| Kidney disease | 19.24 (17.48-20.96) | 19.80 (18.14-21.44) | 0.97 (0.87-1.07) | 0.52 |
| Liver disease | 10.69 (9.37-11.99) | 11.84 (10.50-13.16) | 0.93 (0.81-1.07) | 0.31 |
| Long COVID feature (any) | 64.94 (62.30-67.40) | 65.63 (63.08-68.01) | 1.00 (0.95-1.06) | 0.98 |
| Mood disorder | 22.55 (20.30-24.75) | 21.84 (19.83-23.81) | 1.03 (0.92-1.14) | 0.63 |
| Myalgia | 6.37 (5.32-7.41) | 9.27 (7.98-10.53) | 0.70 (0.59-0.84) | <0.0001 |
| Myocarditis | 3.49 (2.79-4.18) | 5.08 (4.31-5.85) | 0.64 (0.51-0.80) | <0.0001 |
| Myoneural junction/muscle disease | 4.15 (3.40-4.89) | 6.01 (5.12-6.89) | 0.67 (0.54-0.82) | <0.0001 |
| Nerve/nerve root/plexus disorder | 7.66 (6.43-8.88) | 8.69 (7.51- 9.86) | 0.78 (0.66-0.92) | 0.003 |
| Obesity | 37.16 (34.83-39.40) | 37.27 (35.09-39.38) | 0.97 (0.89-1.04) | 0.37 |
| Other pain | 17.75 (15.86-19.60) | 21.22 (19.16-23.23) | 0.85 (0.76-0.96) | 0.007 |
| Oxygen requirement | 10.84 ( 9.60-12.07) | 14.00 (12.65-15.33) | 0.75 (0.67-0.85) | <0.0001 |
| Peripheral neuropathy | 8.27 (7.04-9.48) | 9.54 (8.18-10.88) | 0.82 (0.70-0.97) | 0.02 |
| Psychotic disorder | 4.11 (3.33-4.88) | 6.00 (5.07-6.93) | 0.65 (0.52-0.79) | <0.0001 |
| Respiratory failure | 10.12 (8.80-11.42) | 14.76 (13.40-16.10) | 0.63 (0.56-0.71) | <0.0001 |
| Seizures | 5.13 (4.12-6.14) | 6.59 (5.67-7.50) | 0.67 (0.55-0.82) | <0.0001 |

| | | | | |
|---|---|---|---|---|
| Sleep disorders | 22.85 (20.60-25.03) | 22.92 (20.89-24.89) | 0.96 (0.86-1.06) | 0.41 |
| Type 2 diabetes mellitus | 28.89 (26.80-30.92) | 28.23 (26.29-30.12) | 1.03 (0.94-1.12) | 0.55 |
| Urticaria | 3.98 (3.20-4.76) | 5.58 (4.75-6.41) | 0.65 (0.52-0.80) | <0.0001 |

**Table S9** – Percent incidences and HRs (95% CI) for all outcomes (each as part of a composite endpoint with death as the other component) in the matched vaccinated (1 dose only) and unvaccinated cohorts (*n*=2996 individuals in each cohort).

| Outcome | Incidence % (95% CI) in vaccinated cohort | Incidence % (95% CI) in unvaccinated cohort | HR (95% CI) | p-value |
|---|---|---|---|---|
| Abdominal symptoms | 18.87 (16.84-20.86) | 19.11 (17.03-21.14) | 0.97 (0.83-1.12) | 0.65 |
| Abnormal breathing | 24.32 (22.24-26.35) | 24.23 (22.07-26.34) | 1.00 (0.88-1.13) | 0.95 |
| Anosmia | 3.82 (2.99-4.64) | 5.16 (4.10-6.22) | 0.82 (0.62-1.09) | 0.17 |
| Anxiety disorder | 24.47 (22.25-26.62) | 25.86 (23.52-28.14) | 0.95 (0.84-1.08) | 0.43 |
| Anxiety/Depression | 30.36 (28.03-32.61) | 31.91 (29.49-34.24) | 0.96 (0.85-1.07) | 0.43 |
| Arrhythmia | 20.26 (18.38-22.09) | 21.04 (19.09-22.95) | 0.96 (0.84-1.09) | 0.5 |
| Cardiac failure | 12.46 (10.99-13.90) | 12.72 (11.12-14.30) | 1.01 (0.86-1.20) | 0.86 |
| Cardiomyopathy | 7.21 (6.04-8.36) | 7.21 (5.99-8.42) | 1.02 (0.81-1.27) | 0.88 |
| Cerebral haemorrhage | 3.68 (2.89-4.47) | 5.06 (3.97-6.14) | 0.82 (0.61-1.09) | 0.17 |
| Chest/Throat pain | 15.23 (13.32-17.09) | 14.74 (12.91-16.53) | 1.03 (0.87-1.21) | 0.75 |
| Cognitive symptoms | 10.92 (9.49-12.32) | 13.61 (11.87-15.31) | 0.81 (0.68-0.97) | 0.02 |
| Coronary disease | 6.56 (5.41-7.71) | 6.84 (5.60-8.05) | 0.95 (0.75-1.20) | 0.67 |
| Death | 2.95 (2.26-3.63) | 4.73 (3.70-5.74) | 0.72 (0.53-0.98) | 0.03 |
| Fatigue | 18.28 (16.25-20.25) | 18.93 (17.04-20.78) | 0.87 (0.76-1.01) | 0.07 |
| GORD | 25.14 (22.87-27.33) | 26.09 (23.65-28.44) | 0.99 (0.87-1.12) | 0.84 |
| Hair loss | 4.59 (3.65-5.53) | 6.75 (5.46-8.02) | 0.74 (0.57-0.97) | 0.03 |
| Headache | 15.92 (14.10-17.71) | 15.47 (13.48-17.41) | 1.12 (0.95-1.32) | 0.16 |
| Hospitalisation | 17.99 (16.41-19.54) | 20.57 (18.75-22.35) | 0.90 (0.80-1.02) | 0.11 |
| Hypercoagulopathy/DVT/PE | 8.68 (7.45- 9.89) | 10.45 (8.99-11.89) | 0.88 (0.73-1.07) | 0.19 |
| Hyperlipidaemia | 42.91 (40.19-45.50) | 44.36 (41.50-47.08) | 0.98 (0.89-1.08) | 0.72 |
| Hypertension | 51.16 (48.45-53.72) | 53.90 (50.96-56.68) | 0.98 (0.90-1.07) | 0.64 |
| Hypoxaemia | 10.58 (9.30-11.83) | 13.50 (12.01-14.96) | 0.78 (0.66-0.92) | 0.003 |
| ICU admission | 5.51 (4.58-6.42) | 7.98 (6.72-9.22) | 0.72 (0.58-0.90) | 0.004 |
| Interstitial lung disease | 5.10 (4.15-6.05) | 6.69 (5.56-7.80) | 0.79 (0.62-1.00) | 0.05 |
| Intubation/Ventilation | 3.75 (2.97-4.52) | 5.67 (4.59-6.74) | 0.71 (0.54-0.93) | 0.01 |
| Ischaemic stroke | 6.32 (5.20-7.43) | 6.73 (5.47-7.97) | 0.99 (0.78-1.25) | 0.92 |
| Joint pain | 19.96 (17.75-22.12) | 21.00 (18.71-23.22) | 0.95 (0.82-1.10) | 0.52 |
| Kidney disease | 16.05 (14.39-17.68) | 19.78 (17.80-21.70) | 0.86 (0.74-0.99) | 0.03 |
| Liver disease | 9.17 (7.84-10.48) | 10.18 (8.75-11.60) | 0.91 (0.75-1.10) | 0.33 |
| Long COVID feature (any) | 64.33 (61.54-66.92) | 66.26 (63.46-68.85) | 0.96 (0.89-1.03) | 0.24 |
| Mood disorder | 20.17 (18.09-22.19) | 21.27 (19.21-23.29) | 0.93 (0.81-1.07) | 0.31 |
| Myalgia | 5.41 (4.32-6.48) | 7.39 (6.04-8.72) | 0.75 (0.59-0.97) | 0.03 |
| Myocarditis | 3.24 (2.51-3.96) | 4.94 (3.84-6.03) | 0.77 (0.57-1.04) | 0.09 |
| Myoneural junction/muscle disease | 3.58 (2.79-4.37) | 4.98 (3.97-5.99) | 0.77 (0.57-1.02) | 0.07 |
| Nerve/nerve root/plexus disorder | 7.04 (5.76-8.30) | 8.79 (7.32-10.25) | 0.81 (0.64-1.01) | 0.06 |
| Obesity | 42.08 (39.50-44.55) | 43.89 (41.17-46.48) | 0.98 (0.89-1.08) | 0.7 |
| Other pain | 17.97 (15.91-19.99) | 19.43 (17.27-21.53) | 0.87 (0.75-1.01) | 0.07 |
| Oxygen requirement | 11.61 (10.14-13.06) | 13.54 (11.99-15.05) | 0.84 (0.71-0.99) | 0.04 |
| Peripheral neuropathy | 7.96 (6.68-9.22) | 7.88 (6.50-9.24) | 1.05 (0.84-1.31) | 0.67 |
| Psychotic disorder | 3.61 (2.85-4.36) | 5.98 (4.87-7.08) | 0.65 (0.49-0.85) | 0.002 |
| Respiratory failure | 9.68 (8.44-10.89) | 12.89 (11.40-14.36) | 0.75 (0.64-0.89) | 0.001 |
| Seizures | 4.43 (3.54-5.32) | 6.16 (5.07-7.25) | 0.71 (0.55-0.92) | 0.009 |

| | | | | |
|---|---|---|---|---|
| Sleep disorders | 21.74 (19.59-23.83) | 22.26 (20.01-24.45) | 0.96 (0.84-1.10) | 0.57 |
| Type 2 diabetes mellitus | 25.56 (23.54-27.53) | 27.02 (24.85-29.13) | 0.95 (0.85-1.07) | 0.41 |
| Urticaria | 3.78 (2.95-4.60) | 5.11 (4.02-6.19) | 0.81 (0.60-1.08) | 0.14 |

**Table S10** – HRs (95% CI) for outcomes (each as part of a composite endpoint with death as the other component) comparing vaccinated (1 dose only) to unvaccinated individuals (*n*=2996 in each group), and vaccinated (2 doses only) to unvaccinated individuals (*n*=6957 in each group), both identical to those presented in Table S8 and S9 respectively, as well as their ratio (95% CI) obtained by fitting an interaction term in the Cox model, and the p-value for the test of the null hypothesis that the latter ratio is equal to 1.

| Outcome | HR (95% CI) in the 1-dose cohort | HR (95% CI) in the 2-dose cohort | Ratio between HR (95% CI) | p-value |
|---|---|---|---|---|
| Abdominal symptoms | 0.97 (0.83-1.12) | 1.00 (0.89-1.12) | 0.97 (0.81-1.16) | 0.73 |
| Abnormal breathing | 1.00 (0.88-1.13) | 0.89 (0.81-0.98) | 1.12 (0.96-1.31) | 0.16 |
| Anosmia | 0.82 (0.62-1.09) | 0.68 (0.55-0.84) | 1.20 (0.84-1.70) | 0.32 |
| Anxiety disorder | 0.95 (0.84-1.08) | 1.00 (0.90-1.10) | 0.96 (0.81-1.12) | 0.57 |
| Anxiety/Depression | 0.96 (0.85-1.07) | 1.03 (0.94-1.12) | 0.93 (0.81-1.07) | 0.32 |
| Arrhythmia | 0.96 (0.84-1.09) | 0.90 (0.81-0.99) | 1.06 (0.90-1.25) | 0.46 |
| Cardiac failure | 1.01 (0.86-1.20) | 0.95 (0.84-1.07) | 1.07 (0.87-1.32) | 0.51 |
| Cardiomyopathy | 1.02 (0.81-1.27) | 0.85 (0.72-0.99) | 1.20 (0.91-1.58) | 0.19 |
| Cerebral haemorrhage | 0.82 (0.61-1.09) | 0.69 (0.56-0.85) | 1.19 (0.83-1.70) | 0.35 |
| Chest/Throat pain | 1.03 (0.87-1.21) | 0.92 (0.82-1.04) | 1.11 (0.91-1.36) | 0.31 |
| Cognitive symptoms | 0.81 (0.68-0.97) | 0.87 (0.76-0.99) | 0.93 (0.75-1.16) | 0.54 |
| Coronary disease | 0.95 (0.75-1.20) | 0.85 (0.72-1.01) | 1.11 (0.84-1.49) | 0.46 |
| Death | 0.72 (0.53-0.98) | 0.62 (0.49-0.78) | 1.16 (0.79-1.69) | 0.46 |
| Fatigue | 0.87 (0.76-1.01) | 0.86 (0.77-0.96) | 1.02 (0.85-1.22) | 0.84 |
| GORD | 0.99 (0.87-1.12) | 1.02 (0.93-1.13) | 0.96 (0.82-1.13) | 0.65 |
| Hair loss | 0.74 (0.57-0.97) | 0.66 (0.54-0.81) | 1.13 (0.81-1.58) | 0.47 |
| Headache | 1.12 (0.95-1.32) | 1.00 (0.88-1.13) | 1.12 (0.92-1.38) | 0.26 |
| Hospitalisation | 0.90 (0.80-1.02) | 0.91 (0.83-1.00) | 0.99 (0.85-1.16) | 0.9 |
| Hypercoagulopathy/DVT/PE | 0.88 (0.73-1.07) | 0.80 (0.70-0.92) | 1.09 (0.86-1.39) | 0.46 |
| Hyperlipidaemia | 0.98 (0.89-1.08) | 1.05 (0.98-1.13) | 0.93 (0.83-1.05) | 0.26 |
| Hypertension | 0.98 (0.90-1.07) | 1.05 (0.98-1.12) | 0.93 (0.84-1.04) | 0.22 |
| Hypoxaemia | 0.78 (0.66-0.92) | 0.66 (0.58-0.74) | 1.18 (0.96-1.45) | 0.11 |
| ICU admission | 0.72 (0.58-0.90) | 0.72 (0.61-0.85) | 1.00 (0.76-1.32) | 0.99 |
| Interstitial lung disease | 0.79 (0.62-1.00) | 0.74 (0.62-0.88) | 1.07 (0.79-1.45) | 0.65 |
| Intubation/Ventilation | 0.71 (0.54-0.93) | 0.63 (0.52-0.76) | 1.12 (0.80-1.56) | 0.5 |
| Ischaemic stroke | 0.99 (0.78-1.25) | 0.77 (0.64-0.91) | 1.28 (0.96-1.72) | 0.098 |
| Joint pain | 0.95 (0.82-1.10) | 0.91 (0.81-1.02) | 1.04 (0.87-1.26) | 0.65 |
| Kidney disease | 0.86 (0.74-0.99) | 0.97 (0.87-1.07) | 0.88 (0.74-1.05) | 0.17 |
| Liver disease | 0.91 (0.75-1.10) | 0.93 (0.81-1.07) | 0.98 (0.77-1.24) | 0.85 |
| Long COVID feature (any) | 0.96 (0.89-1.03) | 1.00 (0.95-1.06) | 0.96 (0.87-1.05) | 0.34 |
| Mood disorder | 0.93 (0.81-1.07) | 1.03 (0.92-1.14) | 0.91 (0.76-1.08) | 0.27 |
| Myalgia | 0.75 (0.59-0.97) | 0.70 (0.59-0.84) | 1.08 (0.79-1.46) | 0.63 |
| Myocarditis | 0.77 (0.57-1.04) | 0.64 (0.51-0.80) | 1.21 (0.84-1.76) | 0.31 |
| Myoneural junction/muscle disease | 0.77 (0.57-1.02) | 0.67 (0.54-0.82) | 1.15 (0.81-1.63) | 0.45 |
| Nerve/nerve root/plexus disorder | 0.81 (0.64-1.01) | 0.78 (0.66-0.92) | 1.04 (0.78-1.37) | 0.8 |
| Obesity | 0.98 (0.89-1.08) | 0.97 (0.89-1.04) | 1.02 (0.90-1.15) | 0.78 |
| Other pain | 0.87 (0.75-1.01) | 0.85 (0.76-0.96) | 1.02 (0.84-1.23) | 0.84 |
| Oxygen requirement | 0.84 (0.71-0.99) | 0.75 (0.67-0.85) | 1.11 (0.91-1.36) | 0.31 |
| Peripheral neuropathy | 1.05 (0.84-1.31) | 0.82 (0.70-0.97) | 1.28 (0.97-1.69) | 0.082 |
| Psychotic disorder | 0.65 (0.49-0.85) | 0.65 (0.52-0.79) | 1.01 (0.71-1.42) | 0.97 |

| | | | | |
|---|---|---|---|---|
| Respiratory failure | 0.75 (0.64-0.89) | 0.63 (0.56-0.71) | 1.20 (0.97-1.48) | 0.098 |
| Seizures | 0.71 (0.55-0.92) | 0.67 (0.55-0.82) | 1.05 (0.76-1.46) | 0.76 |
| Sleep disorders | 0.96 (0.84-1.10) | 0.96 (0.86-1.06) | 1.00 (0.85-1.19) | 0.97 |
| Type 2 diabetes mellitus | 0.95 (0.85-1.07) | 1.03 (0.94-1.12) | 0.93 (0.80-1.07) | 0.32 |
| Urticaria | 0.81 (0.60-1.08) | 0.65 (0.52-0.80) | 1.24 (0.87-1.78) | 0.23 |

**Table S11** – Percent incidences and HRs (95% CI) for all outcomes (each as part of a composite endpoint with death as the other component) in the matched under-60 vaccinated and unvaccinated cohorts (*n*=4633 individuals in each cohort).

| Outcome | Incidence % (95% CI) in vaccinated cohort | Incidence % (95% CI) in unvaccinated cohort | HR (95% CI) | p-value |
|---|---|---|---|---|
| Abdominal symptoms | 20.00 (17.22-22.70) | 19.22 (16.79-21.58) | 0.99 (0.85-1.15) | 0.87 |
| Abnormal breathing | 20.70 (18.38-22.96) | 20.44 (18.46-22.37) | 0.88 (0.77-1.00) | 0.05 |
| Anosmia | 2.17 (1.25-3.09) | 2.85 (2.06-3.62) | 0.62 (0.42-0.92) | 0.02 |
| Anxiety disorder | 26.49 (23.85-29.03) | 22.32 (20.20-24.37) | 1.12 (1.00-1.27) | 0.06 |
| Anxiety/Depression | 32.49 (29.71-35.16) | 29.02 (26.48-31.47) | 1.09 (0.97-1.21) | 0.14 |
| Arrhythmia | 12.48 (10.69-14.25) | 11.92 (10.41-13.41) | 0.91 (0.77-1.08) | 0.28 |
| Cardiac failure | 5.39 (4.16-6.60) | 5.88 (4.86-6.90) | 0.85 (0.67-1.08) | 0.19 |
| Cardiomyopathy | 5.27 (3.77-6.74) | 3.95 (3.11-4.78) | 1.00 (0.75-1.32) | 1 |
| Cerebral haemorrhage | 1.37 (0.84-1.89) | 2.50 (1.83-3.18) | 0.47 (0.31-0.73) | 0.0004 |
| Chest/Throat pain | 14.68 (12.10-17.18) | 14.26 (12.24-16.23) | 0.91 (0.77-1.08) | 0.27 |
| Cognitive symptoms | 5.32 (4.13-6.50) | 6.19 (4.93-7.42) | 0.82 (0.64-1.06) | 0.13 |
| Coronary disease | 2.71 (1.68-3.72) | 3.28 (2.49-4.05) | 0.60 (0.42-0.86) | 0.004 |
| Death | 1.17 (0.67-1.68) | 2.25 (1.60-2.89) | 0.43 (0.26-0.68) | 0.0002 |
| Fatigue | 15.49 (13.03-17.89) | 12.98 (11.22-14.70) | 0.96 (0.81-1.13) | 0.61 |
| GORD | 21.81 (19.01-24.51) | 19.08 (16.72-21.38) | 1.08 (0.94-1.25) | 0.29 |
| Hair loss | 2.87 (2.05-3.69) | 4.88 (3.75-5.99) | 0.59 (0.42-0.81) | 0.001 |
| Headache | 18.14 (15.70-20.51) | 17.76 (15.58-19.88) | 1.03 (0.89-1.20) | 0.69 |
| Hospitalisation | 9.84 (8.44-11.23) | 11.63 (10.31-12.93) | 0.81 (0.70-0.94) | 0.006 |
| Hypercoagulopathy/DVT/PE | 5.84 (4.51-7.15) | 6.92 (5.78-8.04) | 0.72 (0.57-0.90) | 0.004 |
| Hyperlipidaemia | 27.45 (24.67-30.13) | 27.44 (24.75-30.04) | 1.01 (0.90-1.14) | 0.83 |
| Hypertension | 35.59 (32.45-38.58) | 35.02 (32.27-37.66) | 0.97 (0.88-1.08) | 0.56 |
| Hypoxaemia | 6.01 (4.67-7.32) | 8.93 (7.75-10.10) | 0.52 (0.42-0.64) | <0.0001 |
| ICU admission | 2.83 (1.90-3.75) | 4.53 (3.61-5.43) | 0.50 (0.38-0.68) | <0.0001 |
| Interstitial lung disease | 2.42 (1.76-3.08) | 4.38 (3.43-5.32) | 0.55 (0.40-0.75) | 0.0002 |
| Intubation/Ventilation | 1.87 (1.11-2.62) | 3.02 (2.32-3.73) | 0.48 (0.33-0.69) | <0.0001 |
| Ischaemic stroke | 2.13 (1.46-2.79) | 3.42 (2.52-4.31) | 0.62 (0.44-0.89) | 0.008 |
| Joint pain | 18.92 (16.13-21.63) | 17.94 (15.50-20.31) | 1.01 (0.86-1.18) | 0.93 |
| Kidney disease | 8.45 (7.23- 9.66) | 10.50 (8.97-12.00) | 0.86 (0.71-1.02) | 0.09 |
| Liver disease | 8.42 (6.73-10.08) | 7.99 (6.65-9.31) | 0.93 (0.75-1.15) | 0.5 |
| Long COVID feature (any) | 62.45 (59.19-65.46) | 61.90 (58.90-64.67) | 0.97 (0.90-1.05) | 0.45 |
| Mood disorder | 21.85 (19.19-24.43) | 18.68 (16.28-21.01) | 1.15 (1.00-1.33) | 0.04 |
| Myalgia | 4.45 (3.23-5.66) | 5.58 (4.47-6.68) | 0.65 (0.49-0.86) | 0.002 |
| Myocarditis | 1.42 (0.87-1.98) | 2.41 (1.74-3.08) | 0.49 (0.32-0.76) | 0.001 |
| Myoneural junction/muscle disease | 1.68 (1.10-2.27) | 2.70 (2.00-3.39) | 0.55 (0.37-0.82) | 0.003 |
| Nerve/nerve root/plexus disorder | 5.00 (3.64-6.34) | 6.17 (4.89-7.43) | 0.68 (0.52-0.89) | 0.005 |
| Obesity | 40.91 (37.97-43.71) | 43.94 (40.91-46.82) | 0.95 (0.87-1.05) | 0.32 |
| Other pain | 14.96 (12.77-17.09) | 14.75 (12.69-16.77) | 0.97 (0.82-1.14) | 0.71 |
| Oxygen requirement | 6.75 (5.36-8.12) | 8.09 (6.91-9.26) | 0.67 (0.54-0.82) | <0.0001 |
| Peripheral neuropathy | 4.22 (3.05-5.38) | 4.12 (3.09-5.14) | 0.86 (0.63-1.18) | 0.34 |
| Psychotic disorder | 2.07 (1.28-2.85) | 3.47 (2.63-4.30) | 0.50 (0.35-0.72) | 0.0001 |
| Respiratory failure | 4.85 (3.78-5.91) | 8.26 (7.16-9.35) | 0.48 (0.39-0.60) | <0.0001 |
| Seizures | 2.41 (1.55-3.27) | 4.09 (3.21-4.95) | 0.45 (0.32-0.64) | <0.0001 |

| | | | | |
|---|---|---|---|---|
| Sleep disorders | 20.24 (17.65-22.75) | 16.44 (14.34-18.48) | 1.06 (0.92-1.23) | 0.41 |
| Type 2 diabetes mellitus | 19.11 (16.92-21.23) | 18.24 (16.24-20.19) | 1.00 (0.87-1.14) | 0.98 |
| Urticaria | 1.85 (1.13-2.57) | 2.84 (2.07-3.60) | 0.51 (0.34-0.77) | 0.0009 |

**Table S12** – Percent incidences and HRs (95% CI) for all outcomes (each as part of a composite endpoint with death as the other component) in the matched over-60 vaccinated and unvaccinated cohorts (*n*=4657 individuals in each cohort).

| Outcome | Incidence % (95% CI) in vaccinated cohort | Incidence % (95% CI) in unvaccinated cohort | HR (95% CI) | p-value |
|---|---|---|---|---|
| Abdominal symptoms | 20.55 (18.69-22.37) | 20.14 (18.15-22.08) | 1.13 (1.00-1.27) | 0.05 |
| Abnormal breathing | 28.86 (26.75-30.92) | 29.70 (27.46-31.87) | 1.01 (0.91-1.11) | 0.86 |
| Anosmia | 5.99 (5.06-6.91) | 6.93 (5.80-8.06) | 0.93 (0.76-1.13) | 0.45 |
| Anxiety disorder | 24.22 (22.07-26.30) | 22.26 (20.23-24.23) | 1.11 (0.99-1.25) | 0.07 |
| Anxiety/Depression | 33.16 (30.81-35.43) | 30.76 (28.48-32.97) | 1.12 (1.02-1.23) | 0.02 |
| Arrhythmia | 28.63 (26.59-30.61) | 28.61 (26.52-30.65) | 1.04 (0.94-1.14) | 0.46 |
| Cardiac failure | 20.44 (18.63-22.21) | 19.33 (17.61-21.00) | 1.09 (0.97-1.22) | 0.15 |
| Cardiomyopathy | 11.26 ( 9.80-12.70) | 12.33 (10.81-13.82) | 0.96 (0.82-1.12) | 0.6 |
| Cerebral haemorrhage | 6.53 (5.55-7.50) | 7.42 (6.20-8.61) | 0.97 (0.80-1.18) | 0.77 |
| Chest/Throat pain | 18.57 (16.64-20.45) | 18.41 (16.55-20.23) | 1.04 (0.91-1.18) | 0.58 |
| Cognitive symptoms | 17.50 (15.81-19.17) | 17.75 (16.08-19.39) | 1.00 (0.89-1.14) | 0.96 |
| Coronary disease | 11.98 (10.44-13.50) | 10.51 (9.11-11.89) | 1.10 (0.94-1.29) | 0.22 |
| Death | 5.68 (4.78-6.57) | 6.68 (5.56-7.79) | 0.91 (0.74-1.12) | 0.38 |
| Fatigue | 23.59 (21.53-25.60) | 24.61 (22.53-26.63) | 0.92 (0.83-1.03) | 0.14 |
| GORD | 29.99 (27.75-32.15) | 30.57 (28.30-32.77) | 1.02 (0.93-1.13) | 0.64 |
| Hair loss | 7.14 (6.04-8.23) | 7.99 (6.76-9.20) | 0.91 (0.75-1.10) | 0.32 |
| Headache | 14.51 (12.87-16.11) | 15.52 (13.73-17.28) | 0.98 (0.85-1.13) | 0.75 |
| Hospitalisation | 24.91 (23.24-26.54) | 25.14 (23.42-26.82) | 1.03 (0.94-1.13) | 0.51 |
| Hypercoagulopathy/DVT/PE | 13.43 (11.97-14.87) | 15.14 (13.55-16.70) | 0.93 (0.81-1.07) | 0.3 |
| Hyperlipidaemia | 58.90 (56.11-61.51) | 55.48 (52.90-57.92) | 1.10 (1.03-1.18) | 0.008 |
| Hypertension | 69.12 (66.35-71.65) | 65.97 (63.37-68.38) | 1.10 (1.03-1.17) | 0.004 |
| Hypoxaemia | 15.33 (13.94-16.69) | 17.77 (16.20-19.31) | 0.90 (0.80-1.02) | 0.09 |
| ICU admission | 9.24 (8.15-10.32) | 10.82 (9.49-12.13) | 0.92 (0.79-1.08) | 0.31 |
| Interstitial lung disease | 9.16 (7.93-10.37) | 10.85 (9.47-12.22) | 0.88 (0.75-1.04) | 0.14 |
| Intubation/Ventilation | 6.36 (5.46-7.25) | 8.19 (7.01-9.35) | 0.85 (0.71-1.03) | 0.09 |
| Ischaemic stroke | 10.33 (8.99-11.64) | 11.13 ( 9.62-12.62) | 0.98 (0.83-1.15) | 0.79 |
| Joint pain | 23.35 (21.16-25.48) | 25.15 (22.81-27.43) | 0.99 (0.88-1.12) | 0.89 |
| Kidney disease | 25.97 (23.97-27.92) | 25.93 (23.97-27.85) | 1.05 (0.95-1.16) | 0.33 |
| Liver disease | 11.81 (10.49-13.11) | 13.20 (11.71-14.66) | 0.95 (0.82-1.10) | 0.48 |
| Long COVID feature (any) | 67.18 (64.67-69.50) | 69.39 (66.89-71.71) | 1.02 (0.95-1.08) | 0.6 |
| Mood disorder | 24.24 (22.03-26.39) | 22.67 (20.64-24.64) | 1.07 (0.95-1.20) | 0.25 |
| Myalgia | 8.78 (7.50-10.05) | 9.50 (8.07-10.90) | 0.95 (0.80-1.14) | 0.61 |
| Myocarditis | 5.78 (4.88-6.68) | 7.05 (5.87-8.21) | 0.90 (0.73-1.10) | 0.3 |
| Myoneural junction/muscle disease | 6.64 (5.66-7.62) | 8.13 (6.92-9.32) | 0.87 (0.72-1.05) | 0.16 |
| Nerve/nerve root/plexus disorder | 10.81 (9.37-12.22) | 10.78 (9.38-12.16) | 0.95 (0.81-1.12) | 0.56 |
| Obesity | 39.35 (36.99-41.62) | 38.65 (36.29-40.92) | 1.03 (0.95-1.12) | 0.46 |
| Other pain | 21.06 (18.97-23.09) | 22.97 (20.90-24.98) | 0.91 (0.81-1.02) | 0.11 |
| Oxygen requirement | 16.21 (14.67-17.71) | 16.38 (14.88-17.85) | 1.02 (0.91-1.15) | 0.71 |
| Peripheral neuropathy | 12.24 (10.74-13.71) | 12.51 (10.88-14.11) | 1.04 (0.89-1.21) | 0.64 |
| Psychotic disorder | 6.28 (5.34-7.21) | 7.94 (6.72-9.15) | 0.86 (0.71-1.05) | 0.14 |
| Respiratory failure | 14.83 (13.43-16.21) | 18.20 (16.61-19.75) | 0.85 (0.76-0.96) | 0.009 |
| Seizures | 7.37 (6.24-8.49) | 8.54 (7.27- 9.79) | 0.88 (0.73-1.06) | 0.18 |

| | | | | |
|---|---|---|---|---|
| Sleep disorders | 25.28 (23.14-27.37) | 25.83 (23.73-27.88) | 1.00 (0.90-1.11) | 0.99 |
| Type 2 diabetes mellitus | 34.83 (32.70-36.90) | 34.51 (32.31-36.64) | 1.07 (0.98-1.17) | 0.14 |
| Urticaria | 6.31 (5.33-7.27) | 7.09 (5.95-8.22) | 0.93 (0.76-1.14) | 0.48 |

**Table S13** – HRs (95% CI) for outcomes (each as part of a composite endpoint with death as the other component) comparing younger (< 60 years-old) vaccinated to younger unvaccinated individuals (*n*=4633 in each group), and older (≥ 60 years-old) vaccinated to older unvaccinated individuals (*n*=4657 in each group), both identical to those presented in Table S11 and S12 respectively, as well as their ratio (95% CI) obtained by fitting an interaction term in the Cox model, and the p-value for the test of the null hypothesis that the latter ratio is equal to 1.

| Outcome | HR (95% CI) in the over-60 cohort | HR (95% CI) in the under-60 cohort | Ratio between HR (95% CI) | p-value |
|---|---|---|---|---|
| Abdominal symptoms | 1.13 (1.00-1.27) | 0.99 (0.85-1.15) | 1.15 (0.95-1.39) | 0.16 |
| Abnormal breathing | 1.01 (0.91-1.11) | 0.88 (0.77-1.00) | 1.15 (0.98-1.35) | 0.096 |
| Anosmia | 0.93 (0.76-1.13) | 0.62 (0.42-0.92) | 1.47 (0.95-2.28) | 0.084 |
| Anxiety disorder | 1.11 (0.99-1.25) | 1.12 (1.00-1.27) | 0.99 (0.83-1.17) | 0.87 |
| Anxiety/Depression | 1.12 (1.02-1.23) | 1.09 (0.97-1.21) | 1.03 (0.89-1.19) | 0.68 |
| Arrhythmia | 1.04 (0.94-1.14) | 0.91 (0.77-1.08) | 1.13 (0.93-1.37) | 0.21 |
| Cardiac failure | 1.09 (0.97-1.22) | 0.85 (0.67-1.08) | 1.28 (0.98-1.67) | 0.066 |
| Cardiomyopathy | 0.96 (0.82-1.12) | 1.00 (0.75-1.32) | 0.96 (0.70-1.33) | 0.82 |
| Cerebral haemorrhage | 0.97 (0.80-1.18) | 0.47 (0.31-0.73) | 2.06 (1.28-3.30) | 0.0029 |
| Chest/Throat pain | 1.04 (0.91-1.18) | 0.91 (0.77-1.08) | 1.14 (0.93-1.41) | 0.21 |
| Cognitive symptoms | 1.00 (0.89-1.14) | 0.82 (0.64-1.06) | 1.22 (0.92-1.61) | 0.17 |
| Coronary disease | 1.10 (0.94-1.29) | 0.60 (0.42-0.86) | 1.81 (1.23-2.66) | 0.0025 |
| Death | 0.91 (0.74-1.12) | 0.43 (0.26-0.68) | 2.13 (1.27-3.58) | 0.0041 |
| Fatigue | 0.92 (0.83-1.03) | 0.96 (0.81-1.13) | 0.97 (0.79-1.19) | 0.76 |
| GORD | 1.02 (0.93-1.13) | 1.08 (0.94-1.25) | 0.95 (0.80-1.13) | 0.57 |
| Hair loss | 0.91 (0.75-1.10) | 0.59 (0.42-0.81) | 1.53 (1.04-2.25) | 0.029 |
| Headache | 0.98 (0.85-1.13) | 1.03 (0.89-1.20) | 0.95 (0.77-1.16) | 0.61 |
| Hospitalisation | 1.03 (0.94-1.13) | 0.81 (0.70-0.94) | 1.27 (1.07-1.51) | 0.0075 |
| Hypercoagulopathy/DVT/PE | 0.93 (0.81-1.07) | 0.72 (0.57-0.90) | 1.30 (1.00-1.70) | 0.054 |
| Hyperlipidaemia | 1.10 (1.03-1.18) | 1.01 (0.90-1.14) | 1.09 (0.95-1.25) | 0.24 |
| Hypertension | 1.10 (1.03-1.17) | 0.97 (0.88-1.08) | 1.13 (1.00-1.28) | 0.045 |
| Hypoxaemia | 0.90 (0.80-1.02) | 0.52 (0.42-0.64) | 1.74 (1.37-2.20) | <0.0001 |
| ICU admission | 0.92 (0.79-1.08) | 0.50 (0.38-0.68) | 1.82 (1.31-2.54) | 0.00042 |
| Interstitial lung disease | 0.88 (0.75-1.04) | 0.55 (0.40-0.75) | 1.63 (1.14-2.33) | 0.0077 |
| Intubation/Ventilation | 0.85 (0.71-1.03) | 0.48 (0.33-0.69) | 1.79 (1.18-2.70) | 0.006 |
| Ischaemic stroke | 0.98 (0.83-1.15) | 0.62 (0.44-0.89) | 1.57 (1.07-2.32) | 0.022 |
| Joint pain | 0.99 (0.88-1.12) | 1.01 (0.86-1.18) | 0.99 (0.81-1.20) | 0.89 |
| Kidney disease | 1.05 (0.95-1.16) | 0.86 (0.71-1.02) | 1.23 (1.00-1.51) | 0.05 |
| Liver disease | 0.95 (0.82-1.10) | 0.93 (0.75-1.15) | 1.02 (0.79-1.32) | 0.89 |
| Long COVID feature (any) | 1.02 (0.95-1.08) | 0.97 (0.90-1.05) | 1.05 (0.95-1.15) | 0.35 |
| Mood disorder | 1.07 (0.95-1.20) | 1.15 (1.00-1.33) | 0.93 (0.78-1.11) | 0.42 |
| Myalgia | 0.95 (0.80-1.14) | 0.65 (0.49-0.86) | 1.48 (1.06-2.07) | 0.023 |
| Myocarditis | 0.90 (0.73-1.10) | 0.49 (0.32-0.76) | 1.79 (1.11-2.90) | 0.017 |
| Myoneural junction/muscle disease | 0.87 (0.72-1.05) | 0.55 (0.37-0.82) | 1.58 (1.01-2.45) | 0.044 |
| Nerve/nerve root/plexus disorder | 0.95 (0.81-1.12) | 0.68 (0.52-0.89) | 1.41 (1.02-1.94) | 0.037 |
| Obesity | 1.03 (0.95-1.12) | 0.95 (0.87-1.05) | 1.08 (0.95-1.22) | 0.22 |
| Other pain | 0.91 (0.81-1.02) | 0.97 (0.82-1.14) | 0.94 (0.76-1.15) | 0.53 |
| Oxygen requirement | 1.02 (0.91-1.15) | 0.67 (0.54-0.82) | 1.54 (1.21-1.95) | 0.00036 |
| Peripheral neuropathy | 1.04 (0.89-1.21) | 0.86 (0.63-1.18) | 1.21 (0.85-1.71) | 0.3 |
| Psychotic disorder | 0.86 (0.71-1.05) | 0.50 (0.35-0.72) | 1.71 (1.13-2.59) | 0.011 |

| | | | | |
|---|---|---|---|---|
| Respiratory failure | 0.85 (0.76-0.96) | 0.48 (0.39-0.60) | 1.77 (1.38-2.26) | <0.0001 |
| Seizures | 0.88 (0.73-1.06) | 0.45 (0.32-0.64) | 1.95 (1.31-2.89) | 0.00092 |
| Sleep disorders | 1.00 (0.90-1.11) | 1.06 (0.92-1.23) | 0.94 (0.79-1.13) | 0.53 |
| Type 2 diabetes mellitus | 1.07 (0.98-1.17) | 1.00 (0.87-1.14) | 1.07 (0.91-1.26) | 0.41 |
| Urticaria | 0.93 (0.76-1.14) | 0.51 (0.34-0.77) | 1.82 (1.16-2.85) | 0.0097 |

**Table S14** – Percent incidences and HRs (95% CI) for all distinct outcomes (i.e. not as part of a composite endpoint with death as the other component, and thus subject to survivorship bias) in the matched vaccinated and unvaccinated cohorts (*n*=9479 individuals in each cohort).

| Outcome | Incidence % (95% CI) in vaccinated cohort | Incidence % (95% CI) in unvaccinated cohort | HR (95% CI) | p-value |
|---|---|---|---|---|
| Abdominal symptoms | 16.97 (15.49-18.43) | 15.94 (14.50-17.35) | 1.16 (1.04-1.28) | 0.008 |
| Abnormal breathing | 23.29 (21.72-24.83) | 22.50 (21.01-23.97) | 1.01 (0.93-1.10) | 0.78 |
| Anosmia | 0.65 (0.36-0.94) | 0.24 (0.11-0.37) | 2.26 (1.20-4.27) | 0.009 |
| Anxiety disorder | 22.08 (20.50-23.63) | 20.08 (18.57-21.57) | 1.13 (1.03-1.24) | 0.008 |
| Anxiety/Depression | 29.95 (28.22-31.64) | 27.27 (25.60-28.89) | 1.13 (1.05-1.22) | 0.001 |
| Arrhythmia | 19.79 (18.38-21.19) | 19.48 (18.16-20.77) | 0.99 (0.91-1.08) | 0.77 |
| Cardiac failure | 11.27 (10.12-12.40) | 10.39 (9.41-11.36) | 1.06 (0.95-1.20) | 0.3 |
| Cardiomyopathy | 4.72 (3.99-5.45) | 4.11 (3.46-4.77) | 1.18 (0.97-1.43) | 0.1 |
| Cerebral haemorrhage | 0.59 (0.39-0.79) | 0.51 (0.30-0.72) | 1.33 (0.81-2.16) | 0.25 |
| Chest/Throat pain | 12.90 (11.53-14.25) | 12.31 (11.08-13.53) | 1.08 (0.96-1.21) | 0.21 |
| Cognitive symptoms | 9.67 (8.58-10.75) | 9.65 (8.64-10.65) | 1.02 (0.90-1.16) | 0.77 |
| Coronary disease | 4.52 (3.68-5.34) | 3.80 (3.13-4.46) | 1.04 (0.86-1.27) | 0.67 |
| Death | 3.74 (3.12-4.35) | 4.50 (3.89-5.10) | 0.75 (0.62-0.91) | 0.003 |
| Fatigue | 17.56 (15.99-19.10) | 16.89 (15.50-18.26) | 0.96 (0.86-1.06) | 0.38 |
| GORD | 23.41 (21.74-25.03) | 21.60 (20.04-23.12) | 1.10 (1.01-1.20) | 0.03 |
| Hair loss | 1.66 (1.18-2.14) | 2.30 (1.73-2.87) | 0.77 (0.55-1.06) | 0.11 |
| Headache | 12.54 (11.27-13.79) | 10.26 (9.13-11.37) | 1.25 (1.10-1.42) | 0.0004 |
| Hospitalisation | 17.01 (15.90-18.10) | 17.64 (16.56-18.71) | 0.94 (0.87-1.02) | 0.14 |
| Hypercoagulopathy/DVT/PE | 7.07 (6.21-7.93) | 8.43 (7.51-9.34) | 0.86 (0.75-0.99) | 0.04 |
| Hyperlipidaemia | 42.67 (40.62-44.64) | 41.48 (39.56-43.33) | 1.05 (0.99-1.12) | 0.12 |
| Hypertension | 52.28 (50.16-54.31) | 50.89 (48.98-52.73) | 1.00 (0.95-1.06) | 0.96 |
| Hypoxaemia | 8.99 (8.10- 9.86) | 11.87 (10.97-12.76) | 0.72 (0.65-0.81) | <0.0001 |
| ICU admission | 4.32 (3.77-4.87) | 5.25 (4.62-5.87) | 0.82 (0.70-0.96) | 0.01 |
| Interstitial lung disease | 2.85 (2.30-3.39) | 3.06 (2.45-3.67) | 1.03 (0.82-1.30) | 0.81 |
| Intubation/Ventilation | 2.12 (1.76-2.49) | 2.59 (2.18-2.99) | 0.81 (0.65-1.01) | 0.06 |
| Ischaemic stroke | 3.28 (2.65-3.90) | 3.21 (2.57-3.85) | 1.01 (0.80-1.26) | 0.94 |
| Joint pain | 17.84 (16.20-19.44) | 16.70 (15.19-18.19) | 1.08 (0.97-1.21) | 0.17 |
| Kidney disease | 16.51 (15.24-17.76) | 17.07 (15.83-18.29) | 1.01 (0.92-1.11) | 0.86 |
| Liver disease | 6.87 (6.02-7.71) | 6.73 (5.87-7.58) | 1.08 (0.93-1.26) | 0.3 |
| Long COVID feature (any) | 63.37 (61.43-65.22) | 64.51 (62.57-66.34) | 1.02 (0.97-1.07) | 0.43 |
| Mood disorder | 19.50 (17.95-21.03) | 16.72 (15.38-18.04) | 1.16 (1.05-1.27) | 0.004 |
| Myalgia | 3.27 (2.53-3.99) | 3.67 (2.95-4.40) | 0.85 (0.67-1.08) | 0.18 |
| Myocarditis | 0.21 (0.07-0.36) | 0.22 (0.04-0.40) | 1.15 (0.49-2.66) | 0.75 |
| Myoneural junction/muscle disease | 0.77 (0.53-1.01) | 0.97 (0.63-1.32) | 0.96 (0.63-1.44) | 0.83 |
| Nerve/nerve root/plexus disorder | 4.61 (3.77-5.45) | 4.42 (3.64-5.19) | 0.92 (0.75-1.13) | 0.43 |
| Obesity | 37.58 (35.76-39.36) | 37.58 (35.79-39.32) | 1.00 (0.93-1.06) | 0.93 |
| Other pain | 15.04 (13.60-16.46) | 16.26 (14.79-17.71) | 0.95 (0.85-1.06) | 0.39 |
| Oxygen requirement | 10.12 (9.14-11.10) | 11.76 (10.76-12.75) | 0.86 (0.77-0.95) | 0.005 |
| Peripheral neuropathy | 5.10 (4.24-5.94) | 4.68 (3.81-5.53) | 1.19 (0.97-1.46) | 0.1 |
| Psychotic disorder | 0.73 (0.50-0.95) | 1.11 (0.76-1.45) | 0.79 (0.53-1.18) | 0.24 |
| Respiratory failure | 8.39 (7.55-9.22) | 11.75 (10.81-12.67) | 0.71 (0.64-0.79) | <0.0001 |
| Seizures | 1.51 (1.02-1.99) | 1.77 (1.34-2.19) | 0.70 (0.51-0.97) | 0.03 |

| | | | | |
|---|---|---|---|---|
| Sleep disorders | 20.03 (18.41-21.61) | 19.44 (17.96-20.89) | 1.02 (0.92-1.12) | 0.74 |
| Type 2 diabetes mellitus | 25.12 (23.61-26.60) | 24.11 (22.70-25.49) | 1.02 (0.94-1.10) | 0.68 |
| Urticaria | 0.55 (0.31-0.79) | 0.50 (0.31-0.70) | 0.97 (0.57-1.64) | 0.9 |

**Table S15** – Percent incidences and HRs (95% CI) for all distinct outcomes (i.e. not as part of a composite endpoint with death as the other component, and thus subject to survivorship bias) in the matched vaccinated (1 dose only) and unvaccinated cohorts (*n*=2996 individuals in each cohort).

| Outcome | Incidence % (95% CI) in vaccinated cohort | Incidence % (95% CI) in unvaccinated cohort | HR (95% CI) | p-value |
|---|---|---|---|---|
| Abdominal symptoms | 16.12 (14.16-18.04) | 16.25 (14.25-18.19) | 0.97 (0.82-1.14) | 0.71 |
| Abnormal breathing | 22.35 (20.29-24.36) | 21.32 (19.24-23.35) | 1.04 (0.92-1.19) | 0.51 |
| Anosmia | 0.77 (0.37-1.17) | 0.55 (0.24-0.85) | 1.23 (0.60-2.52) | 0.58 |
| Anxiety disorder | 22.01 (19.83-24.12) | 22.35 (20.13-24.51) | 0.98 (0.85-1.12) | 0.73 |
| Anxiety/Depression | 28.34 (26.01-30.60) | 28.33 (26.02-30.57) | 0.99 (0.88-1.11) | 0.86 |
| Arrhythmia | 18.89 (17.04-20.71) | 18.56 (16.68-20.40) | 1.01 (0.88-1.17) | 0.85 |
| Cardiac failure | 10.55 (9.18-11.90) | 10.44 (8.99-11.86) | 1.03 (0.86-1.24) | 0.72 |
| Cardiomyopathy | 4.17 (3.26-5.07) | 3.58 (2.74-4.41) | 1.16 (0.85-1.57) | 0.35 |
| Cerebral haemorrhage | 1.04 (0.55-1.54) | 0.36 (0.12-0.60) | 2.06 (0.94-4.53) | 0.06 |
| Chest/Throat pain | 12.20 (10.41-13.96) | 12.04 (10.32-13.72) | 1.00 (0.83-1.20) | 1 |
| Cognitive symptoms | 8.73 (7.42-10.03) | 10.36 (8.94-11.76) | 0.81 (0.67-0.99) | 0.04 |
| Coronary disease | 3.56 (2.69-4.42) | 3.14 (2.31-3.97) | 1.13 (0.81-1.58) | 0.47 |
| Death | 2.95 (2.26-3.63) | 4.73 (3.70-5.74) | 0.72 (0.53-0.98) | 0.03 |
| Fatigue | 15.89 (13.91-17.82) | 16.27 (14.46-18.05) | 0.88 (0.75-1.04) | 0.13 |
| GORD | 22.76 (20.54-24.92) | 22.87 (20.58-25.09) | 1.01 (0.88-1.16) | 0.88 |
| Hair loss | 1.47 (0.92-2.01) | 1.91 (1.25-2.56) | 0.74 (0.46-1.21) | 0.23 |
| Headache | 13.20 (11.43-14.93) | 11.75 ( 9.94-13.53) | 1.25 (1.04-1.51) | 0.02 |
| Hospitalisation | 17.56 (15.99-19.10) | 18.52 (16.81-20.19) | 0.97 (0.85-1.10) | 0.63 |
| Hypercoagulopathy/DVT/PE | 6.40 (5.33-7.47) | 7.20 (6.00-8.39) | 0.94 (0.75-1.18) | 0.61 |
| Hyperlipidaemia | 41.06 (38.31-43.67) | 41.83 (38.95-44.58) | 1.01 (0.91-1.11) | 0.88 |
| Hypertension | 50.02 (47.27-52.61) | 51.70 (48.71-54.51) | 1.00 (0.92-1.10) | 0.93 |
| Hypoxaemia | 8.78 (7.62- 9.92) | 10.91 ( 9.58-12.22) | 0.80 (0.67-0.96) | 0.02 |
| ICU admission | 3.89 (3.13-4.64) | 5.75 (4.70-6.78) | 0.74 (0.57-0.95) | 0.02 |
| Interstitial lung disease | 2.31 (1.67-2.94) | 2.96 (2.20-3.72) | 0.81 (0.56-1.16) | 0.24 |
| Intubation/Ventilation | 1.82 (1.30-2.33) | 2.40 (1.79-3.01) | 0.77 (0.53-1.12) | 0.17 |
| Ischaemic stroke | 3.39 (2.52-4.25) | 2.21 (1.54-2.88) | 1.49 (1.04-2.14) | 0.03 |
| Joint pain | 17.05 (14.90-19.14) | 17.45 (15.27-19.57) | 1.00 (0.85-1.18) | 0.99 |
| Kidney disease | 14.96 (13.28-16.60) | 17.14 (15.27-18.96) | 0.90 (0.78-1.05) | 0.19 |
| Liver disease | 6.57 (5.40-7.73) | 7.23 (5.98-8.46) | 0.91 (0.72-1.15) | 0.45 |
| Long COVID feature (any) | 63.70 (60.88-66.32) | 65.18 (62.35-67.79) | 0.96 (0.89-1.04) | 0.3 |
| Mood disorder | 17.64 (15.63-19.61) | 17.47 (15.56-19.34) | 0.98 (0.84-1.14) | 0.82 |
| Myalgia | 2.40 (1.61-3.19) | 3.40 (2.46-4.33) | 0.69 (0.46-1.02) | 0.06 |
| Myocarditis | 0.30 (0.07-0.52) | 0.45 (0.04-0.86) | 1.30 (0.41-4.09) | 0.66 |
| Myoneural junction/muscle disease | 0.66 (0.31-1.01) | 0.73 (0.38-1.09) | 0.83 (0.41-1.66) | 0.6 |
| Nerve/nerve root/plexus disorder | 4.34 (3.24-5.42) | 4.20 (3.22-5.16) | 0.91 (0.66-1.25) | 0.55 |
| Obesity | 40.84 (38.21-43.36) | 41.02 (38.31-43.61) | 1.02 (0.93-1.13) | 0.63 |
| Other pain | 15.06 (13.10-16.99) | 16.06 (14.07-17.99) | 0.86 (0.73-1.02) | 0.08 |
| Oxygen requirement | 10.32 (8.95-11.67) | 11.25 ( 9.89-12.60) | 0.90 (0.76-1.07) | 0.23 |
| Peripheral neuropathy | 4.84 (3.82-5.84) | 3.97 (3.01-4.92) | 1.27 (0.94-1.72) | 0.12 |
| Psychotic disorder | 0.88 (0.52-1.24) | 2.02 (1.37-2.67) | 0.50 (0.30-0.84) | 0.007 |
| Respiratory failure | 7.85 (6.76-8.93) | 10.14 (8.85-11.40) | 0.79 (0.65-0.95) | 0.01 |
| Seizures | 1.49 (0.91-2.06) | 2.10 (1.50-2.70) | 0.59 (0.38-0.93) | 0.02 |

| | | | | |
|---|---|---|---|---|
| Sleep disorders | 19.05 (16.98-21.06) | 18.66 (16.57-20.70) | 1.01 (0.87-1.17) | 0.95 |
| Type 2 diabetes mellitus | 23.41 (21.43-25.35) | 24.05 (21.93-26.12) | 0.98 (0.87-1.11) | 0.79 |
| Urticaria | 0.67 (0.29-1.06) | 0.47 (0.18-0.75) | 1.25 (0.56-2.79) | 0.59 |

**Table S16** – Percent incidences and HRs (95% CI) for all distinct outcomes (i.e. not as part of a composite endpoint with death as the other component, and thus subject to survivorship bias) in the matched vaccinated (2 doses only) and unvaccinated cohorts (*n*=6957 individuals in each cohort).

| Outcome | Incidence % (95% CI) in vaccinated cohort | Incidence % (95% CI) in unvaccinated cohort | HR (95% CI) | p-value |
|---|---|---|---|---|
| Abdominal symptoms | 17.31 (15.37-19.21) | 16.19 (14.25-18.10) | 1.21 (1.06-1.37) | 0.004 |
| Abnormal breathing | 22.45 (20.51-24.35) | 22.27 (20.39-24.10) | 1.00 (0.91-1.11) | 0.96 |
| Anosmia | 0.89 (0.44-1.35) | 0.37 (0.17-0.58) | 1.79 (0.95-3.37) | 0.06 |
| Anxiety disorder | 20.36 (18.49-22.20) | 22.58 (20.39-24.72) | 1.09 (0.98-1.22) | 0.1 |
| Anxiety/Depression | 28.70 (26.42-30.91) | 30.82 (28.33-33.22) | 1.09 (0.99-1.19) | 0.07 |
| Arrhythmia | 18.62 (17.04-20.16) | 21.27 (19.37-23.12) | 0.95 (0.86-1.05) | 0.34 |
| Cardiac failure | 11.07 ( 9.63-12.49) | 10.79 (9.40-12.16) | 1.06 (0.92-1.22) | 0.42 |
| Cardiomyopathy | 5.37 (4.31-6.42) | 4.11 (3.28-4.93) | 1.19 (0.95-1.50) | 0.13 |
| Cerebral haemorrhage | 0.48 (0.27-0.68) | 0.28 (0.13-0.43) | 1.71 (0.88-3.33) | 0.11 |
| Chest/Throat pain | 13.22 (11.26-15.14) | 12.94 (11.26-14.59) | 1.09 (0.94-1.26) | 0.24 |
| Cognitive symptoms | 9.66 (8.18-11.11) | 9.56 (8.36-10.75) | 1.00 (0.86-1.17) | 0.96 |
| Coronary disease | 5.72 (4.33-7.08) | 3.82 (2.93-4.69) | 1.17 (0.92-1.48) | 0.2 |
| Death | 3.35 (2.66-4.03) | 4.98 (4.21-5.74) | 0.62 (0.49-0.78) | <0.0001 |
| Fatigue | 17.51 (15.50-19.48) | 16.80 (15.10-18.47) | 0.96 (0.85-1.08) | 0.51 |
| GORD | 24.58 (22.32-26.78) | 21.22 (19.19-23.20) | 1.17 (1.05-1.30) | 0.004 |
| Hair loss | 1.17 (0.68-1.66) | 0.90 (0.52-1.28) | 1.23 (0.74-2.06) | 0.42 |
| Headache | 13.18 (11.55-14.78) | 11.96 (10.22-13.66) | 1.19 (1.03-1.38) | 0.02 |
| Hospitalisation | 17.64 (16.01-19.22) | 18.14 (16.67-19.59) | 0.97 (0.88-1.06) | 0.51 |
| Hypercoagulopathy/DVT/PE | 6.97 (5.90-8.02) | 7.30 (6.16-8.43) | 0.98 (0.82-1.17) | 0.83 |
| Hyperlipidaemia | 47.58 (44.48-50.51) | 44.09 (41.37-46.67) | 1.09 (1.01-1.18) | 0.02 |
| Hypertension | 56.45 (53.23-59.45) | 49.49 (46.99-51.88) | 1.11 (1.04-1.19) | 0.002 |
| Hypoxaemia | 8.89 (7.63-10.14) | 12.24 (11.02-13.45) | 0.69 (0.60-0.79) | <0.0001 |
| ICU admission | 4.59 (3.83-5.34) | 6.21 (5.22-7.18) | 0.80 (0.67-0.97) | 0.02 |
| Interstitial lung disease | 2.81 (2.06-3.55) | 3.68 (2.79-4.56) | 0.92 (0.70-1.20) | 0.53 |
| Intubation/Ventilation | 2.21 (1.77-2.65) | 2.96 (2.31-3.60) | 0.85 (0.65-1.10) | 0.21 |
| Ischaemic stroke | 3.32 (2.50-4.12) | 3.58 (2.76-4.38) | 0.96 (0.74-1.25) | 0.77 |
| Joint pain | 18.04 (15.81-20.22) | 19.70 (17.39-21.95) | 0.99 (0.87-1.13) | 0.85 |
| Kidney disease | 17.56 (15.83-19.25) | 18.01 (16.14-19.84) | 1.06 (0.95-1.18) | 0.31 |
| Liver disease | 7.76 (6.56-8.96) | 7.56 (6.30-8.80) | 1.20 (1.01-1.44) | 0.04 |
| Long COVID feature (any) | 64.10 (61.44-66.57) | 64.85 (62.18-67.33) | 1.04 (0.98-1.10) | 0.2 |
| Mood disorder | 19.23 (17.15-21.25) | 18.07 (16.12-19.97) | 1.14 (1.02-1.29) | 0.03 |
| Myalgia | 3.19 (2.32-4.05) | 3.79 (2.90-4.66) | 0.89 (0.68-1.19) | 0.44 |
| Myocarditis | 0.13 (0.03-0.23) | 0.15 (0.05-0.25) | 0.94 (0.34-2.60) | 0.91 |
| Myoneural junction/muscle disease | 0.79 (0.51-1.08) | 1.19 (0.77-1.60) | 0.80 (0.50-1.26) | 0.33 |
| Nerve/nerve root/plexus disorder | 4.18 (3.18-5.18) | 4.72 (3.52-5.89) | 0.96 (0.74-1.23) | 0.73 |
| Obesity | 34.90 (32.59-37.13) | 35.33 (32.94-37.64) | 1.01 (0.93-1.10) | 0.74 |
| Other pain | 15.13 (13.27-16.94) | 17.58 (15.39-19.72) | 0.96 (0.84-1.10) | 0.59 |
| Oxygen requirement | 9.33 (8.23-10.42) | 10.90 ( 9.85-11.94) | 0.81 (0.71-0.92) | 0.001 |
| Peripheral neuropathy | 4.83 (3.85-5.80) | 5.57 (4.33-6.80) | 1.03 (0.81-1.31) | 0.82 |
| Psychotic disorder | 0.70 (0.39-1.01) | 0.77 (0.50-1.03) | 0.70 (0.42-1.18) | 0.18 |
| Respiratory failure | 8.51 (7.30- 9.70) | 12.43 (11.16-13.67) | 0.65 (0.57-0.75) | <0.0001 |
| Seizures | 1.78 (1.04-2.53) | 1.32 (0.92-1.72) | 0.97 (0.65-1.43) | 0.87 |

| | | | | |
|---|---|---|---|---|
| Sleep disorders | 19.96 (17.75-22.10) | 19.11 (17.18-21.00) | 1.07 (0.95-1.20) | 0.28 |
| Type 2 diabetes mellitus | 26.51 (24.44-28.52) | 24.90 (22.96-26.78) | 1.11 (1.01-1.21) | 0.03 |
| Urticaria | 0.51 (0.25-0.78) | 0.74 (0.43-1.06) | 0.68 (0.38-1.22) | 0.19 |

**Table S17** – Percent incidences and HRs (95% CI) for all distinct outcomes (i.e. not as part of a composite endpoint with death as the other component, and thus subject to survivorship bias) in the matched under-60 vaccinated and unvaccinated cohorts (*n*=4633 individuals in each cohort).

| Outcome | Incidence % (95% CI) in vaccinated cohort | Incidence % (95% CI) in unvaccinated cohort | HR (95% CI) | p-value |
|---|---|---|---|---|
| Abdominal symptoms | 18.93 (16.20-21.58) | 17.18 (14.70-19.59) | 1.12 (0.95-1.31) | 0.17 |
| Abnormal breathing | 20.19 (17.80-22.51) | 18.72 (16.68-20.71) | 0.98 (0.86-1.12) | 0.82 |
| Anosmia | 1.23 (0.59-1.86) | 0.68 (0.35-1.02) | 1.38 (0.73-2.58) | 0.32 |
| Anxiety disorder | 25.90 (23.27-28.45) | 23.09 (20.55-25.56) | 1.16 (1.03-1.32) | 0.02 |
| Anxiety/Depression | 31.83 (29.09-34.48) | 30.57 (27.73-33.30) | 1.10 (0.98-1.22) | 0.1 |
| Arrhythmia | 11.97 (10.19-13.71) | 11.31 ( 9.69-12.90) | 0.97 (0.81-1.15) | 0.7 |
| Cardiac failure | 4.28 (3.30-5.25) | 4.67 (3.74-5.59) | 0.93 (0.72-1.22) | 0.61 |
| Cardiomyopathy | 3.87 (2.63-5.10) | 2.46 (1.59-3.31) | 1.49 (1.05-2.13) | 0.02 |
| Cerebral haemorrhage | 0.26 (0.07-0.44) | 1.02 (0.49-1.55) | 0.43 (0.19-0.99) | 0.04 |
| Chest/Throat pain | 13.45 (10.92-15.91) | 11.88 (10.08-13.65) | 1.04 (0.87-1.24) | 0.68 |
| Cognitive symptoms | 4.22 (3.18-5.25) | 5.24 (3.97-6.50) | 0.88 (0.66-1.16) | 0.36 |
| Coronary disease | 1.41 (0.64-2.17) | 2.07 (1.22-2.92) | 0.63 (0.39-1.01) | 0.05 |
| Death | 1.17 (0.67-1.68) | 2.25 (1.60-2.89) | 0.43 (0.26-0.68) | 0.0002 |
| Fatigue | 14.15 (11.78-16.46) | 12.85 (10.92-14.74) | 0.95 (0.80-1.14) | 0.6 |
| GORD | 20.94 (18.20-23.59) | 18.99 (16.39-21.50) | 1.14 (0.98-1.32) | 0.09 |
| Hair loss | 1.66 (1.05-2.26) | 2.24 (1.41-3.06) | 0.95 (0.60-1.53) | 0.84 |
| Headache | 17.05 (14.66-19.37) | 15.25 (13.21-17.24) | 1.12 (0.96-1.31) | 0.16 |
| Hospitalisation | 9.59 (8.21-10.96) | 11.49 (10.23-12.74) | 0.79 (0.68-0.92) | 0.002 |
| Hypercoagulopathy/DVT/PE | 4.60 (3.50-5.70) | 6.51 (5.25-7.75) | 0.71 (0.55-0.91) | 0.007 |
| Hyperlipidaemia | 26.18 (23.52-28.75) | 27.92 (24.77-30.94) | 1.08 (0.95-1.22) | 0.24 |
| Hypertension | 35.04 (31.92-38.02) | 33.30 (30.52-35.98) | 1.00 (0.90-1.11) | 0.95 |
| Hypoxaemia | 4.95 (3.77-6.12) | 7.50 (6.38-8.61) | 0.53 (0.43-0.66) | <0.0001 |
| ICU admission | 1.90 (1.33-2.45) | 3.33 (2.61-4.03) | 0.50 (0.36-0.69) | <0.0001 |
| Interstitial lung disease | 1.54 (1.03-2.05) | 2.19 (1.54-2.84) | 0.72 (0.48-1.08) | 0.11 |
| Intubation/Ventilation | 0.95 (0.59-1.32) | 2.07 (1.58-2.56) | 0.41 (0.27-0.63) | <0.0001 |
| Ischaemic stroke | 1.11 (0.68-1.54) | 1.42 (0.76-2.07) | 1.05 (0.64-1.72) | 0.86 |
| Joint pain | 17.18 (14.57-19.71) | 15.51 (13.05-17.89) | 1.12 (0.94-1.33) | 0.19 |
| Kidney disease | 7.98 (6.79-9.16) | 9.97 (8.45-11.47) | 0.86 (0.71-1.03) | 0.1 |
| Liver disease | 7.54 (5.89-9.16) | 5.86 (4.62-7.08) | 1.14 (0.90-1.45) | 0.27 |
| Long COVID feature (any) | 62.16 (58.89-65.17) | 63.87 (60.57-66.88) | 0.98 (0.91-1.05) | 0.51 |
| Mood disorder | 20.64 (18.06-23.15) | 18.48 (16.10-20.79) | 1.17 (1.02-1.35) | 0.03 |
| Myalgia | 3.39 (2.28-4.50) | 2.96 (2.10-3.82) | 0.96 (0.67-1.38) | 0.84 |
| Myocarditis | 0.29 (0.05-0.52) | 0.05 (0.00-0.11) | 3.81 (0.79-18.39) | 0.07 |
| Myoneural junction/muscle disease | 0.46 (0.21-0.70) | 0.87 (0.49-1.24) | 0.60 (0.31-1.18) | 0.13 |
| Nerve/nerve root/plexus disorder | 4.10 (2.73-5.45) | 2.92 (1.96-3.87) | 1.17 (0.83-1.67) | 0.37 |
| Obesity | 40.48 (37.53-43.30) | 41.95 (39.06-44.70) | 0.94 (0.86-1.03) | 0.21 |
| Other pain | 14.24 (12.07-16.35) | 12.59 (10.47-14.66) | 1.13 (0.95-1.35) | 0.17 |
| Oxygen requirement | 5.64 (4.55-6.71) | 7.17 (6.05-8.28) | 0.65 (0.52-0.80) | <0.0001 |
| Peripheral neuropathy | 2.77 (1.87-3.66) | 2.16 (1.29-3.03) | 1.28 (0.84-1.94) | 0.25 |
| Psychotic disorder | 0.82 (0.46-1.17) | 0.99 (0.51-1.46) | 1.02 (0.57-1.81) | 0.95 |
| Respiratory failure | 3.83 (3.02-4.65) | 7.35 (6.36-8.33) | 0.46 (0.37-0.58) | <0.0001 |
| Seizures | 1.04 (0.58-1.50) | 1.90 (1.26-2.54) | 0.50 (0.31-0.82) | 0.005 |

| | | | | |
|---|---|---|---|---|
| Sleep disorders | 19.57 (16.88-22.18) | 16.70 (14.48-18.87) | 1.06 (0.91-1.24) | 0.45 |
| Type 2 diabetes mellitus | 18.37 (16.22-20.46) | 18.95 (16.80-21.04) | 1.00 (0.87-1.15) | 0.99 |
| Urticaria | 0.61 (0.26-0.97) | 0.65 (0.33-0.98) | 0.77 (0.39-1.54) | 0.46 |

**Table S18** – Percent incidences and HRs (95% CI) for all distinct outcomes (i.e. not as part of a composite endpoint with death as the other component, and thus subject to survivorship bias) in the matched over-60 vaccinated and unvaccinated cohorts (*n*=4657 individuals in each cohort).

| Outcome | Incidence % (95% CI) in vaccinated cohort | Incidence % (95% CI) in unvaccinated cohort | HR (95% CI) | p-value |
|---|---|---|---|---|
| Abdominal symptoms | 15.89 (14.13-17.61) | 14.42 (12.58-16.21) | 1.27 (1.10-1.47) | 0.001 |
| Abnormal breathing | 25.56 (23.48-27.60) | 25.38 (23.20-27.49) | 1.05 (0.94-1.16) | 0.42 |
| Anosmia | 0.26 (0.08-0.45) | 0.24 (0.07-0.40) | 1.07 (0.41-2.78) | 0.89 |
| Anxiety disorder | 19.43 (17.37-21.44) | 16.56 (14.74-18.35) | 1.20 (1.05-1.37) | 0.009 |
| Anxiety/Depression | 29.24 (26.87-31.54) | 25.87 (23.69-27.99) | 1.17 (1.05-1.30) | 0.003 |
| Arrhythmia | 25.84 (23.82-27.81) | 24.78 (22.78-26.73) | 1.07 (0.97-1.19) | 0.18 |
| Cardiac failure | 16.72 (14.97-18.43) | 14.64 (13.09-16.17) | 1.16 (1.02-1.33) | 0.02 |
| Cardiomyopathy | 5.76 (4.67-6.85) | 5.98 (4.89-7.05) | 1.02 (0.81-1.28) | 0.88 |
| Cerebral haemorrhage | 0.86 (0.53-1.19) | 0.88 (0.46-1.30) | 1.33 (0.77-2.31) | 0.31 |
| Chest/Throat pain | 13.43 (11.62-15.21) | 12.79 (11.13-14.41) | 1.08 (0.92-1.26) | 0.33 |
| Cognitive symptoms | 13.54 (11.98-15.07) | 12.95 (11.48-14.41) | 1.06 (0.92-1.23) | 0.4 |
| Coronary disease | 6.87 (5.62-8.10) | 4.85 (3.86-5.82) | 1.34 (1.07-1.67) | 0.01 |
| Death | 5.68 (4.78-6.57) | 6.68 (5.56-7.79) | 0.91 (0.74-1.12) | 0.38 |
| Fatigue | 19.89 (17.86-21.87) | 19.25 (17.32-21.14) | 0.97 (0.86-1.10) | 0.67 |
| GORD | 26.03 (23.80-28.19) | 25.84 (23.61-28.01) | 1.06 (0.95-1.18) | 0.3 |
| Hair loss | 1.33 (0.76-1.89) | 1.18 (0.74-1.63) | 0.87 (0.52-1.46) | 0.59 |
| Headache | 9.10 (7.66-10.52) | 9.14 (7.67-10.59) | 1.03 (0.85-1.25) | 0.78 |
| Hospitalisation | 23.47 (21.85-25.06) | 22.42 (20.78-24.02) | 1.08 (0.98-1.18) | 0.13 |
| Hypercoagulopathy/DVT/PE | 9.10 (7.83-10.36) | 9.01 (7.79-10.21) | 1.04 (0.87-1.23) | 0.68 |
| Hyperlipidaemia | 56.36 (53.47-59.07) | 52.82 (50.14-55.35) | 1.13 (1.04-1.21) | 0.002 |
| Hypertension | 67.01 (64.17-69.62) | 64.03 (61.33-66.54) | 1.11 (1.04-1.19) | 0.002 |
| Hypoxaemia | 12.62 (11.34-13.88) | 14.14 (12.76-15.51) | 0.92 (0.81-1.05) | 0.2 |
| ICU admission | 6.43 (5.56-7.28) | 6.63 (5.66-7.59) | 1.02 (0.85-1.22) | 0.85 |
| Interstitial lung disease | 3.80 (2.94-4.65) | 4.63 (3.66-5.60) | 0.86 (0.66-1.11) | 0.25 |
| Intubation/Ventilation | 3.11 (2.54-3.68) | 3.38 (2.74-4.02) | 0.96 (0.74-1.24) | 0.76 |
| Ischaemic stroke | 4.82 (3.80-5.83) | 4.92 (3.85-5.99) | 1.03 (0.80-1.33) | 0.81 |
| Joint pain | 18.21 (16.13-20.24) | 18.94 (16.77-21.05) | 1.03 (0.90-1.19) | 0.64 |
| Kidney disease | 23.09 (21.15-24.97) | 22.38 (20.49-24.22) | 1.10 (0.98-1.22) | 0.1 |
| Liver disease | 6.68 (5.63-7.72) | 7.07 (6.02-8.10) | 0.97 (0.79-1.18) | 0.74 |
| Long COVID feature (any) | 65.82 (63.25-68.20) | 67.68 (65.11-70.06) | 1.03 (0.96-1.10) | 0.4 |
| Mood disorder | 19.80 (17.62-21.92) | 17.10 (15.26-18.90) | 1.14 (1.00-1.30) | 0.06 |
| Myalgia | 2.82 (2.00-3.64) | 2.76 (1.89-3.62) | 1.03 (0.72-1.46) | 0.88 |
| Myocarditis | 0.09 (0.002-0.19) | 0.38 (0.10-0.66) | 0.45 (0.14-1.48) | 0.17 |
| Myoneural junction/muscle disease | 1.02 (0.65-1.39) | 1.46 (0.99-1.93) | 0.74 (0.47-1.16) | 0.19 |
| Nerve/nerve root/plexus disorder | 5.29 (4.07-6.50) | 4.28 (3.35-5.20) | 1.01 (0.77-1.31) | 0.96 |
| Obesity | 35.92 (33.54-38.22) | 34.59 (32.25-36.85) | 1.05 (0.96-1.15) | 0.31 |
| Other pain | 16.22 (14.19-18.21) | 17.48 (15.58-19.33) | 0.90 (0.78-1.03) | 0.13 |
| Oxygen requirement | 14.10 (12.62-15.56) | 13.57 (12.20-14.92) | 1.08 (0.95-1.23) | 0.26 |
| Peripheral neuropathy | 6.74 (5.48-7.99) | 5.83 (4.63-7.02) | 1.23 (0.97-1.56) | 0.08 |
| Psychotic disorder | 0.69 (0.39-0.99) | 1.13 (0.70-1.56) | 0.66 (0.39-1.14) | 0.13 |
| Respiratory failure | 12.22 (10.95-13.47) | 14.61 (13.22-15.97) | 0.87 (0.76-0.99) | 0.04 |
| Seizures | 1.66 (1.03-2.28) | 1.89 (1.28-2.49) | 0.75 (0.50-1.14) | 0.17 |

| | | | | |
|---|---|---|---|---|
| Sleep disorders | 20.72 (18.63-22.75) | 20.52 (18.55-22.45) | 1.04 (0.92-1.17) | 0.57 |
| Type 2 diabetes mellitus | 30.74 (28.66-32.75) | 30.13 (28.03-32.17) | 1.08 (0.98-1.19) | 0.11 |
| Urticaria | 0.53 (0.22-0.84) | 0.33 (0.15-0.51) | 1.27 (0.60-2.72) | 0.53 |

**The RECORD statement – checklist of items, extended from the STROBE statement, that should be reported in observational studies using routinely collected health data.**

| | Item No. | STROBE items | Location in manuscript where items are reported | RECORD items | Location in manuscript where items are reported |
|---|---|---|---|---|---|
| **Title and abstract** | | | | | |
| | 1 | (a) Indicate the study's design with a commonly used term in the title or the abstract (b) Provide in the abstract an informative and balanced summary of what was done and what was found | Title and Abstract | RECORD 1.1: The type of data used should be specified in the title or abstract. When possible, the name of the databases used should be included.<br><br>RECORD 1.2: If applicable, the geographic region and timeframe within which the study took place should be reported in the title or abstract.<br><br>RECORD 1.3: If linkage between databases was conducted for the study, this should be clearly stated in the title or abstract. | Abstract (Methods)<br><br>Abstract (Methods) and Appendix p. 1<br><br>N/A |
| **Introduction** | | | | | |
| Background rationale | 2 | Explain the scientific background and rationale for the investigation being reported | Abstract, Research in Context, and Introduction | | |
| Objectives | 3 | State specific objectives, including any prespecified hypotheses | Abstract (Background) and Introduction | | |
| **Methods** | | | | | |
| Study Design | 4 | Present key elements of study design early in the paper | Abstract and Methods (first subsection) | | |
| Setting | 5 | Describe the setting, locations, and relevant dates, including periods of recruitment, exposure, follow-up, and data collection | Methods ('Data and study design', 'Covariates', and 'Outcomes') | | |

| | | | | | |
|---|---|---|---|---|---|
| Participants | 6 | *(a) Cohort study* - Give the eligibility criteria, and the sources and methods of selection of participants. Describe methods of follow-up<br>*Case-control study* - Give the eligibility criteria, and the sources and methods of case ascertainment and control selection. Give the rationale for the choice of cases and controls<br>*Cross-sectional study* - Give the eligibility criteria, and the sources and methods of selection of participants<br><br>*(b) Cohort study* - For matched studies, give matching criteria and number of exposed and unexposed<br>*Case-control study* - For matched studies, give matching criteria and the number of controls per case | Methods ('Data and study design', 'Cohorts', and 'Covariates') and Appendix ('Definition of cohorts' and 'Definition of covariates') | RECORD 6.1: The methods of study population selection (such as codes or algorithms used to identify subjects) should be listed in detail. If this is not possible, an explanation should be provided.<br><br>RECORD 6.2: Any validation studies of the codes or algorithms used to select the population should be referenced. If validation was conducted for this study and not published elsewhere, detailed methods and results should be provided.<br><br>RECORD 6.3: If the study involved linkage of databases, consider use of a flow diagram or other graphical display to demonstrate the data linkage process, including the number of individuals with linked data at each stage. | Methods ('Data and study design', 'Cohorts', and 'Covariates') and Appendix ('Definition of cohorts' and 'Definition of covariates')<br><br>Appendix ('TriNetX network – Quality control')<br><br>N/A |
| Variables | 7 | Clearly define all outcomes, exposures, predictors, potential confounders, and effect modifiers. Give diagnostic criteria, if applicable. | Methods ('Cohorts', 'Covariates', 'Outcomes') and Appendix ('Definition of cohorts', 'Definition of covariates', 'Definition of outcomes') | RECORD 7.1: A complete list of codes and algorithms used to classify exposures, outcomes, confounders, and effect modifiers should be provided. If these cannot be reported, an explanation should be provided. | Methods ('Cohorts', 'Covariates', 'Outcomes') and Appendix ('Definition of cohorts', 'Definition of covariates', 'Definition of outcomes') |
| Data sources/ measurement | 8 | For each variable of interest, give sources of data and details | Methods ('Cohorts', 'Covariates', | | |

| | | | |
|---|---|---|---|
| | | of methods of assessment (measurement).<br>Describe comparability of assessment methods if there is more than one group | 'Outcomes') and Appendix ('Definition of cohorts', 'Definition of covariates', 'Definition of outcomes') |
| Bias | 9 | Describe any efforts to address potential sources of bias | Introduction, Methods ('Cohorts'), and Appendix ('Definition of cohorts') |
| Study size | 10 | Explain how the study size was arrived at | Methods ('Cohorts') |
| Quantitative variables | 11 | Explain how quantitative variables were handled in the analyses. If applicable, describe which groupings were chosen, and why | Methods ('Covariates'), Appendix ('Definition of covariates') |
| Statistical methods | 12 | (a) Describe all statistical methods, including those used to control for confounding<br>(b) Describe any methods used to examine subgroups and interactions<br>(c) Explain how missing data were addressed<br>(d) *Cohort study* - If applicable, explain how loss to follow-up was addressed<br>*Case-control study* - If applicable, explain how matching of cases and controls was addressed<br>*Cross-sectional study* - If applicable, describe analytical | Methods ('Statistical analyses') and Appendix ('Details on statistical analyses') |

| | | | | | |
|---|---|---|---|---|---|
| | | methods taking account of sampling strategy<br>(e) Describe any sensitivity analyses | | | |
| Data access and cleaning methods | | .. | | RECORD 12.1: Authors should describe the extent to which the investigators had access to the database population used to create the study population.<br><br>RECORD 12.2: Authors should provide information on the data cleaning methods used in the study. | Author contribution section<br><br>Appendix ('TriNetX network') |
| Linkage | | .. | | RECORD 12.3: State whether the study included person-level, institutional-level, or other data linkage across two or more databases. The methods of linkage and methods of linkage quality evaluation should be provided. | N/A |
| **Results** | | | | | |
| Participants | 13 | (a) Report the numbers of individuals at each stage of the study (*e.g.*, numbers potentially eligible, examined for eligibility, confirmed eligible, included in the study, completing follow-up, and analysed)<br>(b) Give reasons for non-participation at each stage.<br>(c) Consider use of a flow diagram | Results (first paragraph) and Table 1 and Appendix Tables S1–5. | RECORD 13.1: Describe in detail the selection of the persons included in the study (*i.e.,* study population selection) including filtering based on data quality, data availability and linkage. The selection of included persons can be described in the text and/or by means of the study flow diagram. | Methods ('Cohorts') and Appendix ('Definition of cohorts') |
| Descriptive data | 14 | (a) Give characteristics of study participants (*e.g.*, demographic, clinical, social) and information on exposures and potential confounders | (a) Table 1 and Table S1-S5 | | |

| | | | |
|---|---|---|---|
| | | (b) Indicate the number of participants with missing data for each variable of interest<br>(c) *Cohort study* - summarise follow-up time (*e.g.*, average and total amount) | (b) Table 1 and Table S1-S5<br>(c) Results, first paragraph |
| Outcome data | 15 | *Cohort study* - Report numbers of outcome events or summary measures over time<br>*Case-control study* - Report numbers in each exposure category, or summary measures of exposure<br>*Cross-sectional study* - Report numbers of outcome events or summary measures | Fig. 1-3, and Fig. S1-S9 |
| Main results | 16 | (a) Give unadjusted estimates and, if applicable, confounder-adjusted estimates and their precision (e.g., 95% confidence interval). Make clear which confounders were adjusted for and why they were included<br>(b) Report category boundaries when continuous variables were categorized<br>(c) If relevant, consider translating estimates of relative risk into absolute risk for a meaningful time period | Results section, Fig. 1-3, Fig. S1-S9 and Tables S6 and S8-S13<br>N/A<br>N/A |
| Other analyses | 17 | Report other analyses done—e.g., analyses of subgroups and interactions, and sensitivity analyses | Results section and appendix |
| **Discussion** | | | |
| Key results | 18 | Summarise key results with reference to study objectives | Discussion first paragraph |

| | | | | | |
|---|---|---|---|---|---|
| Limitations | 19 | Discuss limitations of the study, taking into account sources of potential bias or imprecision. Discuss both direction and magnitude of any potential bias | Discussion, antepenultimate paragraph | RECORD 19.1: Discuss the implications of using data that were not created or collected to answer the specific research question(s). Include discussion of misclassification bias, unmeasured confounding, missing data, and changing eligibility over time, as they pertain to the study being reported. | Discussion, antepenultimate paragraph |
| Interpretation | 20 | Give a cautious overall interpretation of results considering objectives, limitations, multiplicity of analyses, results from similar studies, and other relevant evidence | Discussion, last paragraph, Abstract, and Research in Context | | |
| Generalisability | 21 | Discuss the generalisability (external validity) of the study results | Discussion | | |
| **Other Information** | | | | | |
| Funding | 22 | Give the source of funding and the role of the funders for the present study and, if applicable, for the original study on which the present article is based | Declaration of interest | | |
| Accessibility of protocol, raw data, and programming code | | .. | | RECORD 22.1: Authors should provide information on how to access any supplemental information such as the study protocol, raw data, or programming code. | Data sharing section |

*Reference: Benchimol EI, Smeeth L, Guttmann A, Harron K, Moher D, Petersen I, Sørensen HT, von Elm E, Langan SM, the RECORD Working Committee. The REporting of studies Conducted using Observational Routinely-collected health Data (RECORD) Statement. *PLoS Medicine* 2015; in press.

*Checklist is protected under Creative Commons Attribution (CC BY) license.
